# Mutational signatures and increased retrotransposon insertions in Xeroderma Pigmentosum variant skin tumors

**DOI:** 10.1101/2022.07.28.22277756

**Authors:** Camila Corradi, Juliana B. Vilar, Vanessa C. Buzatto, Tiago A. de Souza, Ligia P. Castro, Veridiana Munford, Rodrigo De Vecchi, Pedro A. F. Galante, Fernanda Orpinelli, José L. Buzzo, Mirian N. Sotto, Paulo Saldiva, Jocelânio W. de Oliveira, Sulamita C. W. Chaibub, Alain Sarasin, Carlos F. M. Menck

## Abstract

Xeroderma Pigmentosum variant (XP-V) is an autosomal recessive disease with an increased risk to develop cutaneous neoplasms in sunlight exposed regions. These cells are deficient in the translesion synthesis DNA polymerase eta. Eleven skin tumors from a genetic cluster of XP-V patients had their exome sequenced. Mutational signatures identified for most tumors were related to ultraviolet exposure, such as C>T transitions targeted to pyrimidine dimers. However, four samples carry different mutational signatures, with C>A mutations associated with tobacco usage. Basal cell carcinomas showed a distinct C>A mutation spectra reflecting a novel mutational signature. Higher levels for retroposon insertions were detected in the XP-V tumors, compared to non-XP skin tumors. The results reveal other possible causes for XP-V tumors and the involvement of polymerase eta in suppressing retrotransposition. The expected high mutation burden, found in most of these tumors, renders these XP patients good candidates for immunotherapy with checkpoint blockers.

## Introduction

When defined as a single entity, skin cancer is the most common type of cancer worldwide. Both melanoma and non-melanoma forms have increased at an alarming rate in the recent decades, probably due to a combination of factors such as increased exposure to the ultraviolet (UV) component of sunlight, decreased ozone layer and the increase in human longevity^1,2^. Skin cancers are particularly problematic in Xeroderma Pigmentosum (XP) patients. XP is a rare autosomal recessive disorder characterized by extreme sensitivity of their skin to sunlight exposure, with hyper and hypopigmentation, xerosis, ocular abnormalities, and precancerous lesions like actinic keratosis; most patients develop skin cancer before the age of 10 years old.

Cells from these patients are usually defective in removing UV-induced DNA lesions by nucleotide excision repair (NER), commonly called classical XPs (XP-A to XP-G, depending on the mutated gene). However, some XP patients have normal NER but are deficient in the translesion synthesis (TLS) DNA polymerase eta (Pol η or Pol eta), which promotes the error-free bypass of UV-induced DNA lesions during replication and are called XP-variant (XP-V). Currently, there is no specific treatment or prophylaxis for these patients, except surgical intervention to remove the tumors and complete sun protection. Some XP patients present a 100-fold increased frequency for internal tumors, probably due to endogenous DNA damage^3–5^.

XP incidence can vary significantly among countries, such as 1:20,000 in Japan to 1:250,000 in the US and 1:500,000 in Western Europe^6,7^. In Latin America, the data is scarce, and the patients suffer from poor diagnosis and high sunlight incidence, especially in tropical countries. However, recent reports identify several XP mutations in Brazil^8,9^. In West-central Brazil (Goias State, municipality of Faina), a community was identified as one of the densest populations with XP (17 patients among 7,000 inhabitants), due to consanguineous marriage. This genetic cluster involves the variant form of XP (XP-V, with mutations in the *POLH* gene; OMIM#278750), with two independent founder mutations^9^. Both mutations rendered the expected XP-V cellular phenotype, with normal DNA damage removal but abnormal DNA synthesis after UV irradiation and deficient expression of the Pol eta protein^9^.

Pol eta is responsible for bypassing DNA lesions induced by UV light, such as CPDs (Cyclobutane Pyrimidine Dimers, the most frequent UV-induced DNA lesion). In vitro, Pol eta correctly bypasses T-containing dimers^10^. However, UVA-irradiated XP-V cells have a substantial increased frequency of C>T mutations targeted at pyrimidine dimers, indicating that the primary function of Pol eta in the cells is to correctly replicate C-containing dimers inserting G opposite the damaged base^11^. Interestingly, C>A mutations were also identified in non-irradiated and UVA-irradiated XP-V cells, indicating that this polymerase also has roles in suppressing mutagenesis of oxidized bases. In fact, Pol eta bypasses other types of DNA damage, such as 8-oxodeoxyguanosine (8-oxodG) and platinum-based anticancer drugs, such as cisplatin and oxaliplatin, indicating the involvement of this protein in the tolerance to these agents and suggesting that it might also be involved in tumor resistance to anti-tumoral drugs^12,13^.

The XP-V patients of the genetic cluster identified at the Goias State in Brazil are followed with medical attention, and eleven skin tumor samples were obtained from six patients. These tumor samples had their whole-exome sequenced to identify XP-V skin cancers’ mutational profile from this unique, highly endogamic community. The results show a high mutation burden in most samples and indicate a high variation of genes affected by nonsynonymous mutations, with cancer driver genes identified, revealing potential genetic events necessary for tumor transformation. The mutation analyses disclosed that seven tumor samples presented the typical skin cancer mutation signature, including C>T mutations at dipyrimidine sites. Other mutations were also found in these samples, especially C>A transversions that indicate a possible different mutation signature and mutation motif that could be specific for skin tumors in XP-V patients. However, four of the skin tumor samples displayed a pattern of mutation not related to sunlight exposure. At least three of them may be related to tobacco chewing habits, suggesting that XP-V patients are also susceptible to these DNA damaging habits. Moreover, somatic retrocopies and mobile elements insertions were also identified, calling attention to potential effects of retrotransposition in the absence of Pol eta.

## Results

### XP-V tumors’ samples features and single base substitution mutations analysis

The count of point mutations is presented in Table 1, where mutations in tumors exclude the germline mutations detected also in the saliva DNA. Single nucleotide variation (SNVs) and small insertions and deletions (INDELs) were the main mutations detected. Mutation count varies from sample to sample, but the tumors revealed an average of 93% of SNVs. All basal cell carcinomas presented over 95% of SNVs (INDELs ∼5%), while the average for the other two types of skin cancer, the squamous cell carcinoma and the melanoma, did not exceed 86% (INDELs ∼14%) (Table 1). According to Foundation Medicine and FDA, the tumor mutation burden (TMB) is considered high when ten or more mutations per sequenced megabase are identified^14^. Ten of these tumors presented, accordingly, a high TMB.

**Table 1:** Somatic mutations identified by whole-exome sequencing for tumor samples.

| Sample | Mutations<br>(count) | SNVs<br>(count) | INDELs<br>(count) | TMB<br>(high $\geq 10$ mut/Mb) |
| --- | --- | --- | --- | --- |
| XP03GO | 1353 | 1296 | 57 | 30.28 |
| XP06GO_1 | 2766 | 2658 | 108 | 61.82 |
| XP06GO_2 | 6388 | 6229 | 159 | 143.59 |
| XP06GO_3 | 588 | 539 | 49 | 13.13 |
| XP06GO_4 | 591 | 544 | 47 | 13.03 |
| XP11GO | 486 | 410 | 76 | 10.84 |
| XP45GO | 1782 | 1689 | 93 | 40.12 |
| XP52GO_1 | 4039 | 3960 | 79 | 91.11 |
| XP52GO_2 | 2799 | 2711 | 88 | 62.34 |
| XP85GO_1 | 400 | 346 | 54 | 8.81 |
| XP85GO_2 | 844 | 739 | 105 | 18.97 |

The mutation data from tumor samples exclude the mutations found in the saliva, which are considered germline mutations. Total somatic mutations combine SNVs and INDELs detected at exonic and splicing sites. TMB, tumor mutation burden, was calculated as the sum of all mutation types divided by sequenced megabase for each sample.

Table 2 presents the types of mutation identified for all tumor samples, focusing on the most frequent ones (C>T and C>A) and the tandem CC>TT mutations. As expected for sunlight-induced skin tumors, seven tumor samples have most C>T transitions and these mutations are located at dipyrimidine sites (YY) (Y = T or C). Also, most of these tumors displayed an increased tandem CC>TT mutations. However, the tumor samples XP11GO, XP45GO, XP85GO_1, and XP85GO_2 presented higher C>A transversions levels over C>T transitions, and tandem CC>TT mutations were very low. Interestingly, XP45GO tumor is a BCC, as the other seven, whilst XP11GO tumor is a SCC, and XP85GO_1 and XP85GO_2 are melanomas. The frequencies of C>T mutations found at YY sequences were also lower than what was observed for the other tumor samples (Table 2). The frequencies of the other four types of mutations (C>G, T>A, T>C, and T>G) are shown in Supplementary Table 1. Additionally, it was noticed that three tumor samples (XP11GO, XP85GO_1, and XP85GO_2) had a slightly higher proportion of T>C mutations.

**Table 2:** Point mutation types C>A, C>T and tandem CC>TT for each tumor sample.

| Sample | C>A (%) | C>T (%) | CC>TT (%) | YY | % C>T in YY |
| --- | --- | --- | --- | --- | --- |
| XP03GO | 443 (34) | 643 (50) | 106 (8) | 557 | 87 |
| XP06GO_1 | 795 (30) | 1580 (59) | 456 (17) | 1432 | 91 |
| XP06GO_2 | 1489 (24) | 4212 (68) | 986 (16) | 3923 | 93 |
| XP06GO_3 | 148 (27) | 281 (52) | 92 (17) | 231 | 82 |
| XP06GO_4 | 168 (31) | 252 (46) | 100 (18) | 206 | 82 |
| XP11GO* | 190 (46) | 102 (25) | 2 (0) | 62 | 61 |
| XP45GO* | 922 (55) | 522 (31) | 126 (7) | 436 | 84 |
| XP52GO_1 | 1169 (30) | 2358 (60) | 472 (12) | 2287 | 97 |
| XP52GO_2 | 780 (29) | 1644 (61) | 504 (19) | 1529 | 93 |
| XP85GO_1* | 135 (39) | 89 (26) | 14 (4) | 54 | 61 |
| XP85GO_2* | 487 (66) | 117 (16) | 16 (2) | 83 | 71 |

\* These tumor samples presented a different proportion of C>T transitions and C>A transversions.

### Somatic mutational spectra and mutational signatures for XP-V tumors’ samples

The tumor mutation spectra for the eleven XP-V tumors are presented in Fig. 1. For seven of these tumors (XP03GO, XP06GO_1, XP06GO_2, XP06GO_3, XP06GO_4, XP52GO_1, and XP52GO_2), the pattern of C>T transitions at dipyrimidine sites was observed (Fig. 1a). Also, for these seven tumor samples plus XP45GO, the C>A mutations presented clear peaks at A**C**A, C**C**A, G**C**A, and T**C**A (Fig. 1a). Interestingly, and unexpected for skin cancers, four of the tumors (XP11GO, XP45GO, XP85GO_1, and XP85GO_2) did not show the typical C>T mutational spectra at dipyrimidine sites. Two distinctive mutational signatures explained approximately 96% of the data: signatures S1 and S2, shown in Fig. 1b. Samples XP03GO, XP06GO_1, XP06GO_2, XP52GO_1, and XP52GO_2 exhibit a profile like signature S1, with C>T mutations occurring at dipyrimidine sites and the clear peaks at C>A, as explained above. Samples XP11GO, XP85GO_1 and XP85GO_2 have a profile similar to signature S2, which has a higher and more diffused contribution of C>A transversions. Samples XP06GO_3, XP06GO_4 and XP45GO shared these two signatures (Fig. 1b,c).

**Figure 1:**
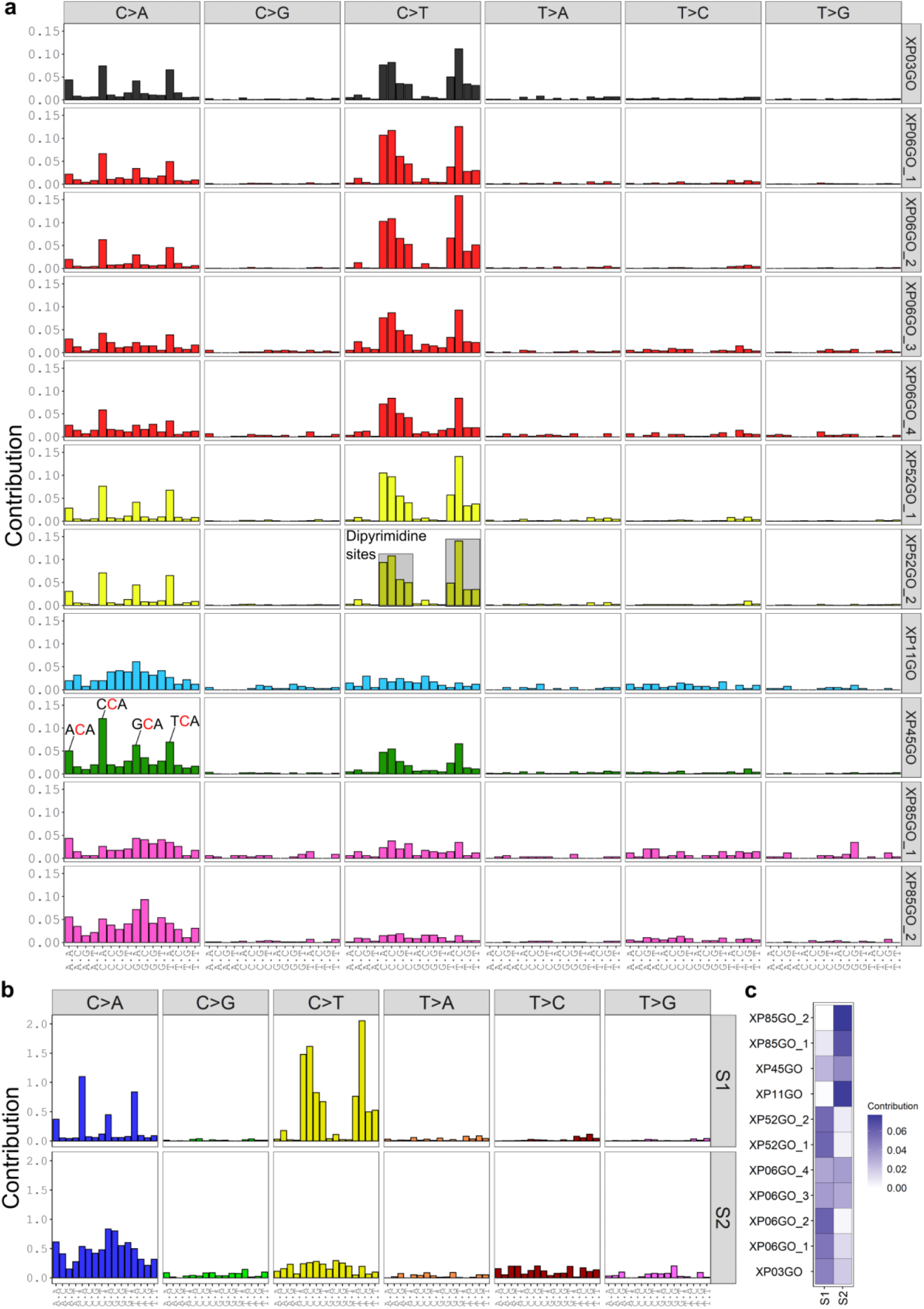
The somatic mutation spectra and mutation signatures for the XP-V tumor samples. (a) Somatic mutational spectra for each tumor sample; (b) mutational signatures S1 and S2 extracted from SNVs explain 96% of the data; and (c) tumor sample classification for those two signatures.

### Sequence motifs for C>T and C>A mutations in XP-V skin tumors

The sequence context for C>T and C>A was also evaluated with the logo representation obtained with pLogo^15^. The results for C>T mutations are shown in Fig. 2. Eight tumors, XP03GO, XP06GO_1, XP06GO_2, XP06GO_3, XP06GO_4, XP52GO_1, XP52GO_2, and XP45GO had C>T transitions located at pyrimidine rich sequences (Fig. 2a,b,c,d,e,f,g,i), mainly at the TY**C**CT motif (except for sample XP06GO_4, which did not present enrichment of the last T at position +2 (Fig. 2e)). The samples XP85GO_1 and XP85GO_2 had statistically significant enrichment of a C base at position -1 (Fig. 2j,k), and there was no significant sequence context enrichment for C>T mutations for the tumor sample XP11GO (Fig. 2h).

**Figure 2:**
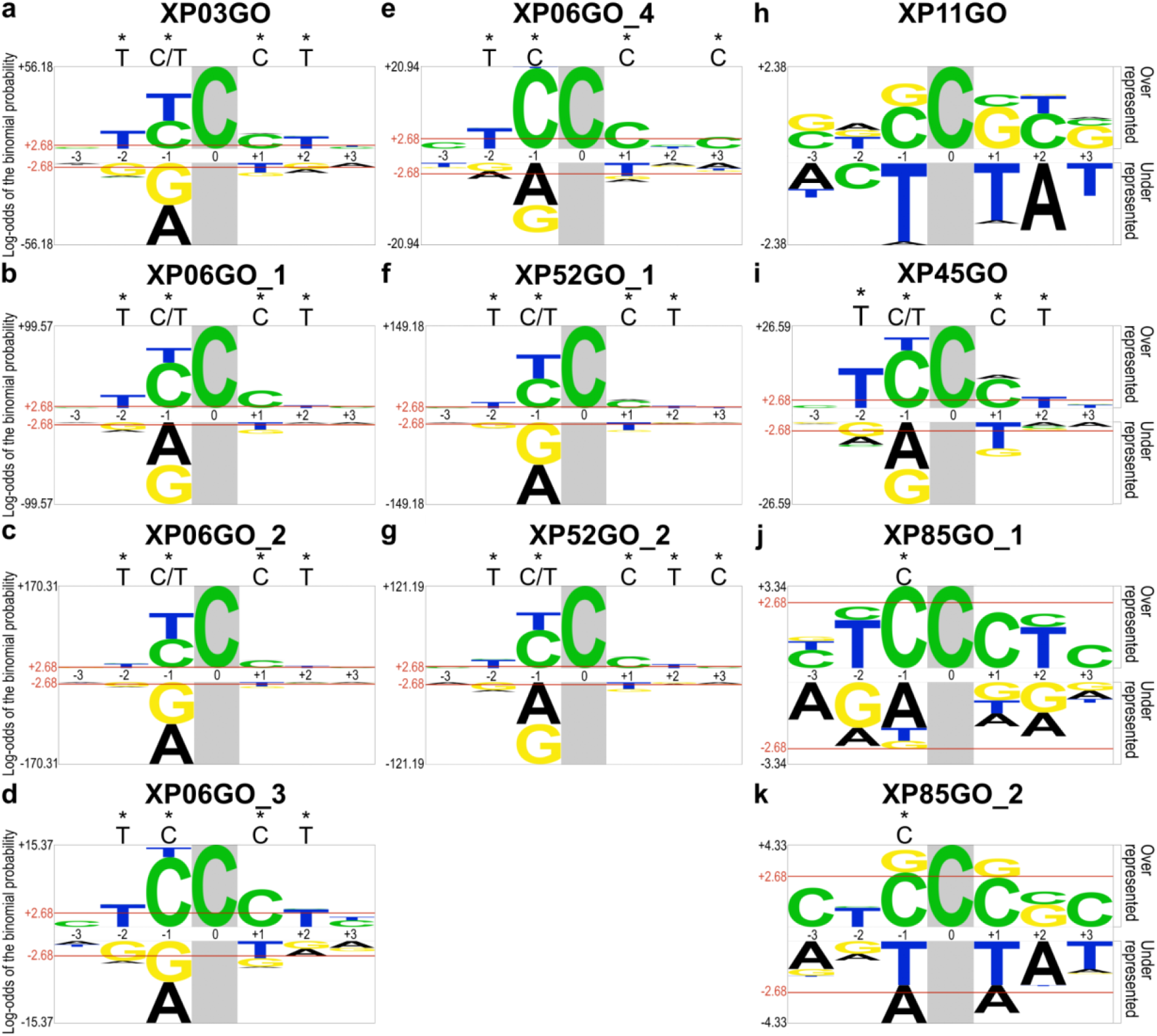
Sequence context for C>T point mutations detected in the XP-V tumor samples. Probability Logo Generator (pLogo, v1.2.0) was addressed to examine the sequence context adjacent to C>T mutations (highlighted in grey background). The red bar at 2.68 binomial probability corresponds to the Bonferroni-corrected statistical significance values (p = 0.05) and the base above the graphic highlighted (*) had a significant enrichment. Aligned foreground (n(fg)) sequences used to generate the image: (a) XP03GO n(fg) = 451, (b) XP06GO_1 n(fg) = 898, (c) XP06GO_2 n(fg) = 1466, (d) XP06GO_3 n(fg) = 242, (e) XP06GO_4 n(fg) = 210, (f) XP52GO_1 n(fg) = 1136, (g) XP52GO_2 n(fg) = 942, (h) XP11GO n(fg) = 95, (i) XP45GO n(fg) = 383, (j) XP85GO_1 n(fg) = 68, (k) XP85GO_2 n(fg) = 106. 4096 background sequences used (human exome).

The sequence context for the C>A mutations is shown in Fig. 3. Interestingly, all samples except for XP11GO and XP85GO_1 (Fig. 3h,j) had a significant bias for an A at 3’ of the mutation, position +1 (Fig. 3a,b,c,d,e,f,g,I,k), an interesting mutational profile of basal cell carcinoma of XP-V patients, since only one of these tumors is a melanoma (XP85GO_2). Less clear, but significant Cs were detected in positions -1 and +3 (Fig. 3b,c,f,g,i). For the three XP-V tumor samples (XP11GO, XP85GO_1, and XP85GO_2), a G was significantly found at 5’ (position -1) of the C>A mutation (Fig. 3h,j,k). In summary, the results suggest a motif targeted for C>A mutations in XP-V tumors defined as C**C**ANC, or (more frequently) **C**A, especially for basal cell carcinomas.

**Figure 3:**
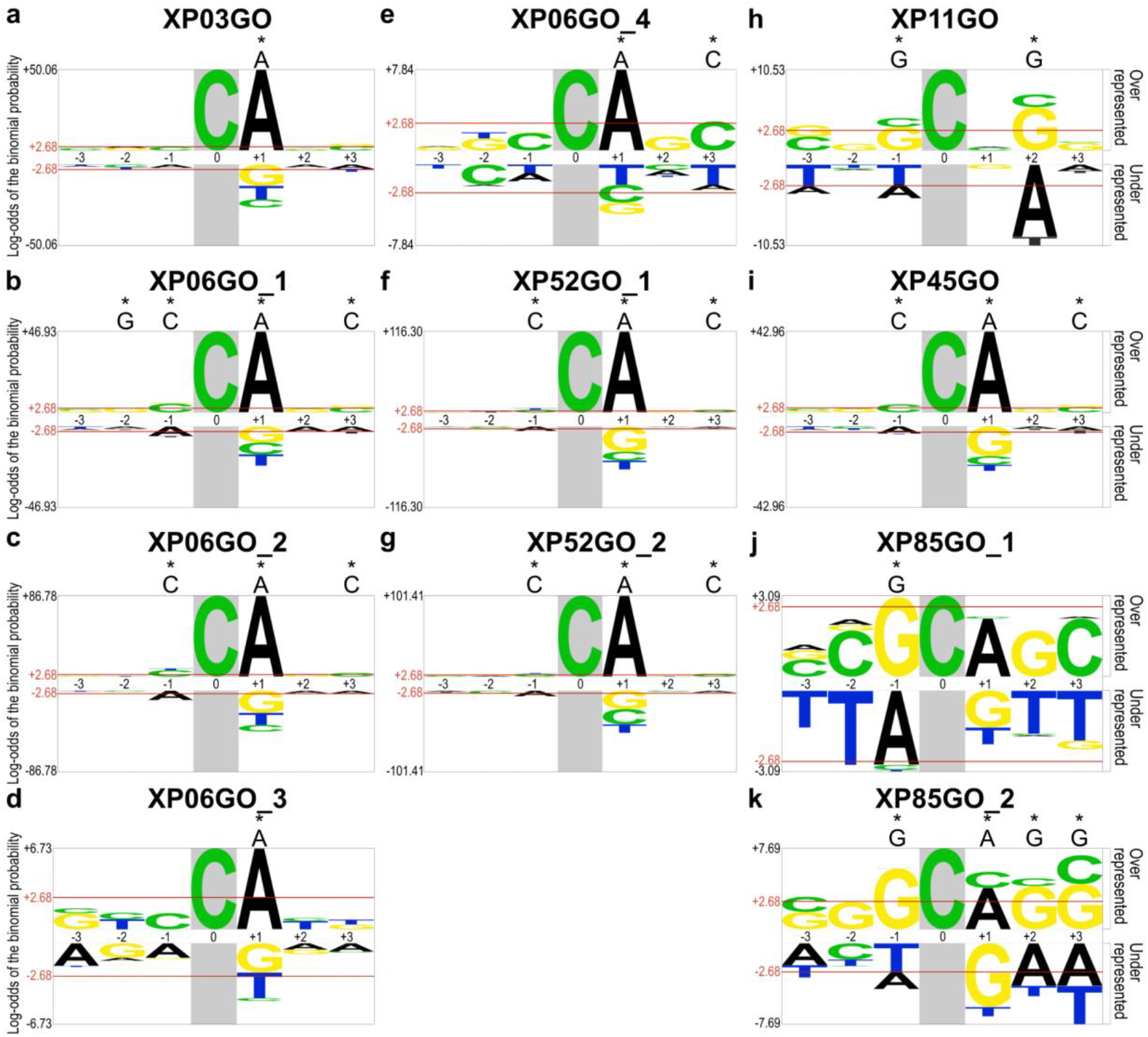
Sequence context for C>A point mutations in XP-V tumor samples. Probability Logo Generator (pLogo, v1.2.0) was addressed to examine the sequence context adjacent to C>A mutations (highlighted in grey background). The red bar at 2.68 binomial probability corresponds to the Bonferroni-corrected statistical significance values (p = 0.05) and the nucleotide above the graphic highlighted (*) had a significant enrichment. Aligned foreground (n(fg)) sequences used to generate the image: (a) XP03GO n(fg) = 365, (b) XP06GO_1 n(fg) = 589, (c) XP06GO_2 n(fg) = 933, (d) XP06GO_3 n(fg) = 128, (e) XP06GO_4 n(fg) = 149, (f) XP52GO_1 n(fg) = 796, (g) XP52GO_2 n(fg) = 556, (h) XP11GO n(fg) = 175, (i) XP45GO n(fg) = 704, (j) XP85GO_1 n(fg) = 114, (k) XP85GO_2 n(fg) = 399. 4096 background sequences used (human exome).

### Somatic mutational signatures

The mutational somatic signature from single base substitution (SBS) was recovered from COSMIC public database (v 3.2)^16^ with XP-V tumor data, using the package MutationalPatterns^17^. Figure 4a displays the 10% most represented mutational signatures obtained, where the seven first tumor samples presented a highly homogenous signature attribution, with signatures SBS7A and SBS7B (UV exposure) responsible for more than 30% of all signatures found for those samples. In addition, SBS18 (damage by ROS) also appears in these seven tumor samples. However, the other four samples, XP11GO, XP45GO, XP85GO_1, and XP85GO_2, had much lower contributions of these signatures. Interestingly, these samples had significant contributions of SBS29 (tobacco chewing) and SBS24 (aflatoxin exposure), specially XP11GO, XP85GO_1 and XP85GO_2, probably because of the significant levels of C>A mutations. Samples XP11GO and XP85GO_2 had a contribution of 35% and 45% of SBS29 signature and SBS24 signature represented 20% and 34% of samples XP85GO_1 and XP85GO_2, respectively.

**Figure 4:**
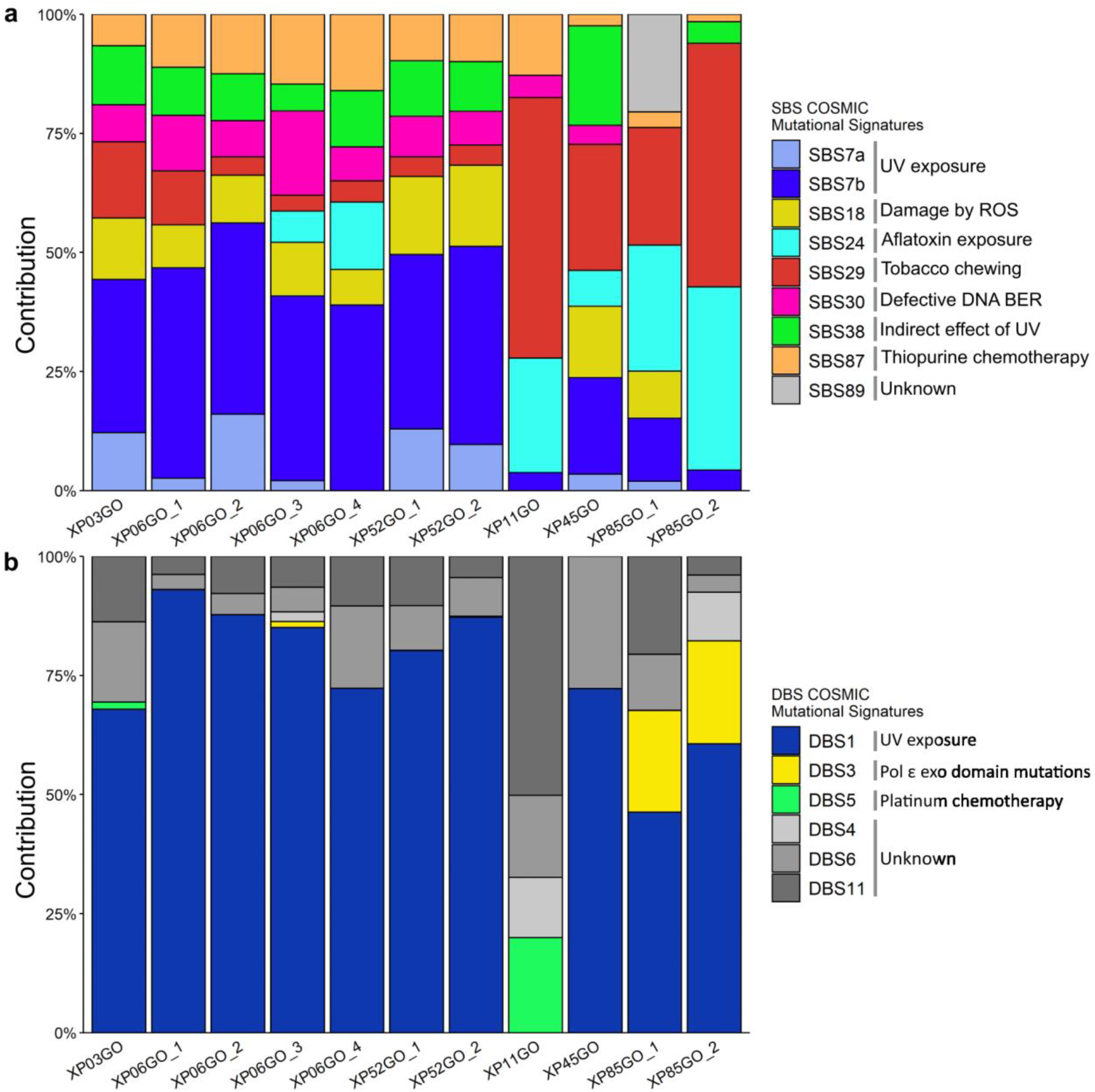
Mutational somatic signature profile for XP-V tumor samples. (a) Single base substitutions (SBS) and (b) double base substitutions (DBS). The signatures were recovered from the COSMIC database (v 3.2) and calculated by MutationalPatterns, a Bioconductor package. The most represented signatures in the data (more than 10%) are shown.

An unsupervised clustering was performed to investigate how these mutations were grouped considering the primary mutation somatic signatures detected for the skin tumors and saliva samples against COSMIC mutational somatic signatures. The results are shown in Supplementary Fig. 1. Clearly, tumor samples are grouped as discussed above, discriminating two clusters, representing the seven ones with mutation signatures indicating the strong influence of C>T in dipyrimidine sites (specially SBS7a, SBS7b and SBS30) and the other four carrying mainly the C>A mutations (SBS18, SBS24 and SBS29). The six saliva samples were clustered together, well separated from the tumor samples.

The contribution of COSMIC double base substitution signatures (DBS) was also investigated. Clearly, except for XP11GO, the tumor samples strongly contributed to DBS1, related to UV exposure and CC>TT tandem substitutions (Fig. 4b). For the sample XP11GO, there was a small contribution of DBS5 (20%), related to platinum chemotherapy, and most of its other signatures were of unknown origins. However, it is important to highlight that the number of double mutations is very small for all tumors, the frequency for the first seven tumors ranged between 6 and 13% and did not go beyond 5% for the last four tumors (Supplementary Table 3).

### Mutation bias for lesions in DNA replicating leading or lagging strands

Considering the pyrimidine at the strand being replicated by the continuous (leading strand) or discontinuous (lagging strand) synthesis, the six types of mutations were evaluated for any bias that could appear considering the DNA damage position. Curiously, the most common mutations (C>T and C>A) as well as T>G, did not show any significant difference when replicated in both strands (Table 3). However, C>G, T>A and T>C, which are not frequent mutations in the XP-V tumors, show significant bias indicating that more mutations are observed when the Ts are being replicated in the lagging strand (Table 3). The sequence contexts of these mutations in the leading and lagging strand were analyzed with pLogo, and the results are shown in the supplementary data. The analysis was performed twice, once considering seven tumor samples (XP03GO, XP06GO_1, XP06GO_2, XP06GO_3, XP06GO_4, XP52GO_1, XP52GO_2) and other with all tumor samples grouped.

**Table 3:**
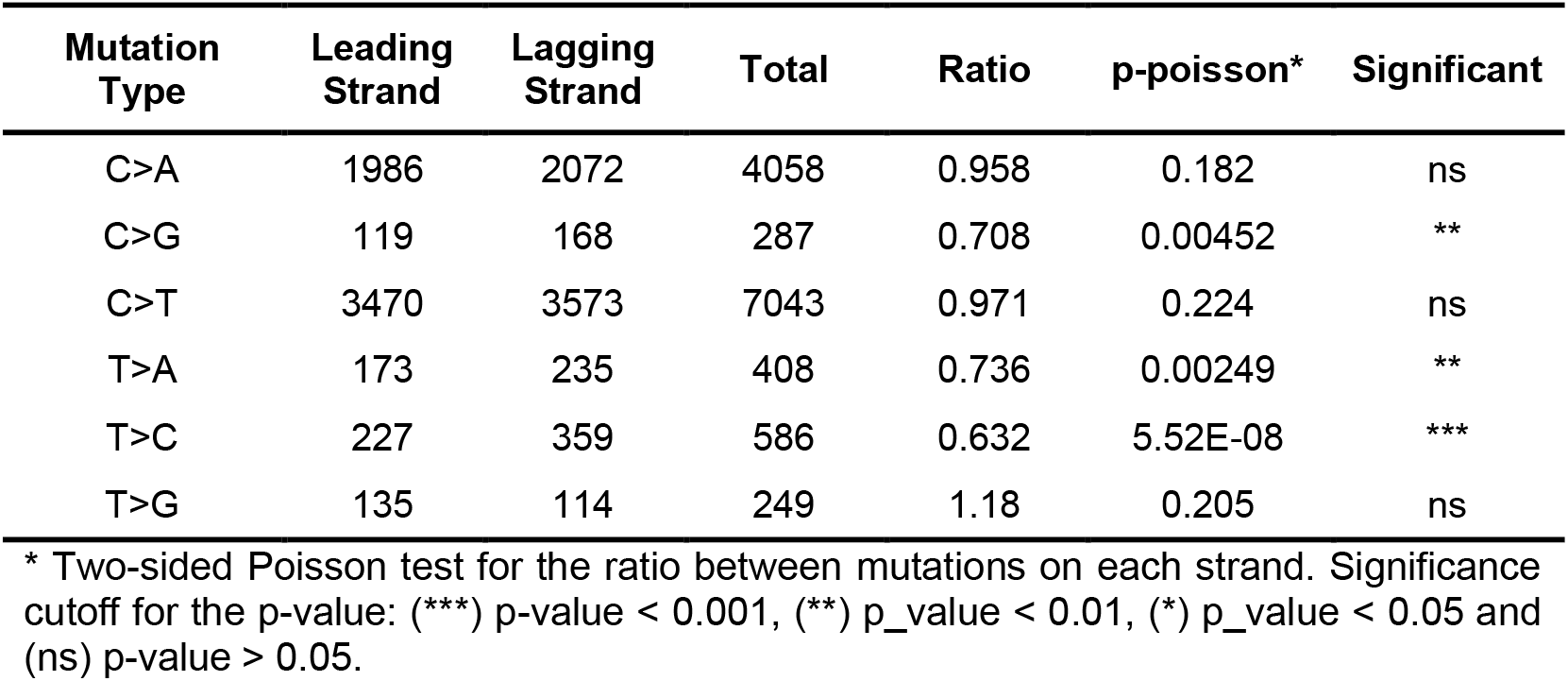
Strand position of the mutations, considering semicontinuous DNA replication.

| Mutation Type | Leading Strand | Lagging Strand | Total | Ratio | p-poisson* | Significant |
| --- | --- | --- | --- | --- | --- | --- |
| C>A | 1986 | 2072 | 4058 | 0.958 | 0.182 | ns |
| C>G | 119 | 168 | 287 | 0.708 | 0.00452 | ** |
| C>T | 3470 | 3573 | 7043 | 0.971 | 0.224 | ns |
| T>A | 173 | 235 | 408 | 0.736 | 0.00249 | ** |
| T>C | 227 | 359 | 586 | 0.632 | 5.52E-08 | *** |
| T>G | 135 | 114 | 249 | 1.18 | 0.205 | ns |

\* Two-sided Poisson test for the ratio between mutations on each strand. Significance cutoff for the p-value: (\*\*\*) p-value < 0.001, (\*\*) p-value < 0.01, (\*) p-value < 0.05 and (ns) p-value > 0.05.

The C>G mutations were found within a pyrimidine rich motif in both strands, specifically at TY**C** in the lagging strand for seven tumor samples (Supplementary Fig. 2a) and TC**C**A for all tumor samples grouped (Supplementary Fig. 2b) and C**C** in the leading strand (Supplementary Fig. 5a,b). The T>A mutations occurred mainly with a T in the 5’ of the mutated base for both strands: lagging strand presented the motif T**T**R (R = A or G) (Supplementary Fig. 3a,b) and motif TT**T**R was observed in the leading strand (Supplementary Fig. 6a,b), similar to what was observed for T>C mutations, where motif TT**T**GT was found in the lagging strand (Supplementary Fig. 4a,b) and T**T**G in the leading one (Supplementary Fig. 7a,b). These results suggest that these mutations could also have occurred at dipyrimidine sites, that is, due to the replication of pyrimidine dimers in both strands.

### INDEL analysis in XP-V tumor samples

Small INDELs were detected in the exome of the eleven tumor samples. Typically, tumor samples presented an average of 4.6% INDELs among all mutations found (SNVs + INDELs). However, this proportion was higher for samples XP11GO, XP85GO_1, and XP85GO_2, due to the low frequency of SNVs in these tumors (Table 1). In addition, the INDEL size for all samples indicated that most are smaller than 10 bp, with 66% having 1 bp of length (Supplementary Table 4).

### Mobile elements profile in XP-V patients

Transposable elements comprise more than half of the human genome^18^, and their mobilization, still active for some retroelements, is a significant source of genomic variations and human diseases^19,20^, including cancer^21^. Thus, we investigated non-reference (polymorphic and somatic) insertions of L1, *Alu* and SVA (here, called Mobile Elements Insertions, MEIs^22^) and retrocopies (here, called retroCNVs^23^) in this cohort of XP-V patients. These mobile genetic structures were searched in saliva and XP-V skin tumors samples. First, to test the approach to identify MEIs and retroCNVs, polymorphic events were searched, which comprise MEIs and retroCNVs shared among these samples and/or those already reported in the literature^23,24^. A large number of MEIs (64 distinct events) and retroCNVs (11 distinct events) were detected, confirming the strategy to identify these classes of insertions (Fig. 5a and Supplementary Table 5).

**Figure 5:**
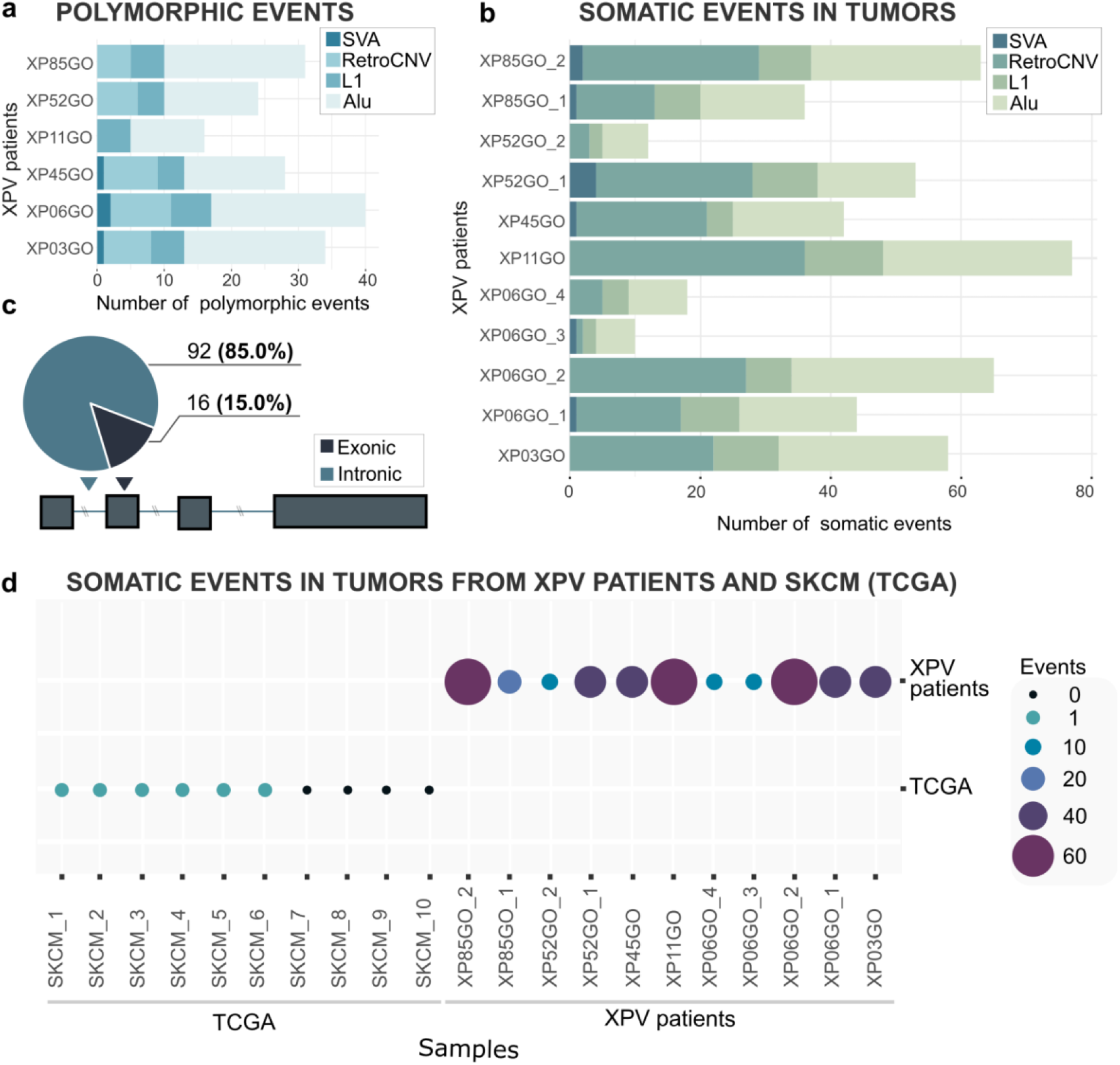
Mobile Elements and retroCNVs insertions in the genomes of our XP-V cohort. (a) Polymorphic MEIs and retroCNVs per individual; (b) somatic insertions of MEIs and retroCNVs in the eleven skin tumor samples from XP-V individuals; (c) gene context (intronic or exonic) of retroCNV insertions into XP-V tumors; (d) number of insertions (MEIs and retroCNVs) in XP-V skin tumors and in a random set of non-XP-V Skin Cancer Melanoma individuals from TCGA.

Next, somatic MEIs and retroCNVs were searched for in the set of eleven tumor samples. We found 285 MEIs (Fig. 5b, Supplementary Table 6), being novel insertions of *Alu* elements and L1 as the most frequent (200 and 75, respectively). Interestingly, we also found putative somatic insertions of SVA (10) and retroCNVs (193), (Fig. 5b and Supplementary Table 6) including retroCNVs insertion into intronic (92 events) and exonic (16 events) regions in tumor samples (Fig. 5c and Supplementary Table 7). The number of retrotransposition insertions in the XP-V tumors was highly variable among the XP-V tumors (a range of 12 to 79 insertions of MEIs plus retroCNVs per sample (Supplementary Table 6)). However, the average of these retrotransposon insertions is much higher when compared to the exome of Skin Cancer tumors from non-XP patients available in The Cancer Genome Atlas (Fig. 5d and Supplementary Table 8).

### Mutated cancer genes’ analyses

Mutations affecting the translated proteins (non-synonymous and stop codons) were identified in 5771 genes in the eleven tumor samples. Among all mutated genes, 301 genes had five or more mutations (Supplementary Table 9). Considering only the ten genes with more mutations, at least three (*TTN, MUC16*, and *ABCA13*) are large genes (more than 5,000 aa), and thus the high number of mutations is probably due to stochastic events. However, the two genes with more mutations are *ZNF717* (539 aa, mutated in all eleven tumors) and *CDC27* (839 aa, mutated in ten out of eleven tumors, not mutated for XP52GO_1) are smaller and interesting genes. Both proteins encoded by these genes are related to DNA processing (*ZNF17* is a Zn-finger protein involved in transcription regulation) and the cell cycle (*CDC27* is part of a complex responsible for the ubiquitin-mediated proteolysis of B-type cyclins). Both have been detected as mutated and indicated as potential tumor gene drivers^25–27^. Three of the other genes with more mutations, *IGSF3* (1194 aa) also appear in the eleven tumors, while *PABPC3* (631 aa) and *TEKT4* (435 aa) occur in seven (out of eleven) tumors. Although these three genes have been related to tumor formation, their roles may be indirect^28–31^. Two other genes, *TAS2R30* (319 aa, found in 6 tumors) and *OFD1* (360 aa, found in 5 tumors), seem interesting but have not been correlated with tumors yet. Curiously, only *MUC16* gene was reported as a putative cancer gene from Tier 2 of the COSMIC Cancer Gene Census. It is a cell surface associated oncogene found in head and neck squamous cell carcinoma and melanomas, mutated in eight of eleven tumor samples.

According to the COSMIC Cancer Gene Census^32^, 289 (Supplementary Table 10) were identified as cancer genes from all genes carrying mutations. All eleven tumors present mutations in genes listed at Supplementary Table 10. The COSMIC Cancer Gene Census identifies each gene as an oncogene, tumor suppressor gene (TSG), or fusion gene, depending on its somatic mutation profile and potential functional role in oncogenesis^32^. Interestingly, considering all the genes, the XP-V tumor samples had a similar proportion of oncogenes and TSGs (Fig. 6a) and curiously, only 14% of those genes are usually found in skin cancer or melanoma from DNA repair proficient patients (Fig. 6b). The genes identified with more than five mutations (301) had their major biological functions identified using the DAVID platform^33^. Four different pathways were retrieved from KEGG^34^ with low p-value (Supplementary Table 11) and the protein digestion and absorption pathway were the most significant ones with 17 genes involved (Supplementary Table 12), and the extracellular matrix (ECM) receptor interaction pathway had 13 of the gene lists involved (Supplementary Table 13).

**Figure 6:**
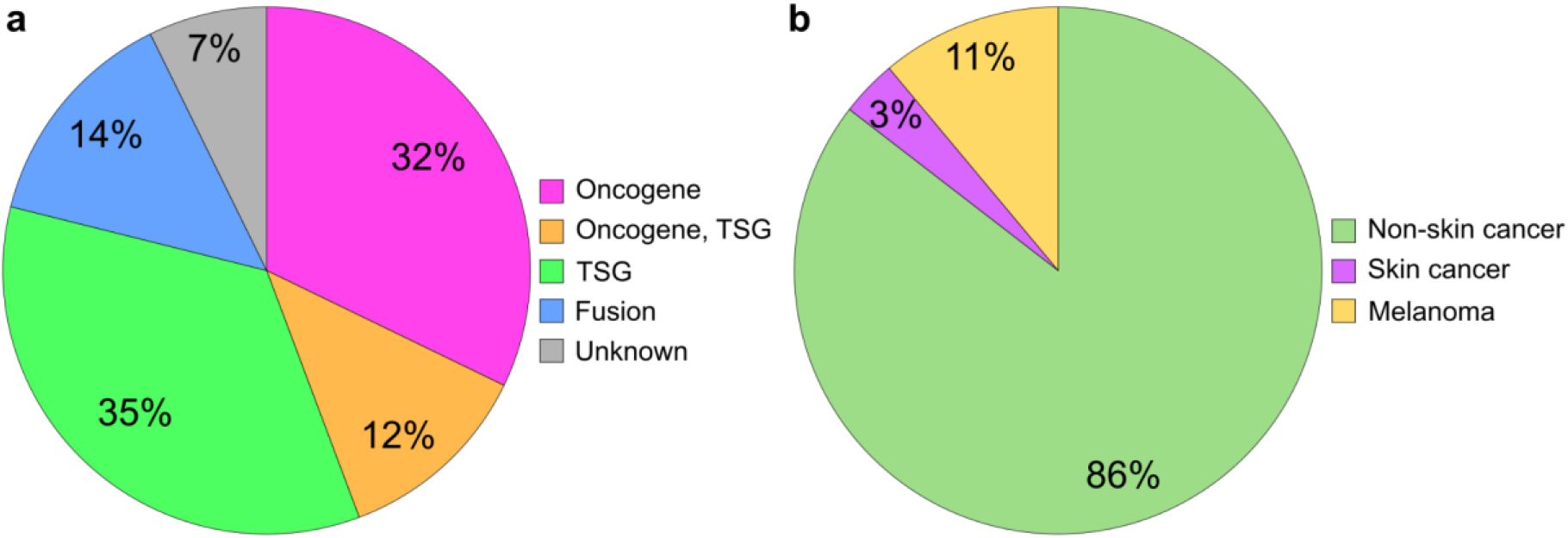
Summarized mutated cancer genes data filtered based on Cancer Gene Census, COSMIC v94. (a) Count of mutated cancer genes divided into five groups: “Oncogene”, “Oncogene, TSG”, “TSG”, “Fusion” and “Unknown”. (b) Count of mutated cancer genes divided into non-skin cancer, skin cancer and melanoma.

Three clusters were obtained from the Gene Ontology database^35^ (GO): GOTERM_BP_DIRECT, GOTERM_MF_DIRECT, GOTERM_CC_DIRECT, which stands for Biological Process, Molecular Function, and Cellular Component, respectively. Among the most significant biological processes detected in the mutated gene list (Supplementary Table 14), extracellular matrix organization was the most significant, with 28 genes (Supplementary Table 15), and collagen fibril organization involved a total of 14 genes (Supplementary Table 16). The main molecular functions in the mutated gene list are presented in Supplementary Table 17: extracellular matrix components and calcium ion binding were the most significant. The 22 genes of the extracellular matrix structural constituent playing a role are listed in Supplementary Table 18; the calcium ion binding molecular function with most genes participating (44 genes) (Supplementary Table 19). Moreover, the more significant cellular components were identified from the mutated gene list (Supplementary Table 20). Again, extracellular matrix was the most significant, with 28 genes involved (Supplementary Table 21), and plasma membrane had a total of 123 genes participating (Supplementary Table 22).

## Discussion

XP patients have defective DNA repair and tolerance mechanisms that process sunlight DNA damage, and one of the leading clinical consequences is the dramatic increase in skin cancer incidence. Investigating the mutations in the DNA of these tumors may disclose the most likely types of DNA damage and mechanisms responsible for their origins. Recent work was performed on internal tumors (leukemia and sarcoma) of XP-C patients, defective in nucleotide excision repair^36^. The results demonstrated a high mutation burden in XP tumors and a causal role for the DNA repair deficiency for endogenous lesions. Our work focused on XP-V skin tumors from a genetic cluster recently described in Brazil^9^, reducing eventual effects from human genetic background variation, except for their deficiency in the TLS polymerase eta. Unlike the work on internal tumors, the patients were highly exposed to well-characterized environmental factors, including high sunlight incidence.

The results confirm that most of the tumors have increased levels of base substitutions, and seven out of eleven tumors present mutations that primarily reflect skin cancer signatures (SBS7A and SBS7B), demonstrating that the lack of Pol eta results in C>T mutations at dipyrimidine sites. Most of the mutations are in sequence context rich in pyrimidines (TY**C**CT motif), pointing to an origin due to sunlight DNA damage, mainly CPDs. These tumors also show a high frequency of CC>TT tandem mutations, a consequence of CPDs and typical of sunlight-induced skin cancers. As observed in XP-V human cells following UVA irradiation in vitro^11^, few mutations were found on Ts, indicating that even in the absence of Pol eta, the human cells replicate T-containing CPDs correctly. Thus, Pol eta is responsible for replicating C-containing dimers, correctly inserting a G opposite to them. These results agree with the hypothesis that Pol eta can accurately replicate CPDs in most cases^37^. The C>T mutations may occur due to a high increase in cytosine deamination (generating uridine) within the CPDs, before the delayed lesion replication in the TLS deficient XP-V cells.

Most XP-V skin tumors also carry C>A transversions, although this type of mutation was not the most frequent. C>A mutations are commonly associated with oxidized bases, particularly 8-oxodG lesions. These lesions are frequently induced on DNA by UVA-induced oxidation and may pair with adenine, which after the second round of replication may result in G:C to T:A (or C>A) mutations. Pol eta is known to replicate 8-oxodG and, in its absence, other polymerases may be more error-prone^38,39^. C>A mutations are also detected in UVA-irradiated XP-V cells^11^, agreeing with the findings in XPV skin tumors. However, eight skin tumors show a mutation pattern not observed in the irradiated cells, clearly disclosing a CA motif. This motif may reflect a novel mutational signature specific for skin tumors deficient in Pol eta and may result from a general oxidative stress^40^ (and the generation of oxidized bases) induced after sunlight irradiation.

Curiously, four of the eleven skin tumors did not show a typical sunlight-induced mutation signature, indicating a different mutation pattern. A substantial contribution to the mutation signature of tobacco chewing was detected in these four tumors (and not in the other seven). One of the patients (XP85GO.br) recognized he had this habit during his whole life and clinically his mouth and jaw were severely affected with many surgeries for removing tumor lesions. Two of his skin tumors (two melanomas from his back) studied could thus have a contribution of this habit in their formation. For patients XP11GO.br and XP45GO.br, the correlation with tobacco chewing is less clear, although patient XP45GO.br recognized she had smoked for a period of her life. The SCC from patient XP11GO.br shows a different mutation pattern. This may be because the tumor was related to the patient clinical history, as she reports to have been exposed to a tumor chemotherapy treatment for skin cancer before 1970. Although this chemotherapy may have contributed to the mutations either in the saliva DNA or in the skin tumor, we could not identify the chemotherapy type the patient was submitted to.

The different mutation patterns in these four tumors indicate that XP-V patients are also more sensitive to other DNA damaging agents, not only sunlight, which can also trigger tumorigenic processes. Tobacco chewing and smoking habits are strong candidates for the cause of these tumors, and XP-V patients should be advised to avoid these habits. At least three of these four tumors (except XP11GO) also carry the dipyrimidine CC>TT tandem mutations, so the contribution of sunlight-induced damage to these tumors’ formation cannot be discarded.

As Pol eta functions basically during the replication process, the mutations were investigated considering their positions in the replication forks to check any bias when the putative lesions were replicated in the leading or lagging strands. The most common mutation types, C>T and C>A, did not show any significant bias. These results are curious, and one could expect some bias for C>T mutations, most likely due to the wrong replication of pyrimidine dimers. CPDs are preferentially repaired in the transcribed strand of active genes, and gene transcription usually is co-directional with the direction of fork movement^41^. However, no difference was observed in the number of these mutations replicating in both strands. Similarly, T>G mutations did not show a significant bias in replicating any of the strands. However, although not frequent in the skin tumors investigated, C>G, T>A and T>C mutations revealed significantly increased levels of mutations when the pyrimidine is replicated in the lagging strand. The sequence context of these mutations analysis indicated that the replication of pyrimidine dimers in both leading and lagging strand could be responsible for their formation. Interestingly, for the T mutations (T>A and T>C), a T at the 5’ of the mutated base suggests that specific TT dimers could be replicated, in the absence of Pol eta, in an error-prone mode. One possibility is that specific types of lesions (such as 6-4PP) may lead to the formation of gaps during the replication, and this could be replicated by specific TLS mutator polymerases, like Polymerase zeta, as shown previously^42^.

The profile of insertions (polymorphic and somatic) of mobile elements (MEIs) and retroCNVs was also investigated in these XP-V individuals and into their skin tumors. We found an expected number of polymorphic events into these patients of MEIs and retroCNVs. On the other hand, a surprisingly high number of somatic insertions of MEI, majorly *Alu* elements and L1, and retroCNVs, which was superior to the number of insertions found in a randomly set of skin cancer melanoma from TCGA. Interestingly, while this cohort of XP-V patients presents roughly 40 putative somatic events per skin tumor, skin cancers (melanoma) from TCGA present a lower number of somatic events (less than ten events per tumor). These results indicate that Pol eta protects the human genome also by preventing uncontrolled retrotransposon insertions. Pieces of evidence for the induction of retrotransposition after DNA damage induction or defective DNA repair were previously observed in yeast^43,44^. More recently, retrotransposition in yeast has been associated with aging and genomic instability^45,46^. Moreover, the activation of retrotransposons in mammalian cells has been reported during neurogenesis^47^ and aging^48^.

However, little is known about transposition activation due to DNA damage response and the consequences of this process in human cells. Since Pol eta is a TLS polymerase that also functions in other DNA/RNA processing pathways^49^, we speculate that its deficiency may result in more insertion of elements using an RNA intermediate, such as *Alu*, L1, SVA and retrocopies. The mechanisms leading Pol eta protecting the genome from retransposition are still obscure. Several noncanonical roles of Pol eta may participate in these mechanisms. For example, *bonafide* Pol eta also participates in the stability of fragile sites by preventing under-replicated DNA synthesis during mitosis^50^, which may be an entry point of new MEIs and retroCNVs insertions^51^. Pol eta has also been shown to have a reverse transcriptase activity^52^ and participates in homologous recombination^53^. A more trivial mechanism would rely on the canonical TLS function of Pol eta. The formation of single-stranded DNA gaps in cells replicating DNA after lesion induction, in the absence of TLS polymerases, has been demonstrated^42^. These single-stranded DNA regions could also arise during replication of endogenous damage, and their formation and persistence could provide substrates for retrotransposon insertions. Although these are not well-known activities for Pol eta, the insertions of mobile elements and retroCNVs observed in the XP-V tumors stimulate further research to test these hypotheses.

Among the XP-V tumors, more than 5,000 genes were found carrying mutations that would affect protein function. They are highly variable among the tumors, but a few of those genes contain many mutations, despite their small size. For example, genes *ZNF7* and *CDC27* are mutated in most tumors and have previously been reported as potential tumor gene drivers^25–27^. Their functions, either in transcription regulation (*ZNF7*) and cell cycle (*CDC27*), also call attention to possible involvement in the tumorigenic phenotype of these cells. Other four genes have mutations in most tumors and should also be looked at carefully, as possibly related to tumor phenotypes, such as *IGSF3, PABPC3*, and *TEKT4*. Although these genes are not included in the COSMIC Cancer Gene Census^32^, the results obtained in this work reinforce the possibility that they are not merely passenger genes in skin tumors and may represent novel driver genes.

Moreover, 289 genes carrying mutations were classified as cancer genes in the COSMIC Cancer Gene Census^32^. Interestingly, approximately similar frequencies of these genes are identified as oncogenes or tumor suppressor genes, and only a few (14%) are described in skin tumors from DNA repair proficient patients (including melanoma). The mutation burden in XP-V tumors is commonly high, so their involvement in tumor formation needs confirmation.

The main pathways affected by mutations were identified considering the 301 genes containing five or more mutations. Curiously, extracellular matrix-related pathways were found in most functions detected in KEGG and GO databases. These pathways are the major structural components of the tumor microenvironment, related to cell remodeling and signaling. Therefore, dysregulation by these mutations may contribute to tumor formation and to cancer stemness^54,55^.

Most of the XP-V skin tumors have, as expected, a high frequency of mutations. A high TMB is, *per se*, an emerging biomarker of sensitivity to immune checkpoint inhibitors, with accumulating evidence of more significant benefit of immunotherapy in tumors with high TMB^14,56^. Therefore, based on recent findings’ assumption that TMB is an important biomarker for a good outcome in cancer patients^57^, we strongly argue that immune checkpoint inhibitors might benefit to XP patients. These findings provide scientific evidence for clinicians to decide on the use of immunotherapies for the treatment or prophylaxis of XP-V patients in a more personalized, advanced manner.

Finally, although most (seven) of the tumors carry mutations that demonstrate the impact of sunlight-induced DNA damage in the XP-V patients, some tumors show that other types and patterns of mutations may also occur in the tumors. These data contribute to the understanding of Pol eta’s role in maintaining genomic stability and reveal novel environmental agents that may cause mutations in the patients’ cells. Thus, novel concerns and medical pieces of advice should alert XP-V patients to avoid tobacco chewing and smoking. Finally, tumors with a high mutation burden may respond better to immunotherapies to treat cancers, which may explain results indicating a rapid and long responses of immunotherapy in XP-reported treatment cases^58,59^, giving hope to these patients in controlling these cancers.

## Methods

### Samples, genomic DNA extractions, exome capture and sequencing

This study was approved by the Ethics Committee for Human Research of the Institute of Biomedical Sciences, University of São Paulo (SP, Brazil; protocols #1007 CEP, OF. CEPSH 045-13, and CAAE: 48347515.3.0000.5467). The clinical characteristics of those patients were previously described^9^, and all samples were obtained with the signed informed consent form from six individuals (XP03GO.br, XP06GO.br, XP11GO.br, XP45GO.br, XP52GO.br, XP85GO.br). The samples included eleven skin tumors and saliva samples (control-germline DNA), from the six patients. The tissue samples were also subjected to histopathologic analysis. Eight tumors had a clinical diagnosis of basal cell carcinoma, two others were melanomas, and only one was squamous cell carcinoma (Table 4).

**Table 4:** Main features of the XP-V patients and the samples (saliva and tumors).

| Patient ID | Gender (F/M) | Smoker (Y/N) | *POLH* mutation | Sample ID | Sample Type | Sample Origin | HPE | Age at surgery (y) |
| --- | --- | --- | --- | --- | --- | --- | --- | --- |
| XP03GO.br | F | N | intron6/<br>exon8 (het.) | nT-XP03GO | nonTumor | Saliva | - | 35 |
|  |  |  |  | T-XP03GO | Tumor | Neck | BCC | 37 |
| XP06GO.br | M | N | intron 6<br>(hom.) | nT-XP06GO | nonTumor | Saliva | - | 12 |
|  |  |  |  | T-XP06GO_1 | Tumor | Nose | BCC | 13 |
|  |  |  |  | T-XP06GO_2 | Tumor | Nape | BCC | 13 |
|  |  |  |  | T-XP06GO_3 | Tumor | Nose | BCC | 15 |
|  |  |  |  | T-XP06GO_4 | Tumor | Jaw | BCC | 15 |
| XP11GO.br | F | Y | intron 6<br>(hom.) | nT-XP11GO | nonTumor | Saliva | - | 64 |
|  |  |  |  | T-XP11GO | Tumor | Ankle | SCC | 66 |
| XP45GO.br | F | Y | intron 6<br>(hom.) | nT-XP45GO | nonTumor | Saliva | - | 46 |
|  |  |  |  | T-XP45GO | Tumor | Upper Lip | BCC | 40 |
| XP52GO.br | M | Y | intron 6<br>(hom.) | nT-XP52GO | nonTumor | Saliva | - | 44 |
|  |  |  |  | T-XP52GO_1 | Tumor | Arm | BCC | 46 |
|  |  |  |  | T-XP52GO_2 | Tumor | Arm | BCC | 48 |
| XP85GO.br* | M | Y | intron 6<br>(hom.) | nT-XP85GO | nonTumor | Saliva | - | 77 |
|  |  |  |  | T-XP85GO_1 | Tumor | Back | Mel | 79 |
|  |  |  |  | T-XP85GO_2 | Tumor | Back | Mel | 79 |

\*The patient deceased a while after the melanoma onset. Smoker: Y (yes) or N (no). Het. heterozygous or hom. homozygous for the *POLH* mutations. BCC, Basal Cell Carcinoma, SCC, Squamous Cell Carcinoma and Mel, Melanoma; HPE, Histopathology Exam.

All tumor samples were collected with PAXgene Tissue Container (Qiagen GmbH, Hilden, Germany) and the saliva samples, with Oragene^®^ saliva collectors (DNA Genotek^®^, Ottawa, Canada). For the saliva’s DNA extraction, the Qiagen Genomic DNA kit was used, and for the tumors’ DNA, QIAmp^®^ Fast DNA Tissue Kit. For the whole-exome sequence (WES), the genomic DNA library preparation and the exome enrichment were processed with 5 µg of DNA using the Illumina Nextera^®^ Rapid Capture Exome Enrichment Kit (Illumina, San Diego, CA). Libraries were sequenced in paired-end mode (2×175 bp). Quality control of libraries were performed through the High Sensitivity DNA kit (Agilent Technologies, Santa Clara, CA). The main characteristics for the WES generated (such as the exome sequencing platform, coverage and number of bases read) for saliva and tumor samples are presented in Supplementary Table 1.

### Calling single nucleotide variations and small Insertions and deletions

The FASTQ file for each sample was mapped against the human reference genome (GRCh37) using the BWA-MEM aligner, (version 0.7.17, default parameters)^60^. Potential PCR duplicated reads were removed using Picard (version 2.9.2; default parameters; http://broadinstitute.github.io/picard). Single Nucleotide Variations (SNVs) and Insertions and Deletions (INDELs) calling were performed using the Genomic Analysis Toolkit (GATK) HaplotyperCaller^61^ (version 3.7.0) and variations were annotated using ANNOVAR^62^. Variants with poor mapping quality (QC < 30) were filtered out. For each patient, variants shared between their germline (saliva) and tumor samples were excluded from the set of somatic mutations. Additionally, variants with minor allele frequency (MAF) < 0.001 in genome aggregation database (gnomAD^63^) and in the Brazilian genomic variants (ABraOM^64^) database was used to classify variants as rare.

### Calling novel Mobile Elements and retroCNVs insertions

To investigate novel mobile elements and retroCNVs insertions, we used two different tools: Scramble^65^ and sideRETRO^66^. The first one is a mobile element insertion (MEI) detection tool that identifies three classes of MEIs (insertion of L1, *Alu* and SVA) using WXS data. Briefly, Scramble collects and clusterizes a specific type of alignments (soft clipped reads) from a BAM file. After, it re-aligns these reads to a set of mobile element sequences (e.g., L1Ta, *Alu*Ya5, and SVA-E, for humans) which are, in potential, still active mobile elements. At the end, Scramble gives as output MEI calls in a text file format. Our results were obtained using Scramble’s default parameters. Next, to investigate novel retrocopy insertions (retroCNVs), we use sideRETRO. sideRETRO also uses a specific set of alignments (discordant alignments and supplementary alignments) which are clustered and processed using an unsupervised machine learning method. As a result, sideRETRO generates a text format file that contains features related to each retroCNVs (e.g., their parental gene name, genomic position, strandness, genomic context, and genotyping). The results were obtained using sideRETRO’s default parameters. A somatic event was defined as a retroCNVs or MEI occurring uniquely in a tumor. MEIs or retroCNVs shared by two or more individuals or found in public datasets were defined as polymorphic/germinative.

### Point mutations, mutational spectra, and mutational signature analyses

Unique point mutations, simple motif sequences, and exploratory analyses were obtained using WOLAND (https://github.com/tiagoantonio/woland). The mutational signatures and somatic spectra of the 11 tumors were analyzed using the SomaticSignatures^67^, which explore the SNV frequencies together with the adjacent 3’ and 5’ bases^68^ with a non-negative matrix factorization (NMF) algorithm^69^. Mutational mechanisms can be characterized by the frequency of tri-nucleotides motifs, and the frequency of the 96 possible motifs across all samples defines the mutational spectrum^68^.The sequence motif was built with probability logo (pLogo) (https://plogo.uconn.edu/) using exome sequence as background and removing duplicate from both foreground and background sequences. pLogo plots the motif visualization with residues’ height scaled relative to their statistical significance^15^.All identified single and double base substitutions may leave a characteristic mark on the genome, a pattern of somatic mutation. MutationalPatterns^17^ package (v. 3.4.1) provides an efficient method to determine the contribution of known mutational signatures in the tumor’s data and evaluate the activity of signatures in individual tumors. For this purpose, we used the mutational signatures from Catalogue Of Somatic Mutations In Cancer (COSMIC) (https://cancer.sanger.ac.uk/signatures/), release v93 version 3.2 - March 2021^16^.

### Identification of altered pathways of mutated genes

All mutated genes obtained from the 11 tumor samples were annotated and identified. Silent mutations were not considered for these analyses. Genes with five or more mutations for all tumors were selected for function annotation search using the Database for Annotation, Visualization and Integrated Discovery (DAVID) Bioinformatics Resources (2021 Update) (https://david.ncifcrf.gov/)^33^. The search focused on Kyoto Encyclopedia of Genes and Genomes pathway database (https://www.genome.jp/kegg/)^34^ and Gene Ontology Biological Process (BP), Cellular Component (CC) and Molecular Function (MF) (http://geneontology.org/)^35^. The whole list of mutated genes was compared to COSMIC Cancer Gene Census (tier 1 and 2) for cancer genes^32^.

### Statistical Analysis

Significant nucleotide enrichment for sequence motifs was evaluated and marked as horizontal red lines above and below the x axis and corresponds to a log-odd threshold to achieve a p-value of 0.05 after Bonferroni-corrected statistical significance value^70^.

### Data availability

To analyze point mutation patterns, we used WOLAND, available at https://github.com/tiagoantonio/woland. All sequencing data that support the findings of this study have been deposited in the National Center for Biotechnology Information Sequence Read Archive (SRA) under SRA accession number PRJNA852428. All other relevant data are available from the corresponding author on request.

## Data Availability

All data produced in the present study are available upon reasonable request to the authors.

## Acknowledgements

This work was supported by FAPESP (Sao Paulo Research Foundation, SP, Brazil) [grants #2019/19435-3, #2013/08028-1 and #2017/24418-5] under the International Collaboration Research from FAPESP and The Netherlands Organization for Scientific Research (NWO, The Netherlands); Conselho Nacional de Desenvolvimento Científico e Tecnológico (CNPq, Brasília, DF, Brazil, grant #308868/2018-8); Coordenação de Aperfeiçoamento de Pessoal de Nível Superior (CAPES, Brasília, DF, Brazil, financial code 001). This work was also supported by a grant from L’Oréal Research & Innovation. We are grateful to the support with NGS at the core Facility for Scientific Research – USP (CEFAP-USP/GENIAL), and Multi-user genomic Section of the Human Genome & Stem Cell Research Center (HUG-CELL).

## Author contributions

C.C., C.F.M.M. and J.B.V. wrote the paper. C.F.M.M., J.B.V and R.D.V. conceived the study. J.B.V., L.P.C., V.M., S.C.W.C. and C.F.M.M. contributed to biological material acquisition. M.N.S. and P.S. performed histopathological exams. C.C, V.C.B., T.A.S., P.A.F.G., F.O., J.L.B., R.D.V. and J.W.O. analyzed the data. A.S. and C.F.M.M. supervised the research.

## Materials & Correspondence

Correspondence and material requests should be addressed to.

