## Supplementary material for "Mutational signatures and increased retrotransposon insertions in Xeroderma Pigmentosum variant skin tumors": Suplementary_data

Camila Corradi, Juliana Brandstetter Vilar, Vanessa Candiotti Buzatto, Tiago Antonio de Souza, Ligia Pereira Castro, Veridiana Munford, Rodrigo De Vecchi, Pedro Alexandre Favoretto Galante, Fernanda Orpinelli, José Leonel Buzzo, Mirian Nacagami Sotto, Paulo Saldiva, Jocelânio Wesley de Oliveira, Sulamita Costa Wirth Chaibub, Alain Sarasin, Carlos Frederico Martins Menck

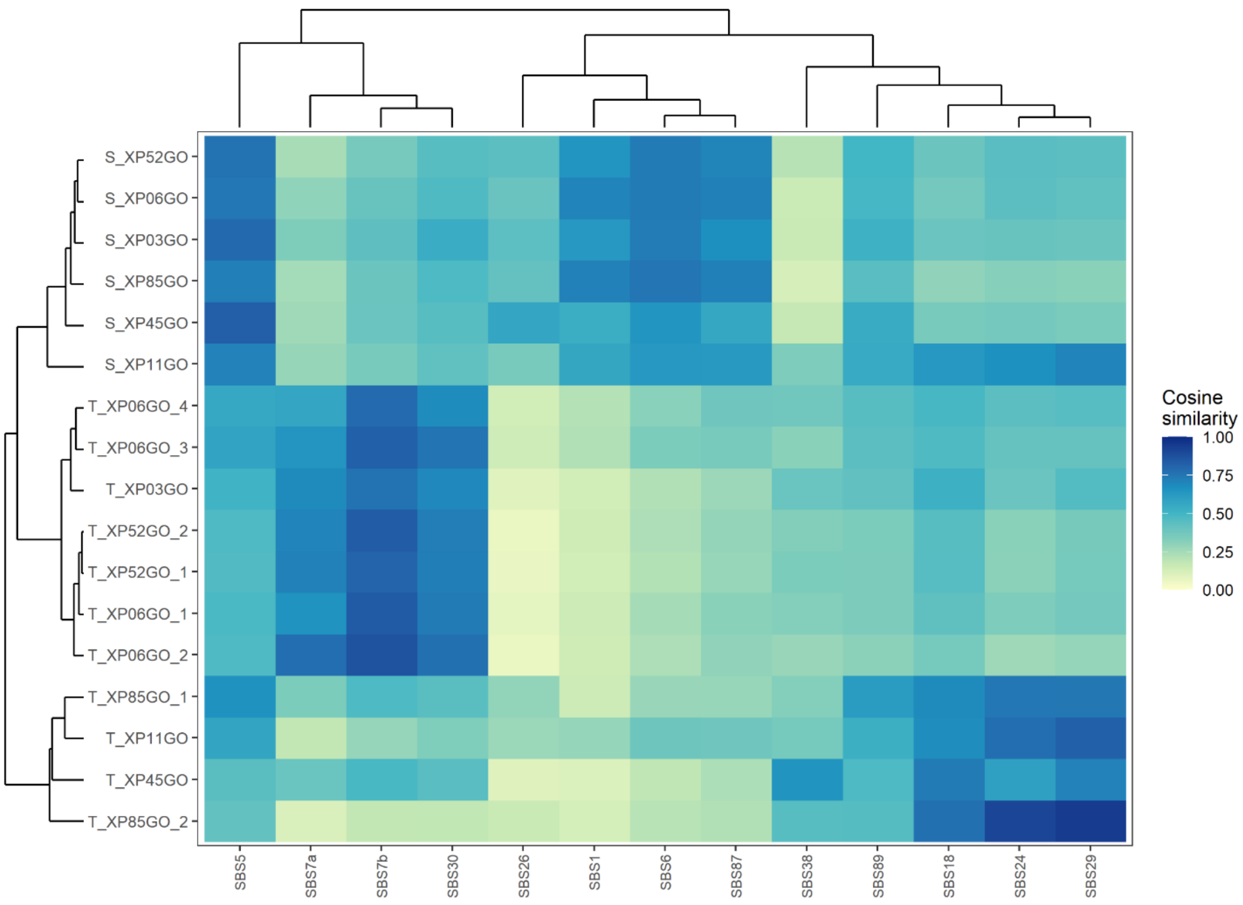


**Supplementary Figure 1:** Similarity between mutational profile of tumor and saliva samples and significant COSMIC mutational signatures (v 3.2) detected. Samples and signatures were clustered based on their cosine similarity by MutationalPatterns.

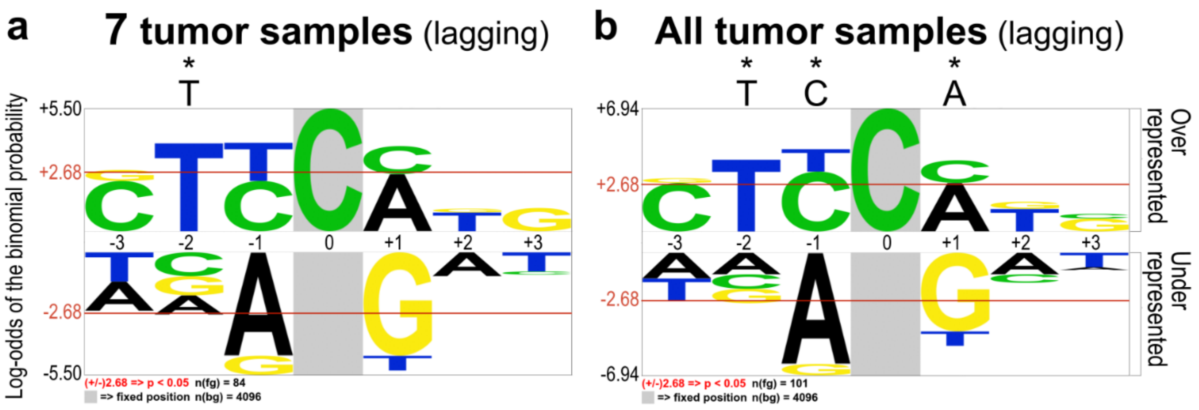


**Supplementary Figure 2:** Sequence context for C>G point mutations detected in the DNA lagging strand of (A) seven XP-V tumor samples (XP03GO, XP06GO_1, XP06GO_2, XP06GO_3, XP06GO_4, XP52GO_1, XP52GO_2) and (B) all tumor samples analyzed together. Probability Logo Generator (pLogo, v1.2.0) was addressed to examine the sequence context adjacent to C>G mutations (highlighted in grey background).

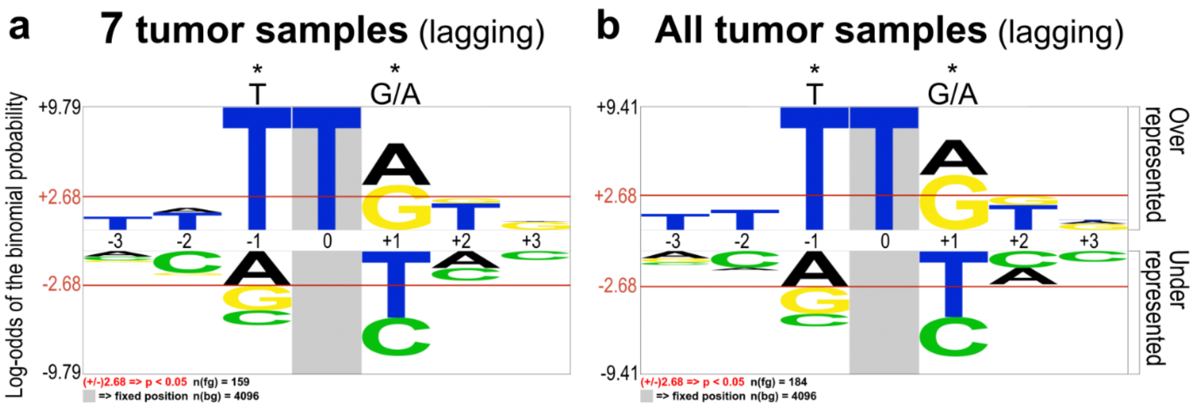


**Supplementary Figure 3:** Sequence context for T>A point mutations detected in the DNA lagging strand of (A) seven XP-V tumor samples (XP03GO, XP06GO_1, XP06GO_2, XP06GO_3, XP06GO_4, XP52GO_1, XP52GO_2) and (B) all tumor samples analyzed together. Probability Logo Generator (pLogo, v1.2.0) was addressed to examine the sequence context adjacent to T>A mutations (highlighted in grey background).

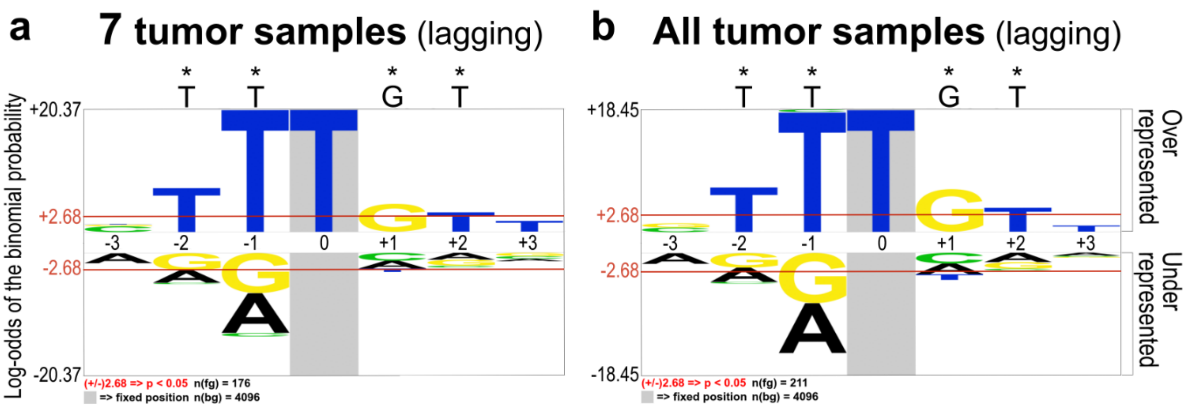


**Supplementary Figure 4:** Sequence context for T>C point mutations detected in the DNA lagging strand of (A) seven XP-V tumor samples (XP03GO, XP06GO_1, XP06GO_2, XP06GO_3, XP06GO_4, XP52GO_1, XP52GO_2) and (B) all tumor samples analyzed together. Probability Logo Generator (pLogo, v1.2.0) was addressed to examine the sequence context adjacent to T>C mutations (highlighted in grey background).

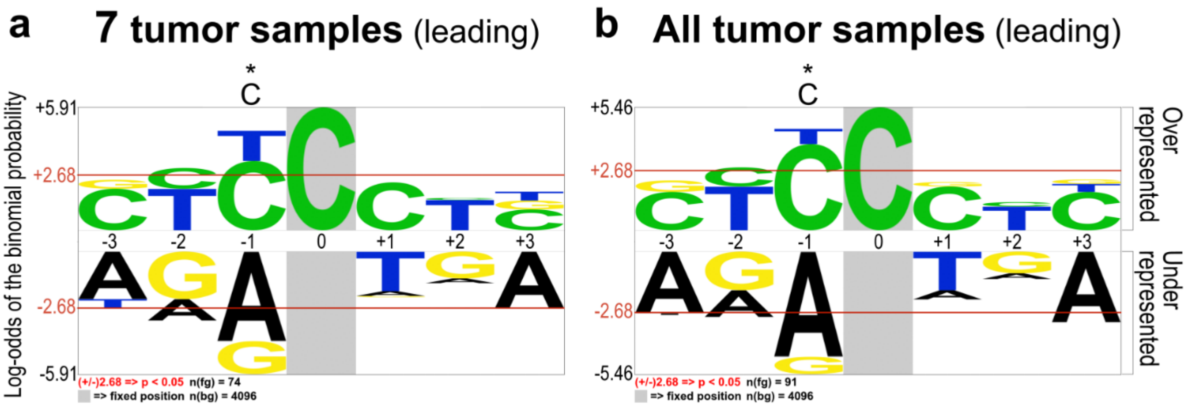


**Supplementary Figure 5:** Sequence context for C>G point mutations detected in the DNA leading strand of (A) seven XP-V tumor samples (XP03GO, XP06GO_1, XP06GO_2, XP06GO_3, XP06GO_4, XP52GO_1, XP52GO_2) and (B) all tumor samples analyzed together. Probability Logo Generator (pLogo, v1.2.0) was addressed to examine the sequence context adjacent to C>G mutations (highlighted in grey background).

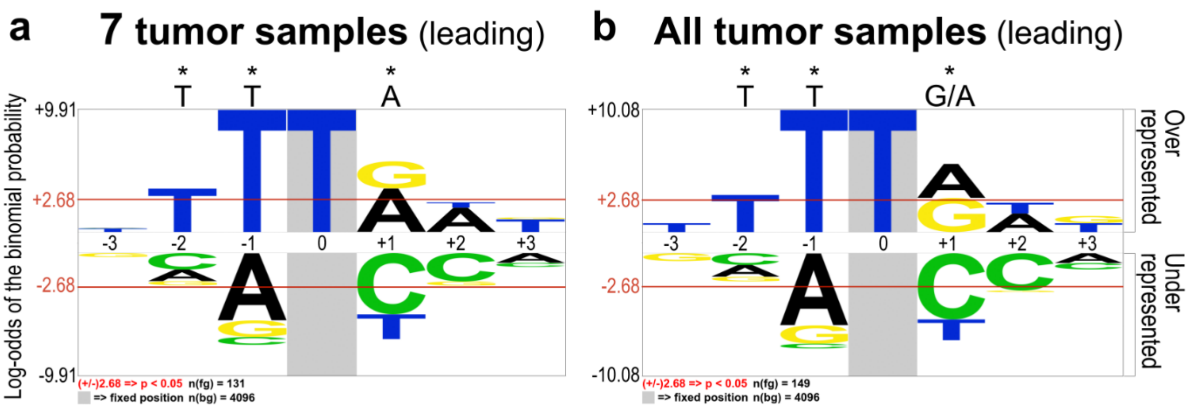


**Supplementary Figure 6:** Sequence context for T>A point mutations detected in the DNA leading strand of (A) seven XP-V tumor samples (XP03GO, XP06GO_1, XP06GO_2, XP06GO_3, XP06GO_4, XP52GO_1, XP52GO_2) and (B) all tumor samples analyzed together. Probability Logo Generator (pLogo, v1.2.0) was addressed to examine the sequence context adjacent to T>A mutations (highlighted in grey background).

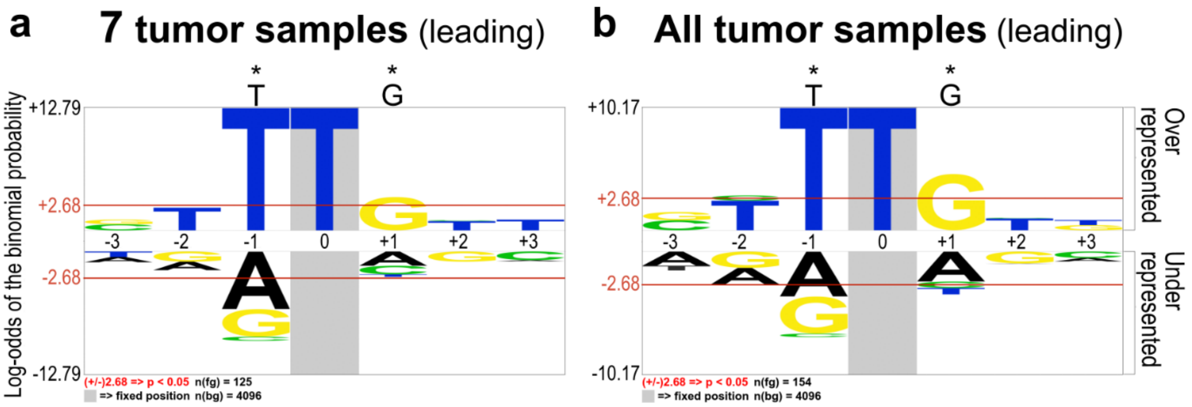


**Supplementary Figure 7:** Sequence context for T>C point mutations detected in the DNA leading strand of (A) seven XP-V tumor samples (XP03GO, XP06GO_1, XP06GO_2, XP06GO_3, XP06GO_4, XP52GO_1, XP52GO_2) and (B) all tumor samples analyzed together. Probability Logo Generator (pLogo, v1.2.0) was addressed to examine the sequence context adjacent to T>C mutations (highlighted in grey background).

**Supplementary Table 1**: Whole exome sequencing platform, depth of coverage and bases read for each tumor and saliva sample.

| Group | Sample | Sequencing platform | Count | | Depth of coverage (%) | | | | | | |
|---|---|---|---|---|---|---|---|---|---|---|---|
| | | | Total bases | Not covered | 1x | 2x | 30x | 50x | 100x | 150x | 200x |
| Saliva | XP03GO | HiSeq 1500 | 41257559 | 351 | 100 | 100 | 86 | 77 | 56 | 39 | 26 |
| | XP06GO | HiSeq 1500 | 41827367 | 392 | 100 | 100 | 87 | 80 | 63 | 48 | 35 |
| | XP11GO | HiSeq 1500 | 40344874 | 381 | 100 | 100 | 74 | 61 | 36 | 20 | 11 |
| | XP45GO | NovaSeq 6000 | 41535164 | 235 | 100 | 100 | 95 | 87 | 44 | 12 | 3 |
| | XP52GO | HiSeq 1500 | 41254872 | 376 | 100 | 100 | 82 | 72 | 50 | 34 | 22 |
| | XP85GO | HiSeq 1500 | 44013054 | 380 | 100 | 100 | 84 | 70 | 39 | 20 | 10 |
| Tumor | XP03GO | NextSeq 550 | 44349745 | 289 | 100 | 100 | 94 | 89 | 70 | 50 | 35 |
| | XP06GO_1 | NextSeq 550 | 44144904 | 340 | 100 | 100 | 86 | 73 | 43 | 23 | 13 |
| | XP06GO_2 | NextSeq 550 | 44035313 | 288 | 100 | 100 | 87 | 76 | 50 | 31 | 18 |
| | XP06GO_3 | NextSeq 550 | 44331540 | 312 | 100 | 100 | 84 | 66 | 29 | 12 | 5 |
| | XP06GO_4 | NextSeq 550 | 44375242 | 209 | 100 | 100 | 91 | 82 | 53 | 31 | 17 |
| | XP11GO | NextSeq 550 | 43891564 | 390 | 100 | 100 | 80 | 64 | 32 | 15 | 8 |
| | XP45GO | NextSeq 550 | 44228848 | 292 | 100 | 100 | 83 | 66 | 30 | 12 | 5 |
| | XP52GO_1 | NextSeq 550 | 44268006 | 227 | 100 | 100 | 91 | 83 | 58 | 37 | 24 |
| | XP52GO_2 | NextSeq 550 | 43797408 | 449 | 100 | 100 | 81 | 68 | 37 | 19 | 10 |
| | XP85GO_1 | NextSeq 550 | 44395507 | 298 | 100 | 100 | 92 | 83 | 57 | 37 | 24 |
| | XP85GO_2 | NextSeq 550 | 44129038 | 406 | 100 | 100 | 87 | 75 | 47 | 27 | 15 |

**Supplementary Table 2**: Point mutation types C>G, T>A, T>C and T>G for each tumor sample.

| Sample | C>G (%) | T>A (%) | T>C (%) | T>G (%) |
|---|---|---|---|---|
| XP03GO | 38 (3) | 68 (5) | 69 (5) | 35 (3) |
| XP06GO_1 | 49 (2) | 72 (3) | 125 (5) | 37 (1) |
| XP06GO_2 | 85 (1) | 162 (3) | 198 (3) | 83 (1) |
| XP06GO_3 | 24 (4) | 17 (3) | 47 (9) | 22 (4) |
| XP06GO_4 | 28 (5) | 27 (5) | 43 (8) | 26 (5) |
| XP11GO* | 26 (6) | 18 (4) | 57 (14) | 17 (4) |
| XP45GO* | 41 (2) | 73 (4) | 96 (6) | 35 (2) |
| XP52GO_1 | 67 (2) | 159 (4) | 154 (4) | 53 (1) |
| XP52GO_2 | 49 (2) | 94 (3) | 95 (4) | 49 (2) |
| XP85GO_1* | 24 (7) | 12 (3) | 54 (16) | 32 (9) |
| XP85GO_2* | 26 (4) | 15 (2) | 75 (10) | 19 (3) |

* These tumor samples presented a higher proportion of T>C than the other tumor samples.

**Supplementary Table 3**: Total count of double mutations for each tumor sample and its frequency compared to total mutations found.

| Group | Sample | Tandem mutations (count) | % In total mutations |
|---|---|---|---|
| Tumor | XP03GO | 77 | 6 |
| | XP06GO_1 | 270 | 10 |
| | XP06GO_2 | 592 | 9 |
| | XP06GO_3 | 61 | 10 |
| | XP06GO_4 | 74 | 13 |
| | *XP11GO* | 9 | 2 |
| | *XP45GO* | 97 | 5 |
| | XP52GO_1 | 309 | 8 |
| | XP52GO_2 | 308 | 11 |
| | *XP85GO_1* | 16 | 4 |
| | *XP85GO_2* | 17 | 2 |

**Supplementary Table 4**: Range of INDEL size from 1 to 76 bp in length and total count for each tumor sample.

This table is large and presented in a separated file.

**Supplementary Table 5**: Polymorphic insertions of Mobile Elements and retroCNVs in XP-V individuals. Sequencing data from normal and tumor samples were used in combination to identify these events.

This table is large and presented in a separated file.

**Supplementary Table 6**: Somatic insertions of Mobile Elements and retroCNVs in tumor samples from XP-V patients.

This table is large and presented in a separated file.

**Supplementary Table 7**: RetroCNVs events into genic (exonic or intronic) positions.

This table is large and presented in a separated file.

**Supplementary Table 8**: Somatic insertions of Mobile Elements and retroCNVs in Skin Cancer Melanoma genomes from The Cancer Genome Atlas.

| Sample | Chr | Start | Class | Putative somatic |
|---|---|---|---|---|
| SKCM-1 | chr5 | 36680619 | MEI-Alu | yes |
| SKCM-2 | chr10 | 104057456 | MEI-Alu | yes |
| SKCM-3 | chr20 | 35174669 | MEI-SVA | yes |
| SKCM-4 | chr21 | 40080138 | MEI-Alu | yes |
| SKCM-6 | chr12 | 110490083 | MEI-Alu | yes |
| SKCM-5 | chr11 | 74522364 | retroCNV | yes |
| SKCM-5 | chr22 | 29767465 | retroCNV | yes |
| SKCM-7 | - | - | - | - |
| SKCM-8 | - | - | - | - |
| SKCM-9 | - | - | - | - |
| SKCM-10 | - | - | - | - |

**Supplementary Table 9**: List of 301 genes carrying four or more mutations that may affect protein function detected in samples from XP-V skin tumors.

This table is large and presented in a separated file.

**Supplementary Table 10**: List of 289 genes carrying mutations in the samples from XP-V skin tumors which were identified as Cancer Genes in the COSMIC Cancer Gene Census.

This table is large and presented in a separated file.

**Supplementary Table 11**: Main pathways identified using DAVID platform in the KEGG database (p-value <10E-05).

| Category | Term | Count | % | p-value |
|---|---|---|---|---|
| KEGG_PATHWAY | Protein digestion and absorption | 17 | 5.6 | 2.7E-11 |
| KEGG_PATHWAY | ECM-receptor interaction | 13 | 4.3 | 4.1E-8 |
| KEGG_PATHWAY | Calcium signaling pathway | 17 | 5.6 | 5.1E-6 |
| KEGG_PATHWAY | Focal adhesion | 14 | 4.7 | 5.7E-5 |

**Supplementary Table 12:** The protein digestion and absorption pathway were identified as the most significant pathways, considering the mutated genes, with 17 genes involved.

| **Protein digestion and absorption pathway** | | |
|---|---|---|
| **ID** | **Gene symbol** | **Gene name** |
| 477 | ATP1A2 | ATPase Na+/K+ transporting subunit alpha 2 |
| 1278 | COL1A2 | collagen type I alpha 2 chain |
| 1280 | COL2A1 | collagen type II alpha 1 chain |
| 1281 | COL3A1 | collagen type III alpha 1 chain |
| 1285 | COL4A3 | collagen type IV alpha 3 chain |
| 1286 | COL4A4 | collagen type IV alpha 4 chain |
| 1287 | COL4A5 | collagen type IV alpha 5 chain |
| 1293 | COL6A3 | collagen type VI alpha 3 chain |
| 256076 | COL6A5 | collagen type VI alpha 5 chain |
| 131873 | COL6A6 | collagen type VI alpha 6 chain |
| 1302 | COL11A2 | collagen type XI alpha 2 chain |
| 1310 | COL19A1 | collagen type XIX alpha 1 chain |
| 1306 | COL15A1 | collagen type XV alpha 1 chain |
| 169044 | COL22A1 | collagen type XXII alpha 1 chain |
| 255631 | COL24A1 | collagen type XXIV alpha 1 chain |
| 6546 | SLC8A1 | solute carrier family 8 member A1 |
| 6547 | SLC8A3 | solute carrier family 8 member A3 |

**Supplementary Table 13**: The extracellular matrix (ECM) receptor interaction pathway was also identified among the most significant ones, with 13 genes mutated.

| ECM-receptor interaction pathway | | |
|---|---|---|
| **ID** | **Gene symbol** | **Gene name** |
| 1278 | COL1A2 | collagen type I alpha 2 chain |
| 1280 | COL2A1 | collagen type II alpha 1 chain |
| 1285 | COL4A3 | collagen type IV alpha 3 chain |
| 1286 | COL4A4 | collagen type IV alpha 4 chain |
| 1287 | COL4A5 | collagen type IV alpha 5 chain |
| 1293 | COL6A3 | collagen type VI alpha 3 chain |
| 256076 | COL6A5 | collagen type VI alpha 5 chain |
| 131873 | COL6A6 | collagen type VI alpha 6 chain |
| 2335 | FN1 | fibronectin 1 |
| 3339 | HSPG2 | heparan sulfate proteoglycan 2 |
| 8516 | ITGA8 | integrin subunit alpha 8 |
| 284217 | LAMA1 | laminin subunit alpha 1 |
| 5649 | RELN | reelin |

**Supplementary Table 14**: The main biological processes detected among mutated genes obtained from GOTERM_BP_DIRECT (p-value <10E-05).

| Category | Term | Count | % | p-value |
|---|---|---|---|---|
| GOTERM_BP_DIRECT | extracellular matrix organization | 28 | 9.3 | 9.4E-16 |
| GOTERM_BP_DIRECT | collagen fibril organization | 14 | 4.7 | 3.6E-9 |
| GOTERM_BP_DIRECT | sensory perception of sound | 16 | 5.3 | 6.5E-9 |
| GOTERM_BP_DIRECT | homophilic cell adhesion via plasma membrane adhesion molecules | 15 | 5.0 | 1.7E-7 |
| GOTERM_BP_DIRECT | cell adhesion | 26 | 8.6 | 4.4E-7 |
| GOTERM_BP_DIRECT | membrane depolarization during action potential | 6 | 2.0 | 2.3E-6 |
| GOTERM_BP_DIRECT | microtubule-based movement | 10 | 3.3 | 4.4E-6 |
| GOTERM_BP_DIRECT | sensory perception of light stimulus | 5 | 1.7 | 5.0E-6 |
| GOTERM_BP_DIRECT | sodium ion transmembrane transport | 10 | 3.3 | 1.4E-5 |
| GOTERM_BP_DIRECT | visual perception | 13 | 4.3 | 4.5E-5 |
| GOTERM_BP_DIRECT | neuronal action potential | 6 | 2.0 | 6.3E-5 |

**Supplementary Table 15**: Mutated genes (28) detected in the extracellular matrix organization in biological process.

| Extracellular matrix organization | | |
|---|---|---|
| **ID** | **Gene symbol** | **Gene name** |
| 81792 | ADAMTS12 | ADAM metallopeptidase with thrombospondin type 1 motif 12 |
| 140766 | ADAMTS14 | ADAM metallopeptidase with thrombospondin type 1 motif 14 |
| 92949 | ADAMTSL1 | ADAMTS like 1 |
| 1278 | COL1A2 | collagen type I alpha 2 chain |
| 1280 | COL2A1 | collagen type II alpha 1 chain |
| 1281 | COL3A1 | collagen type III alpha 1 chain |
| 1285 | COL4A3 | collagen type IV alpha 3 chain |
| 1286 | COL4A4 | collagen type IV alpha 4 chain |
| 1287 | COL4A5 | collagen type IV alpha 5 chain |
| 1293 | COL6A3 | collagen type VI alpha 3 chain |
| 256076 | COL6A5 | collagen type VI alpha 5 chain |
| 131873 | COL6A6 | collagen type VI alpha 6 chain |
| 1302 | COL11A2 | collagen type XI alpha 2 chain |
| 1310 | COL19A1 | collagen type XIX alpha 1 chain |
| 1306 | COL15A1 | collagen type XV alpha 1 chain |
| 169044 | COL22A1 | collagen type XXII alpha 1 chain |
| 255631 | COL24A1 | collagen type XXIV alpha 1 chain |
| 2200 | FBN1 | fibrillin 1 |
| 2243 | FGA | fibrinogen alpha chain |
| 2335 | FN1 | fibronectin 1 |
| 3339 | HSPG2 | heparan sulfate proteoglycan 2 |
| 8516 | ITGA8 | integrin subunit alpha 8 |
| 3681 | ITGAD | integrin subunit alpha D |
| 50939 | IMPG2 | interphotoreceptor matrix proteoglycan 2 |
| 3791 | KDR | kinase insert domain receptor |
| 284217 | LAMA1 | laminin subunit alpha 1 |
| 131149 | OTOL1 | otolin 1 |
| 64116 | SLC39A8 | solute carrier family 39 member 8 |

**Supplementary Table 16**: Mutated genes (14) detected in the collagen fibril organization in biological process.

| Collagen fibril organization | | |
|---|---|---|
| **ID** | **Gene symbol** | **Gene name** |
| 81792 | ADAMTS12 | ADAM metallopeptidase with thrombospondin type 1 motif 12 |
| 140766 | ADAMTS14 | ADAM metallopeptidase with thrombospondin type 1 motif 14 |
| 1278 | COL1A2 | collagen type I alpha 2 chain |
| 1280 | COL2A1 | collagen type II alpha 1 chain |
| 1281 | COL3A1 | collagen type III alpha 1 chain |
| 1285 | COL4A3 | collagen type IV alpha 3 chain |
| 1286 | COL4A4 | collagen type IV alpha 4 chain |
| 1287 | COL4A5 | collagen type IV alpha 5 chain |
| 1293 | COL6A3 | collagen type VI alpha 3 chain |
| 1302 | COL11A2 | collagen type XI alpha 2 chain |
| 1310 | COL19A1 | collagen type XIX alpha 1 chain |
| 1306 | COL15A1 | collagen type XV alpha 1 chain |
| 169044 | COL22A1 | collagen type XXII alpha 1 chain |
| 255631 | COL24A1 | collagen type XXIV alpha 1 chain |

**Supplementary Table 17**: The main molecular functions detected among mutated genes obtained from GOTERM_MF_DIRECT (p-value <10E-05).

| Category | Term | Count | % | p-value |
|---|---|---|---|---|
| GOTERM_MF_DIRECT | extracellular matrix structural constituent | 22 | 7.3 | 4.3E-16 |
| GOTERM_MF_DIRECT | calcium ion binding | 44 | 14.6 | 2.4E-15 |
| GOTERM_MF_DIRECT | extracellular matrix structural constituent conferring tensile strength | 13 | 4.3 | 2.6E-13 |
| GOTERM_MF_DIRECT | ATP-dependent microtubule motor activity, minus-end-directed | 8 | 2.7 | 2.8E-9 |
| GOTERM_MF_DIRECT | dynein light intermediate chain binding | 8 | 2.7 | 7.0E-8 |
| GOTERM_MF_DIRECT | dynein intermediate chain binding | 8 | 2.7 | 5.9E-7 |
| GOTERM_MF_DIRECT | calmodulin binding | 15 | 5.0 | 1.4E-6 |
| GOTERM_MF_DIRECT | cation channel activity | 8 | 2.7 | 1.5E-6 |
| GOTERM_MF_DIRECT | ATP binding | 45 | 15.0 | 4.8E-6 |
| GOTERM_MF_DIRECT | voltage-gated sodium channel activity | 6 | 2.0 | 1.4E-5 |
| GOTERM_MF_DIRECT | actin filament binding | 13 | 4.3 | 6.9E-5 |
| GOTERM_MF_DIRECT | transmembrane receptor protein tyrosine phosphatase activity | 5 | 1.7 | 7.9E-5 |

**Supplementary Table 18**: Mutated genes (22) detected in the extracellular matrix structural constituent molecular function.

| Extracellular matrix structural constituent | | |
|---|---|---|
| **ID** | **Gene symbol** | **Gene name** |
| 1278 | COL1A2 | collagen type I alpha 2 chain |
| 1280 | COL2A1 | collagen type II alpha 1 chain |
| 1281 | COL3A1 | collagen type III alpha 1 chain |
| 1285 | COL4A3 | collagen type IV alpha 3 chain |
| 1286 | COL4A4 | collagen type IV alpha 4 chain |
| 1287 | COL4A5 | collagen type IV alpha 5 chain |
| 1302 | COL11A2 | collagen type XI alpha 2 chain |
| 1310 | COL19A1 | collagen type XIX alpha 1 chain |
| 1306 | COL15A1 | collagen type XV alpha 1 chain |
| 169044 | COL22A1 | collagen type XXII alpha 1 chain |
| 255631 | COL24A1 | collagen type XXIV alpha 1 chain |
| 2200 | FBN1 | fibrillin 1 |
| 2243 | FGA | fibrinogen alpha chain |
| 2335 | FN1 | fibronectin 1 |
| 83872 | HMCN1 | hemicentin 1 |
| 50939 | IMPG2 | interphotoreceptor matrix proteoglycan 2 |
| 284217 | LAMA1 | laminin subunit alpha 1 |
| 4052 | LTBP1 | latent transforming growth factor beta binding protein 1 |
| 727897 | MUC5B | mucin 5B, oligomeric mucus/gel-forming |
| 4588 | MUC6 | mucin 6, oligomeric mucus/gel-forming |
| 22915 | MMRN1 | multimerin 1 |
| 131149 | OTOL1 | otolin 1 |

**Supplementary Table 19**: Mutated genes (44) detected in the calcium ion binding molecular function.

| Calcium ion binding | | |
|---|---|---|
| **ID** | **Gene symbol** | **Gene name** |
| 491 | ATP2B2 | ATPase plasma membrane Ca2+ transporting 2 |
| 2195 | FAT1 | FAT atypical cadherin 1 |
| 120114 | FAT3 | FAT atypical cadherin 3 |
| 79633 | FAT4 | FAT atypical cadherin 4 |
| 53353 | LRP1B | LDL receptor related protein 1B |
| 4036 | LRP2 | LDL receptor related protein 2 |
| 340527 | NHSL2 | NHS like 2 |
| 84059 | ADGRV1 | adhesion G protein-coupled receptor V1 |
| 1008 | CDH10 | cadherin 10 |
| 1010 | CDH12 | cadherin 12 |
| 64072 | CDH23 | cadherin related 23 |
| 773 | CACNA1A | calcium voltage-gated channel subunit alpha1 A |
| 8029 | CUBN | cubilin |
| 54798 | DCHS2 | dachsous cadherin-related 2 |
| 56171 | DNAH7 | dynein axonemal heavy chain 7 |
| 346007 | EYS | eyes shut homolog |
| 2200 | FBN1 | fibrillin 1 |
| 84075 | FSCB | fibrous sheath CABYR binding protein |
| 57493 | HEG1 | heart development protein with EGF like domains 1 |
| 83872 | HMCN1 | hemicentin 1 |
| 3339 | HSPG2 | heparan sulfate proteoglycan 2 |
| 388697 | HRNR | hornerin |
| 4052 | LTBP1 | latent transforming growth factor beta binding protein 1 |
| 23499 | MACF1 | microtubule actin crosslinking factor 1 |
| 22915 | MMRN1 | multimerin 1 |
| 9378 | NRXN1 | neurexin 1 |
| 9381 | OTOF | otoferlin |
| 131149 | OTOL1 | otolin 1 |
| 27445 | PCLO | piccolo presynaptic cytomatrix protein |
| 56126 | PCDHB10 | protocadherin beta 10 |
| 56131 | PCDHB4 | protocadherin beta 4 |
| 56112 | PCDHGA3 | protocadherin gamma subfamily A, 3 |
| 65217 | PCDH15 | protocadherin related 15 |
| 5979 | RET | ret proto-oncogene |
| 6261 | RYR1 | ryanodine receptor 1 |
| 6262 | RYR2 | ryanodine receptor 2 |
| 6263 | RYR3 | ryanodine receptor 3 |
| 9353 | SLIT2 | slit guidance ligand 2 |
| 6546 | SLC8A1 | solute carrier family 8 member A1 |
| 6708 | SPTA1 | spectrin alpha, erythrocytic 1 |
| 79987 | SVEP1 | sushi, von Willebrand factor type A, EGF and pentraxin domain containing 1 |
| 7273 | TTN | titin |
| 440279 | UNC13C | unc-13 homolog C |
| 221806 | VWDE | von Willebrand factor D and EGF domains |

**Supplementary Table 20:** The main cellular components detected among mutated genes obtained from GOTERM_CC_DIRECT (p-value <10E-05).

| Category | Term | Count | % | p-value |
|---|---|---|---|---|
| GOTERM_CC_DIRECT | extracellular matrix | 28 | 9.3 | 4.3E-16 |
| GOTERM_CC_DIRECT | plasma membrane | 123 | 40.9 | 1.2E-11 |
| GOTERM_CC_DIRECT | axon | 25 | 8.3 | 2.0E-10 |
| GOTERM_CC_DIRECT | Z disc | 16 | 5.3 | 5.7E-10 |
| GOTERM_CC_DIRECT | collagen trimer | 14 | 4.7 | 6.8E-10 |
| GOTERM_CC_DIRECT | extracellular region | 65 | 21.6 | 3.4E-9 |
| GOTERM_CC_DIRECT | endoplasmic reticulum lumen | 20 | 6.6 | 9.1E-8 |
| GOTERM_CC_DIRECT | dynein complex | 8 | 2.7 | 9.8E-8 |
| GOTERM_CC_DIRECT | basement membrane | 11 | 3.7 | 8.1E-7 |
| GOTERM_CC_DIRECT | integral component of plasma membrane | 45 | 15.0 | 1.3E-6 |
| GOTERM_CC_DIRECT | apical plasma membrane | 20 | 6.6 | 1.5E-6 |
| GOTERM_CC_DIRECT | sarcolemma | 11 | 3.7 | 2.1E-6 |
| GOTERM_CC_DIRECT | voltage-gated sodium channel complex | 6 | 2.0 | 3.0E-6 |
| GOTERM_CC_DIRECT | integral component of membrane | 112 | 37.2 | 5.4E-6 |
| GOTERM_CC_DIRECT | synapse | 22 | 7.3 | 6.9E-6 |
| GOTERM_CC_DIRECT | T-tubule | 7 | 2.3 | 5.7E-5 |
| GOTERM_CC_DIRECT | receptor complex | 13 | 4.3 | 6.4E-5 |
| GOTERM_CC_DIRECT | glutamatergic synapse | 17 | 5.6 | 7.5E-5 |
| GOTERM_CC_DIRECT | postsynaptic membrane | 11 | 3.7 | 8.9E-5 |
| GOTERM_CC_DIRECT | collagen type IV trimer | 4 | 1.3 | 9.5E-5 |

**Supplementary Table 21**: Mutated genes (28) detected in the extracellular matrix in cellular components.

| Extracellular matrix | | |
|---|---|---|
| **ID** | **Gene symbol** | **Gene name** |
| 81792 | ADAMTS12 | ADAM metallopeptidase with thrombospondin type 1 motif 12 |
| 140766 | ADAMTS14 | ADAM metallopeptidase with thrombospondin type 1 motif 14 |
| 92949 | ADAMTSL1 | ADAMTS like 1 |
| 1278 | COL1A2 | collagen type I alpha 2 chain |
| 1280 | COL2A1 | collagen type II alpha 1 chain |
| 1281 | COL3A1 | collagen type III alpha 1 chain |
| 1285 | COL4A3 | collagen type IV alpha 3 chain |
| 1286 | COL4A4 | collagen type IV alpha 4 chain |
| 1287 | COL4A5 | collagen type IV alpha 5 chain |
| 1293 | COL6A3 | collagen type VI alpha 3 chain |
| 131873 | COL6A6 | collagen type VI alpha 6 chain |
| 1302 | COL11A2 | collagen type XI alpha 2 chain |
| 1310 | COL19A1 | collagen type XIX alpha 1 chain |
| 1306 | COL15A1 | collagen type XV alpha 1 chain |
| 169044 | COL22A1 | collagen type XXII alpha 1 chain |
| 255631 | COL24A1 | collagen type XXIV alpha 1 chain |
| 1755 | DMBT1 | deleted in malignant brain tumors 1 |
| 2200 | FBN1 | fibrillin 1 |
| 2335 | FN1 | fibronectin 1 |
| 50939 | IMPG2 | interphotoreceptor matrix proteoglycan 2 |
| 284217 | LAMA1 | laminin subunit alpha 1 |
| 4052 | LTBP1 | latent transforming growth factor beta binding protein 1 |
| 4585 | MUC4 | mucin 4, cell surface associated |
| 727897 | MUC5B | mucin 5B, oligomeric mucus/gel-forming |
| 4588 | MUC6 | mucin 6, oligomeric mucus/gel-forming |
| 340990 | OTOG | otogelin |
| 131149 | OTOL1 | otolin 1 |
| 5649 | RELN | reelin |

**Supplementary Table 22**: Mutated genes (123) detected in the plasma membrane in cellular components.

This table is large and presented in a separated file.
