## Supplementary material for "Mutational signatures and increased retrotransposon insertions in Xeroderma Pigmentosum variant skin tumors": Suplementary_tables

**Supplementary Table 4**

| INDEL size (bp) | Tumor | | | | | | | | | | | |
|---|---|---|---|---|---|---|---|---|---|---|---|---|
| | XP03GO | XP06GO_1 | XP06GO_2 | XP06GO_3 | XP06GO_4 | XP52GO_1 | XP52GO_2 | XP11GO | XP45GO | XP85GO_1 | XP85GO_2 | Total |
| **1** | 31 | 73 | 111 | 29 | 26 | 65 | 44 | 53 | 68 | 36 | 74 | 610 |
| **2** | 7 | 15 | 14 | 6 | 5 | 7 | 20 | 6 | 12 | 3 | 9 | 104 |
| **3** | 5 | 5 | 12 | 4 | 2 | 4 | 9 | 3 | 6 | 3 | 6 | 59 |
| **4** | 5 | 3 | 5 | 1 | 4 | | 5 | 2 | 3 | 3 | 4 | 35 |
| **5** | 1 | 1 | 2 | 1 | 1 | | 1 | 1 | 1 | 1 | | 10 |
| **6** | 1 | | 3 | | | 1 | 2 | | | | 3 | 10 |
| **7** | 1 | 1 | 1 | 1 | 1 | 1 | 1 | 2 | | | 1 | 10 |
| **8** | | | | | | | 1 | 1 | | | 2 | 4 |
| **10** | 1 | 1 | 1 | 2 | 2 | 1 | 2 | 2 | 2 | | 1 | 15 |
| **12** | 1 | | 2 | | | | | | | 1 | 1 | 5 |
| **13** | | 3 | 2 | 1 | 1 | | | | | 1 | | 8 |
| **15** | 1 | | | | | | | | | | | 1 |
| **16** | | 2 | | 1 | | | | | | | | 3 |
| **17** | | | 1 | | | | 1 | | | | | 2 |
| **18** | 1 | | | | 1 | | | | | | | 2 |
| **21** | | | | | | | | | | 1 | 1 | 2 |
| **22** | | | | | | | 1 | | | | | 1 |
| **23** | | | | | | | | 1 | | | | 1 |
| **24** | | | 1 | | | | | | | 1 | 1 | 3 |
| **25** | | 1 | 1 | 1 | | | | | | 2 | 1 | 6 |
| **28** | 1 | | | | | | | | 1 | | | 2 |
| **31** | 1 | | | | | | | | | | | 1 |
| **34** | | | | | | | | 1 | | | | 1 |
| **35** | | 1 | | | | | | | | | | 1 |
| **39** | | | 1 | | | | | | | | | 1 |
| **40** | | 1 | | | | | | | | | | 1 |
| **43** | | | | | | | | 1 | | | | 1 |

| | | | | | | | | | | | | |
|---|---|---|---|---|---|---|---|---|---|---|---|---|
| **45** | | | | | | | | 1 | | | | 1 |
| **46** | | | 1 | | | | | | | | | 1 |
| **47** | | | 1 | | | | | | | | | 1 |
| **51** | | | | 1 | | | 1 | 1 | | | | 3 |
| **53** | | | | | 1 | | | | | | | 1 |
| **55** | | | | | | | | 1 | | | | 1 |
| **60** | | | | | 1 | | | | | | | 1 |
| **70** | | 1 | | 1 | 1 | | | | | | 1 | 4 |
| **73** | | | | | | | | | | 1 | | 1 |
| **74** | | | | | 1 | | | | | | | 1 |
| **76** | | | | | | | | | | 1 | | 1 |
| **Total** | 57 | 108 | 159 | 49 | 47 | 79 | 88 | 76 | 93 | 54 | 105 | 915 |

| Supplementary Table 5 | | | | |
|---|---|---|---|---|
| **Sample** | **Chr** | **Start** | **Class** | **Polimorphic** |
| XP03GO | chr1 | 20618244 | MEI-Alu | yes |
| XP03GO | chr15 | 76800144 | MEI-Alu | yes |
| XP03GO | chr18 | 9401405 | MEI-Alu | yes |
| XP03GO | chr4 | 186172333 | MEI-Alu | yes |
| XP03GO | chr8 | 41946763 | MEI-Alu | yes |
| XP03GO | chr1 | 21424735 | MEI-Alu | yes |
| XP03GO | chr1 | 92701962 | MEI-Alu | yes |
| XP03GO | chr11 | 106299754 | MEI-Alu | yes |
| XP03GO | chr11 | 428014 | MEI-Alu | yes |
| XP03GO | chr13 | 21320625 | MEI-Alu | yes |
| XP03GO | chr14 | 44544896 | MEI-Alu | yes |
| XP03GO | chr17 | 32572168 | MEI-Alu | yes |
| XP03GO | chr18 | 76241969 | MEI-Alu | yes |
| XP03GO | chr19 | 52384817 | MEI-Alu | yes |
| XP03GO | chr4 | 178277157 | MEI-Alu | yes |
| XP03GO | chr4 | 185440770 | MEI-Alu | yes |
| XP03GO | chr4 | 185460992 | MEI-Alu | yes |
| XP03GO | chr5 | 170774335 | MEI-Alu | yes |
| XP03GO | chr5 | 73037329 | MEI-Alu | yes |
| XP03GO | chr5 | 95658505 | MEI-Alu | yes |
| XP03GO | chr7 | 38169597 | MEI-Alu | yes |
| XP06GO | chr1 | 92701962 | MEI-Alu | yes |
| XP06GO | chr10 | 6229164 | MEI-Alu | yes |
| XP06GO | chr11 | 104274020 | MEI-Alu | yes |
| XP06GO | chr11 | 428014 | MEI-Alu | yes |
| XP06GO | chr11 | 43855889 | MEI-Alu | yes |
| XP06GO | chr13 | 21320625 | MEI-Alu | yes |
| XP06GO | chr17 | 63488529 | MEI-Alu | yes |
| XP06GO | chr18 | 9401405 | MEI-Alu | yes |
| XP06GO | chr19 | 41807206 | MEI-Alu | yes |
| XP06GO | chr19 | 52384817 | MEI-Alu | yes |
| XP06GO | chr22 | 17224400 | MEI-Alu | yes |
| XP06GO | chr3 | 121885667 | MEI-Alu | yes |
| XP06GO | chr4 | 185440770 | MEI-Alu | yes |
| XP06GO | chr4 | 185460992 | MEI-Alu | yes |
| XP06GO | chr8 | 119788552 | MEI-Alu | yes |
| XP06GO | chrX | 53215274 | MEI-Alu | yes |
| XP06GO | chr13 | 21956223 | MEI-Alu | yes |
| XP06GO | chr2 | 207607677 | MEI-Alu | yes |
| XP06GO | chr15 | 40808120 | MEI-Alu | yes |
| XP06GO | chr20 | 29885844 | MEI-Alu | yes |

| | | | | |
|---|---|---|---|---|
| XP06GO | chr17 | 80082249 | MEI-Alu | yes |
| XP06GO | chr5 | 1812526 | MEI-Alu | yes |
| XP06GO | chr12 | 63236703 | MEI-Alu | yes |
| XP11GO | chr1 | 21424735 | MEI-Alu | yes |
| XP11GO | chr10 | 53046195 | MEI-Alu | yes |
| XP11GO | chr11 | 428014 | MEI-Alu | yes |
| XP11GO | chr13 | 38878403 | MEI-Alu | yes |
| XP11GO | chr14 | 102562102 | MEI-Alu | yes |
| XP11GO | chr15 | 76800144 | MEI-Alu | yes |
| XP11GO | chr15 | 94345318 | MEI-Alu | yes |
| XP11GO | chr17 | 63488529 | MEI-Alu | yes |
| XP11GO | chr19 | 52384817 | MEI-Alu | yes |
| XP11GO | chr4 | 185440770 | MEI-Alu | yes |
| XP11GO | chr4 | 186172333 | MEI-Alu | yes |
| XP11GO | chr5 | 62561291 | MEI-Alu | yes |
| XP11GO | chr5 | 95658505 | MEI-Alu | yes |
| XP11GO | chr7 | 38169597 | MEI-Alu | yes |
| XP11GO | chr9 | 136768496 | MEI-Alu | yes |
| XP45GO | chr1 | 21424735 | MEI-Alu | yes |
| XP45GO | chr11 | 428014 | MEI-Alu | yes |
| XP45GO | chr13 | 21320625 | MEI-Alu | yes |
| XP45GO | chr15 | 94345318 | MEI-Alu | yes |
| XP45GO | chr19 | 52384817 | MEI-Alu | yes |
| XP45GO | chr19 | 9820429 | MEI-Alu | yes |
| XP45GO | chr3 | 121885667 | MEI-Alu | yes |
| XP45GO | chr4 | 185440770 | MEI-Alu | yes |
| XP45GO | chr5 | 62561291 | MEI-Alu | yes |
| XP45GO | chr5 | 95658505 | MEI-Alu | yes |
| XP45GO | chr8 | 41946763 | MEI-Alu | yes |
| XP52GO | chr11 | 428014 | MEI-Alu | yes |
| XP52GO | chr11 | 43855889 | MEI-Alu | yes |
| XP52GO | chr11 | 7695684 | MEI-Alu | yes |
| XP52GO | chr17 | 63488529 | MEI-Alu | yes |
| XP52GO | chr2 | 137502222 | MEI-Alu | yes |
| XP52GO | chr4 | 185440770 | MEI-Alu | yes |
| XP52GO | chr4 | 185460992 | MEI-Alu | yes |
| XP52GO | chr4 | 186172333 | MEI-Alu | yes |
| XP52GO | chr6 | 71544668 | MEI-Alu | yes |
| XP52GO | chr8 | 119788552 | MEI-Alu | yes |
| XP52GO | chr9 | 136768496 | MEI-Alu | yes |
| XP52GO | chr19 | 20563166 | MEI-Alu | yes |
| XP52GO | chr2 | 21573955 | MEI-Alu | yes |
| XP52GO | chr8 | 74463111 | MEI-Alu | yes |
| XP85GO | chr1 | 20618244 | MEI-Alu | yes |

| | | | | |
|---|---|---|---|---|
| XP85GO | chr1 | 92701962 | MEI-Alu | yes |
| XP85GO | chr10 | 53046195 | MEI-Alu | yes |
| XP85GO | chr11 | 428014 | MEI-Alu | yes |
| XP85GO | chr13 | 101737016 | MEI-Alu | yes |
| XP85GO | chr15 | 76800144 | MEI-Alu | yes |
| XP85GO | chr17 | 63488529 | MEI-Alu | yes |
| XP85GO | chr19 | 41807206 | MEI-Alu | yes |
| XP85GO | chr19 | 52384817 | MEI-Alu | yes |
| XP85GO | chr19 | 9820429 | MEI-Alu | yes |
| XP85GO | chr2 | 137502222 | MEI-Alu | yes |
| XP85GO | chr22 | 17224400 | MEI-Alu | yes |
| XP85GO | chr4 | 185440770 | MEI-Alu | yes |
| XP85GO | chr4 | 185460992 | MEI-Alu | yes |
| XP85GO | chr4 | 186172333 | MEI-Alu | yes |
| XP85GO | chr5 | 95658505 | MEI-Alu | yes |
| XP85GO | chr6 | 14133803 | MEI-Alu | yes |
| XP85GO | chr6 | 71544668 | MEI-Alu | yes |
| XP85GO | chr8 | 119788552 | MEI-Alu | yes |
| XP85GO | chr8 | 41946763 | MEI-Alu | yes |
| XP85GO | chr9 | 7852538 | MEI-Alu | yes |
| XP03GO | chr2 | 102706037 | MEI-L1 | yes |
| XP03GO | chr2 | 32916254 | MEI-L1 | yes |
| XP03GO | chr2 | 32916256 | MEI-L1 | yes |
| XP03GO | chr2 | 32916257 | MEI-L1 | yes |
| XP03GO | chr9 | 97913251 | MEI-L1 | yes |
| XP06GO | chr1 | 169555621 | MEI-L1 | yes |
| XP06GO | chr2 | 102706037 | MEI-L1 | yes |
| XP06GO | chr2 | 32916254 | MEI-L1 | yes |
| XP06GO | chr2 | 32916256 | MEI-L1 | yes |
| XP06GO | chr2 | 32916257 | MEI-L1 | yes |
| XP06GO | chr5 | 115057973 | MEI-L1 | yes |
| XP11GO | chr2 | 32916254 | MEI-L1 | yes |
| XP11GO | chr2 | 32916256 | MEI-L1 | yes |
| XP11GO | chr2 | 32916257 | MEI-L1 | yes |
| XP11GO | chr5 | 115057973 | MEI-L1 | yes |
| XP45GO | chr2 | 32916254 | MEI-L1 | yes |
| XP45GO | chr2 | 32916256 | MEI-L1 | yes |
| XP45GO | chr2 | 32916257 | MEI-L1 | yes |
| XP45GO | chr22 | 43928709 | MEI-L1 | yes |
| XP45GO | chr9 | 97913251 | MEI-L1 | yes |
| XP52GO | chr2 | 102706037 | MEI-L1 | yes |
| XP52GO | chr2 | 32916254 | MEI-L1 | yes |
| XP52GO | chr2 | 32916257 | MEI-L1 | yes |
| XP52GO | chr3 | 101712435 | MEI-L1 | yes |

| | | | | |
|---|---|---|---|---|
| XP85GO | chr2 | 102706037 | MEI-L1 | yes |
| XP85GO | chr2 | 32916254 | MEI-L1 | yes |
| XP85GO | chr2 | 32916256 | MEI-L1 | yes |
| XP85GO | chr2 | 32916257 | MEI-L1 | yes |
| XP85GO | chr22 | 43928709 | MEI-L1 | yes |
| XP03GO | chr4 | 9703222 | MEI-SVA | yes |
| XP06GO | chr4 | 9703222 | MEI-SVA | yes |
| XP06GO | chr6 | 43687667 | MEI-SVA | yes |
| XP11GO | chr6 | 43687667 | MEI-SVA | yes |
| XP03GO | chr1 | 109108012 | retroCNV | yes |
| XP03GO | chr1 | 221182809 | retroCNV | yes |
| XP03GO | chr11 | 38791107 | retroCNV | yes |
| XP03GO | chr11 | 58199876 | retroCNV | yes |
| XP03GO | chr12 | 125316601 | retroCNV | yes |
| XP03GO | chr15 | 40561980 | retroCNV | yes |
| XP03GO | chr9 | 33130550 | retroCNV | yes |
| XP06GO | chr1 | 109108012 | retroCNV | yes |
| XP06GO | chr1 | 221182809 | retroCNV | yes |
| XP06GO | chr11 | 27194661 | retroCNV | yes |
| XP06GO | chr11 | 38791107 | retroCNV | yes |
| XP06GO | chr12 | 125316601 | retroCNV | yes |
| XP06GO | chr13 | 40046135 | retroCNV | yes |
| XP06GO | chr15 | 40561980 | retroCNV | yes |
| XP06GO | chr8 | 22443288 | retroCNV | yes |
| XP06GO | chr9 | 33130550 | retroCNV | yes |
| XP11GO | chr1 | 109108012 | retroCNV | yes |
| XP11GO | chr1 | 221182809 | retroCNV | yes |
| XP11GO | chr11 | 27194661 | retroCNV | yes |
| XP11GO | chr11 | 58199876 | retroCNV | yes |
| XP11GO | chr12 | 125316601 | retroCNV | yes |
| XP11GO | chr15 | 40561980 | retroCNV | yes |
| XP11GO | chr8 | 22443288 | retroCNV | yes |
| XP11GO | chr9 | 33130550 | retroCNV | yes |
| XP52GO | chr1 | 221182809 | retroCNV | yes |
| XP52GO | chr11 | 27194661 | retroCNV | yes |
| XP52GO | chr11 | 38791107 | retroCNV | yes |
| XP52GO | chr12 | 125316601 | retroCNV | yes |
| XP52GO | chr15 | 40561980 | retroCNV | yes |
| XP52GO | chr17 | 7885350 | retroCNV | yes |
| XP85GO | chr1 | 221182809 | retroCNV | yes |
| XP85GO | chr12 | 125316601 | retroCNV | yes |
| XP85GO | chr13 | 40046135 | retroCNV | yes |
| XP85GO | chr15 | 40561980 | retroCNV | yes |
| XP85GO | chr9 | 33130550 | retroCNV | yes |

| Supplementary Table 6 | | | | |
|---|---|---|---|---|
| **Sample** | **Chr** | **Start** | **Class** | **Putative somatic** |
| XP03GO | chr1 | 227414577 | MEI-Alu | yes |
| XP03GO | chr12 | 19722736 | MEI-Alu | yes |
| XP03GO | chr13 | 19032019 | MEI-Alu | yes |
| XP03GO | chr13 | 77560960 | MEI-Alu | yes |
| XP03GO | chr14 | 31422743 | MEI-Alu | yes |
| XP03GO | chr14 | 71402515 | MEI-Alu | yes |
| XP03GO | chr15 | 79503530 | MEI-Alu | yes |
| XP03GO | chr16 | 67805140 | MEI-Alu | yes |
| XP03GO | chr17 | 16769811 | MEI-Alu | yes |
| XP03GO | chr17 | 80082253 | MEI-Alu | yes |
| XP03GO | chr19 | 45325154 | MEI-Alu | yes |
| XP03GO | chr2 | 151458477 | MEI-Alu | yes |
| XP03GO | chr2 | 162822107 | MEI-Alu | yes |
| XP03GO | chr2 | 236062557 | MEI-Alu | yes |
| XP03GO | chr20 | 3057234 | MEI-Alu | yes |
| XP03GO | chr3 | 9635210 | MEI-Alu | yes |
| XP03GO | chr4 | 68079061 | MEI-Alu | yes |
| XP03GO | chr4 | 83637030 | MEI-Alu | yes |
| XP03GO | chr5 | 13831015 | MEI-Alu | yes |
| XP03GO | chr5 | 157139729 | MEI-Alu | yes |
| XP03GO | chr5 | 160080694 | MEI-Alu | yes |
| XP03GO | chr6 | 70411459 | MEI-Alu | yes |
| XP03GO | chr8 | 113780679 | MEI-Alu | yes |
| XP03GO | chr8 | 127344421 | MEI-Alu | yes |
| XP03GO | chr9 | 73375741 | MEI-Alu | yes |
| XP03GO | chrX | 45459626 | MEI-Alu | yes |
| XP03GO | chr11 | 108514359 | MEI-L1 | yes |
| XP03GO | chr11 | 134435753 | MEI-L1 | yes |
| XP03GO | chr15 | 55958954 | MEI-L1 | yes |
| XP03GO | chr15 | 87278910 | MEI-L1 | yes |
| XP03GO | chr2 | 15026175 | MEI-L1 | yes |
| XP03GO | chr2 | 32916253 | MEI-L1 | yes |
| XP03GO | chr3 | 186654352 | MEI-L1 | yes |
| XP03GO | chr4 | 116779942 | MEI-L1 | yes |
| XP03GO | chr5 | 110144556 | MEI-L1 | yes |
| XP03GO | chr5 | 32415140 | MEI-L1 | yes |
| XP03GO | chr1 | 191743287 | retroCNV | yes |
| XP03GO | chr11 | 67759312 | retroCNV | yes |
| XP03GO | chr11 | 73391889 | retroCNV | yes |
| XP03GO | chr12 | 5799706 | retroCNV | yes |
| XP03GO | chr17 | 32157815 | retroCNV | yes |
| XP03GO | chr2 | 15965136 | retroCNV | yes |
| XP03GO | chr2 | 96259884 | retroCNV | yes |
| XP03GO | chr2 | 114469868 | retroCNV | yes |

| | | | | |
|---|---|---|---|---|
| XP03GO | chr20 | 8125823 | retroCNV | yes |
| XP03GO | chr3 | 13144967 | retroCNV | yes |
| XP03GO | chr3 | 42201917 | retroCNV | yes |
| XP03GO | chr3 | 142566603 | retroCNV | yes |
| XP03GO | chr4 | 40797843 | retroCNV | yes |
| XP03GO | chr4 | 98517880 | retroCNV | yes |
| XP03GO | chr4 | 188107213 | retroCNV | yes |
| XP03GO | chr5 | 3079846 | retroCNV | yes |
| XP03GO | chr5 | 87281071 | retroCNV | yes |
| XP03GO | chr5 | 142977287 | retroCNV | yes |
| XP03GO | chr7 | 114698620 | retroCNV | yes |
| XP03GO | chr8 | 108118141 | retroCNV | yes |
| XP03GO | chr9 | 123035315 | retroCNV | yes |
| XP03GO | chrX | 49269510 | retroCNV | yes |
| XP06GO_1 | chr1 | 196811798 | MEI-Alu | yes |
| XP06GO_1 | chr1 | 207767565 | MEI-Alu | yes |
| XP06GO_1 | chr1 | 57962641 | MEI-Alu | yes |
| XP06GO_1 | chr10 | 46022526 | MEI-Alu | yes |
| XP06GO_1 | chr10 | 57142454 | MEI-Alu | yes |
| XP06GO_1 | chr10 | 60365100 | MEI-Alu | yes |
| XP06GO_1 | chr11 | 107946134 | MEI-Alu | yes |
| XP06GO_1 | chr12 | 4661708 | MEI-Alu | yes |
| XP06GO_1 | chr12 | 47918707 | MEI-Alu | yes |
| XP06GO_1 | chr16 | 11579499 | MEI-Alu | yes |
| XP06GO_1 | chr17 | 22141618 | MEI-Alu | yes |
| XP06GO_1 | chr4 | 179335800 | MEI-Alu | yes |
| XP06GO_1 | chr4 | 26484866 | MEI-Alu | yes |
| XP06GO_1 | chr6 | 106365679 | MEI-Alu | yes |
| XP06GO_1 | chr6 | 106481273 | MEI-Alu | yes |
| XP06GO_1 | chr7 | 158930169 | MEI-Alu | yes |
| XP06GO_1 | chrM | 10850 | MEI-Alu | yes |
| XP06GO_1 | chrX | 118456970 | MEI-Alu | yes |
| XP06GO_1 | chr1 | 233826463 | MEI-L1 | yes |
| XP06GO_1 | chr10 | 23553112 | MEI-L1 | yes |
| XP06GO_1 | chr13 | 37993308 | MEI-L1 | yes |
| XP06GO_1 | chr17 | 2377533 | MEI-L1 | yes |
| XP06GO_1 | chr2 | 146084981 | MEI-L1 | yes |
| XP06GO_1 | chr3 | 123871879 | MEI-L1 | yes |
| XP06GO_1 | chr4 | 123971835 | MEI-L1 | yes |
| XP06GO_1 | chr4 | 131260492 | MEI-L1 | yes |
| XP06GO_1 | chr5 | 129107059 | MEI-L1 | yes |
| XP06GO_1 | chr10 | 26939156 | MEI-SVA | yes |
| XP06GO_1 | chr11 | 22482602 | retroCNV | yes |
| XP06GO_1 | chr12 | 62302147 | retroCNV | yes |
| XP06GO_1 | chr15 | 98287269 | retroCNV | yes |
| XP06GO_1 | chr17 | 34151550 | retroCNV | yes |
| XP06GO_1 | chr18 | 34352976 | retroCNV | yes |

| | | | | |
|---|---|---|---|---|
| XP06GO_1 | chr21 | 28618935 | retroCNV | yes |
| XP06GO_1 | chr3 | 14842879 | retroCNV | yes |
| XP06GO_1 | chr3 | 180986068 | retroCNV | yes |
| XP06GO_1 | chr3 | 186100783 | retroCNV | yes |
| XP06GO_1 | chr5 | 11629149 | retroCNV | yes |
| XP06GO_1 | chr6 | 15945934 | retroCNV | yes |
| XP06GO_1 | chr6 | 23322335 | retroCNV | yes |
| XP06GO_1 | chr8 | 47677159 | retroCNV | yes |
| XP06GO_1 | chr8 | 101739598 | retroCNV | yes |
| XP06GO_1 | chr8 | 133055875 | retroCNV | yes |
| XP06GO_1 | chr9 | 114930289 | retroCNV | yes |
| XP06GO_2 | chr1 | 212145296 | MEI-Alu | yes |
| XP06GO_2 | chr1 | 247125683 | MEI-Alu | yes |
| XP06GO_2 | chr1 | 39351729 | MEI-Alu | yes |
| XP06GO_2 | chr1 | 70997553 | MEI-Alu | yes |
| XP06GO_2 | chr10 | 123843236 | MEI-Alu | yes |
| XP06GO_2 | chr10 | 43102664 | MEI-Alu | yes |
| XP06GO_2 | chr11 | 61001491 | MEI-Alu | yes |
| XP06GO_2 | chr12 | 57750921 | MEI-Alu | yes |
| XP06GO_2 | chr13 | 37375844 | MEI-Alu | yes |
| XP06GO_2 | chr13 | 41327735 | MEI-Alu | yes |
| XP06GO_2 | chr13 | 78697466 | MEI-Alu | yes |
| XP06GO_2 | chr13 | 81236057 | MEI-Alu | yes |
| XP06GO_2 | chr13 | 99132558 | MEI-Alu | yes |
| XP06GO_2 | chr14 | 57277732 | MEI-Alu | yes |
| XP06GO_2 | chr15 | 42156533 | MEI-Alu | yes |
| XP06GO_2 | chr18 | 47907702 | MEI-Alu | yes |
| XP06GO_2 | chr19 | 10151719 | MEI-Alu | yes |
| XP06GO_2 | chr2 | 217818211 | MEI-Alu | yes |
| XP06GO_2 | chr2 | 230252768 | MEI-Alu | yes |
| XP06GO_2 | chr3 | 156522084 | MEI-Alu | yes |
| XP06GO_2 | chr3 | 185544949 | MEI-Alu | yes |
| XP06GO_2 | chr4 | 105449241 | MEI-Alu | yes |
| XP06GO_2 | chr4 | 110013803 | MEI-Alu | yes |
| XP06GO_2 | chr6 | 5348529 | MEI-Alu | yes |
| XP06GO_2 | chr6 | 79077973 | MEI-Alu | yes |
| XP06GO_2 | chr8 | 101201606 | MEI-Alu | yes |
| XP06GO_2 | chr8 | 134712799 | MEI-Alu | yes |
| XP06GO_2 | chr8 | 22922452 | MEI-Alu | yes |
| XP06GO_2 | chr8 | 41412939 | MEI-Alu | yes |
| XP06GO_2 | chr8 | 60614220 | MEI-Alu | yes |
| XP06GO_2 | chr8 | 92038356 | MEI-Alu | yes |
| XP06GO_2 | chr1 | 42775364 | MEI-L1 | yes |
| XP06GO_2 | chr1 | 75700352 | MEI-L1 | yes |
| XP06GO_2 | chr14 | 36205554 | MEI-L1 | yes |
| XP06GO_2 | chr15 | 100341874 | MEI-L1 | yes |
| XP06GO_2 | chr17 | 30071290 | MEI-L1 | yes |

| | | | | |
|---|---|---|---|---|
| XP06GO_2 | chr4 | 190117296 | MEI-L1 | yes |
| XP06GO_2 | chr4 | 85783942 | MEI-L1 | yes |
| XP06GO_2 | chr1 | 111959485 | retroCNV | yes |
| XP06GO_2 | chr1 | 191608630 | retroCNV | yes |
| XP06GO_2 | chr1 | 206485306 | retroCNV | yes |
| XP06GO_2 | chr1 | 220712244 | retroCNV | yes |
| XP06GO_2 | chr10 | 44510929 | retroCNV | yes |
| XP06GO_2 | chr10 | 117483590 | retroCNV | yes |
| XP06GO_2 | chr12 | 343209 | retroCNV | yes |
| XP06GO_2 | chr12 | 106594694 | retroCNV | yes |
| XP06GO_2 | chr16 | 82286877 | retroCNV | yes |
| XP06GO_2 | chr2 | 23972787 | retroCNV | yes |
| XP06GO_2 | chr2 | 78867933 | retroCNV | yes |
| XP06GO_2 | chr2 | 159858461 | retroCNV | yes |
| XP06GO_2 | chr21 | 29297506 | retroCNV | yes |
| XP06GO_2 | chr3 | 12829439 | retroCNV | yes |
| XP06GO_2 | chr3 | 48683547 | retroCNV | yes |
| XP06GO_2 | chr3 | 73112464 | retroCNV | yes |
| XP06GO_2 | chr4 | 99413345 | retroCNV | yes |
| XP06GO_2 | chr5 | 39115382 | retroCNV | yes |
| XP06GO_2 | chr5 | 140365871 | retroCNV | yes |
| XP06GO_2 | chr5 | 150413520 | retroCNV | yes |
| XP06GO_2 | chr6 | 6193235 | retroCNV | yes |
| XP06GO_2 | chr6 | 108589443 | retroCNV | yes |
| XP06GO_2 | chr7 | 43625598 | retroCNV | yes |
| XP06GO_2 | chr7 | 69225629 | retroCNV | yes |
| XP06GO_2 | chr9 | 3776260 | retroCNV | yes |
| XP06GO_2 | chr9 | 122586748 | retroCNV | yes |
| XP06GO_2 | chr9 | 128635413 | retroCNV | yes |
| XP06GO_3 | chr10 | 69850419 | MEI-Alu | yes |
| XP06GO_3 | chr12 | 117055163 | MEI-Alu | yes |
| XP06GO_3 | chr2 | 9748660 | MEI-Alu | yes |
| XP06GO_3 | chr22 | 26601643 | MEI-Alu | yes |
| XP06GO_3 | chr3 | 172605146 | MEI-Alu | yes |
| XP06GO_3 | chr7 | 125908192 | MEI-Alu | yes |
| XP06GO_3 | chr10 | 68394359 | MEI-L1 | yes |
| XP06GO_3 | chr6 | 77765940 | MEI-L1 | yes |
| XP06GO_3 | chr18 | 11960675 | MEI-SVA | yes |
| XP06GO_3 | chr15 | 93296472 | retroCNV | yes |
| XP06GO_4 | chr1 | 19471896 | MEI-Alu | yes |
| XP06GO_4 | chr10 | 80353984 | MEI-Alu | yes |
| XP06GO_4 | chr11 | 103330538 | MEI-Alu | yes |
| XP06GO_4 | chr11 | 127045430 | MEI-Alu | yes |
| XP06GO_4 | chr18 | 33953227 | MEI-Alu | yes |
| XP06GO_4 | chr2 | 147883186 | MEI-Alu | yes |
| XP06GO_4 | chr3 | 78394509 | MEI-Alu | yes |
| XP06GO_4 | chr5 | 155863614 | MEI-Alu | yes |

| | | | | |
|---|---|---|---|---|
| XP06GO_4 | chr8 | 30984336 | MEI-Alu | yes |
| XP06GO_4 | chr3 | 38584591 | MEI-L1 | yes |
| XP06GO_4 | chr6 | 22147156 | MEI-L1 | yes |
| XP06GO_4 | chr7 | 145560488 | MEI-L1 | yes |
| XP06GO_4 | chr7 | 57668467 | MEI-L1 | yes |
| XP06GO_4 | chr1 | 22729253 | retroCNV | yes |
| XP06GO_4 | chr1 | 201303808 | retroCNV | yes |
| XP06GO_4 | chr12 | 63323494 | retroCNV | yes |
| XP06GO_4 | chr16 | 8930336 | retroCNV | yes |
| XP06GO_4 | chr2 | 223908560 | retroCNV | yes |
| XP11GO | chr1 | 160077875 | MEI-Alu | yes |
| XP11GO | chr1 | 196917129 | MEI-Alu | yes |
| XP11GO | chr1 | 233727113 | MEI-Alu | yes |
| XP11GO | chr11 | 23501258 | MEI-Alu | yes |
| XP11GO | chr11 | 25583592 | MEI-Alu | yes |
| XP11GO | chr11 | 42034362 | MEI-Alu | yes |
| XP11GO | chr11 | 81454388 | MEI-Alu | yes |
| XP11GO | chr12 | 111448186 | MEI-Alu | yes |
| XP11GO | chr12 | 56133724 | MEI-Alu | yes |
| XP11GO | chr13 | 22496055 | MEI-Alu | yes |
| XP11GO | chr14 | 28158712 | MEI-Alu | yes |
| XP11GO | chr14 | 34483263 | MEI-Alu | yes |
| XP11GO | chr14 | 35009343 | MEI-Alu | yes |
| XP11GO | chr15 | 68767800 | MEI-Alu | yes |
| XP11GO | chr16 | 11579494 | MEI-Alu | yes |
| XP11GO | chr16 | 58766988 | MEI-Alu | yes |
| XP11GO | chr16 | 68940586 | MEI-Alu | yes |
| XP11GO | chr3 | 118453221 | MEI-Alu | yes |
| XP11GO | chr3 | 18993960 | MEI-Alu | yes |
| XP11GO | chr4 | 55974806 | MEI-Alu | yes |
| XP11GO | chr5 | 162804931 | MEI-Alu | yes |
| XP11GO | chr5 | 87076878 | MEI-Alu | yes |
| XP11GO | chr6 | 26994392 | MEI-Alu | yes |
| XP11GO | chr8 | 113729336 | MEI-Alu | yes |
| XP11GO | chr8 | 136679302 | MEI-Alu | yes |
| XP11GO | chr8 | 88014563 | MEI-Alu | yes |
| XP11GO | chr9 | 101461593 | MEI-Alu | yes |
| XP11GO | chrX | 3957594 | MEI-Alu | yes |
| XP11GO | chrX | 50634935 | MEI-Alu | yes |
| XP11GO | chr1 | 199646838 | MEI-L1 | yes |
| XP11GO | chr12 | 48324886 | MEI-L1 | yes |
| XP11GO | chr14 | 39010090 | MEI-L1 | yes |
| XP11GO | chr17 | 4735501 | MEI-L1 | yes |
| XP11GO | chr2 | 220639029 | MEI-L1 | yes |
| XP11GO | chr21 | 42407333 | MEI-L1 | yes |
| XP11GO | chr3 | 90872023 | MEI-L1 | yes |
| XP11GO | chr4 | 114211547 | MEI-L1 | yes |

| | | | | |
|---|---|---|---|---|
| XP11GO | chr4 | 132632536 | MEI-L1 | yes |
| XP11GO | chr5 | 92621451 | MEI-L1 | yes |
| XP11GO | chr6 | 147800366 | MEI-L1 | yes |
| XP11GO | chr9 | 112133994 | MEI-L1 | yes |
| XP11GO | chr1 | 94604228 | retroCNV | yes |
| XP11GO | chr1 | 176028861 | retroCNV | yes |
| XP11GO | chr10 | 3520437 | retroCNV | yes |
| XP11GO | chr12 | 29873548 | retroCNV | yes |
| XP11GO | chr13 | 19114615 | retroCNV | yes |
| XP11GO | chr15 | 40735497 | retroCNV | yes |
| XP11GO | chr16 | 57100779 | retroCNV | yes |
| XP11GO | chr16 | 68569361 | retroCNV | yes |
| XP11GO | chr17 | 11328943 | retroCNV | yes |
| XP11GO | chr17 | 81957100 | retroCNV | yes |
| XP11GO | chr18 | 7619420 | retroCNV | yes |
| XP11GO | chr18 | 44573486 | retroCNV | yes |
| XP11GO | chr18 | 66272128 | retroCNV | yes |
| XP11GO | chr18 | 75374358 | retroCNV | yes |
| XP11GO | chr19 | 1581509 | retroCNV | yes |
| XP11GO | chr19 | 36418814 | retroCNV | yes |
| XP11GO | chr19 | 52308189 | retroCNV | yes |
| XP11GO | chr2 | 53324236 | retroCNV | yes |
| XP11GO | chr2 | 65092929 | retroCNV | yes |
| XP11GO | chr22 | 30952155 | retroCNV | yes |
| XP11GO | chr22 | 44438629 | retroCNV | yes |
| XP11GO | chr3 | 13367614 | retroCNV | yes |
| XP11GO | chr4 | 6545462 | retroCNV | yes |
| XP11GO | chr4 | 31026179 | retroCNV | yes |
| XP11GO | chr4 | 44258565 | retroCNV | yes |
| XP11GO | chr4 | 48809166 | retroCNV | yes |
| XP11GO | chr4 | 109820286 | retroCNV | yes |
| XP11GO | chr5 | 1681033 | retroCNV | yes |
| XP11GO | chr6 | 95641643 | retroCNV | yes |
| XP11GO | chr7 | 75951606 | retroCNV | yes |
| XP11GO | chr7 | 106190581 | retroCNV | yes |
| XP11GO | chr8 | 22692988 | retroCNV | yes |
| XP11GO | chr8 | 46996611 | retroCNV | yes |
| XP11GO | chr8 | 83209270 | retroCNV | yes |
| XP11GO | chr8 | 109478786 | retroCNV | yes |
| XP11GO | chr8 | 128099452 | retroCNV | yes |
| XP45GO | chr1 | 16057263 | MEI-Alu | yes |
| XP45GO | chr1 | 27253754 | MEI-Alu | yes |
| XP45GO | chr11 | 133555206 | MEI-Alu | yes |
| XP45GO | chr12 | 83692333 | MEI-Alu | yes |
| XP45GO | chr13 | 26376203 | MEI-Alu | yes |
| XP45GO | chr20 | 16248011 | MEI-Alu | yes |
| XP45GO | chr20 | 55145311 | MEI-Alu | yes |

| | | | | |
|---|---|---|---|---|
| XP45GO | chr21 | 29703769 | MEI-Alu | yes |
| XP45GO | chr3 | 171908583 | MEI-Alu | yes |
| XP45GO | chr3 | 174692946 | MEI-Alu | yes |
| XP45GO | chr3 | 2772357 | MEI-Alu | yes |
| XP45GO | chr5 | 37688709 | MEI-Alu | yes |
| XP45GO | chr7 | 2424371 | MEI-Alu | yes |
| XP45GO | chr7 | 95248186 | MEI-Alu | yes |
| XP45GO | chr8 | 22368514 | MEI-Alu | yes |
| XP45GO | chr9 | 42746579 | MEI-Alu | yes |
| XP45GO | chr9 | 76337149 | MEI-Alu | yes |
| XP45GO | chr11 | 62575356 | MEI-L1 | yes |
| XP45GO | chr11 | 8399234 | MEI-L1 | yes |
| XP45GO | chr12 | 31020266 | MEI-L1 | yes |
| XP45GO | chr4 | 41354432 | MEI-L1 | yes |
| XP45GO | chr19 | 23845119 | MEI-SVA | yes |
| XP45GO | chr1 | 59552137 | retroCNV | yes |
| XP45GO | chr1 | 116536227 | retroCNV | yes |
| XP45GO | chr10 | 64950545 | retroCNV | yes |
| XP45GO | chr11 | 69120088 | retroCNV | yes |
| XP45GO | chr13 | 113727350 | retroCNV | yes |
| XP45GO | chr15 | 78248231 | retroCNV | yes |
| XP45GO | chr18 | 56489936 | retroCNV | yes |
| XP45GO | chr19 | 45976634 | retroCNV | yes |
| XP45GO | chr2 | 38509459 | retroCNV | yes |
| XP45GO | chr2 | 122663903 | retroCNV | yes |
| XP45GO | chr2 | 178495137 | retroCNV | yes |
| XP45GO | chr22 | 32793639 | retroCNV | yes |
| XP45GO | chr3 | 32890868 | retroCNV | yes |
| XP45GO | chr3 | 173636930 | retroCNV | yes |
| XP45GO | chr4 | 168482442 | retroCNV | yes |
| XP45GO | chr4 | 185251360 | retroCNV | yes |
| XP45GO | chr5 | 12935329 | retroCNV | yes |
| XP45GO | chr8 | 123025031 | retroCNV | yes |
| XP45GO | chr8 | 139560859 | retroCNV | yes |
| XP45GO | chr9 | 77651505 | retroCNV | yes |
| XP52GO_1 | chr1 | 70446750 | MEI-Alu | yes |
| XP52GO_1 | chr10 | 95612859 | MEI-Alu | yes |
| XP52GO_1 | chr11 | 41448552 | MEI-Alu | yes |
| XP52GO_1 | chr13 | 87924838 | MEI-Alu | yes |
| XP52GO_1 | chr17 | 48539858 | MEI-Alu | yes |
| XP52GO_1 | chr17 | 74372417 | MEI-Alu | yes |
| XP52GO_1 | chr20 | 54108683 | MEI-Alu | yes |
| XP52GO_1 | chr22 | 19585578 | MEI-Alu | yes |
| XP52GO_1 | chr3 | 96155673 | MEI-Alu | yes |
| XP52GO_1 | chr5 | 109715291 | MEI-Alu | yes |
| XP52GO_1 | chr7 | 124141306 | MEI-Alu | yes |
| XP52GO_1 | chr7 | 127835618 | MEI-Alu | yes |

| | | | | |
|---|---|---|---|---|
| XP52GO_1 | chr8 | 71293822 | MEI-Alu | yes |
| XP52GO_1 | chr8 | 78616457 | MEI-Alu | yes |
| XP52GO_1 | chr9 | 74909421 | MEI-Alu | yes |
| XP52GO_1 | chr1 | 85582466 | MEI-L1 | yes |
| XP52GO_1 | chr10 | 105553984 | MEI-L1 | yes |
| XP52GO_1 | chr12 | 111901216 | MEI-L1 | yes |
| XP52GO_1 | chr18 | 22285088 | MEI-L1 | yes |
| XP52GO_1 | chr2 | 49214691 | MEI-L1 | yes |
| XP52GO_1 | chr3 | 111380393 | MEI-L1 | yes |
| XP52GO_1 | chr8 | 119476661 | MEI-L1 | yes |
| XP52GO_1 | chr8 | 133959999 | MEI-L1 | yes |
| XP52GO_1 | chr9 | 100303735 | MEI-L1 | yes |
| XP52GO_1 | chr9 | 72289997 | MEI-L1 | yes |
| XP52GO_1 | chr10 | 69213563 | MEI-SVA | yes |
| XP52GO_1 | chr12 | 78946590 | MEI-SVA | yes |
| XP52GO_1 | chr18 | 49810945 | MEI-SVA | yes |
| XP52GO_1 | chr9 | 122853996 | MEI-SVA | yes |
| XP52GO_1 | chr1 | 175342356 | retroCNV | yes |
| XP52GO_1 | chr1 | 208311225 | retroCNV | yes |
| XP52GO_1 | chr10 | 45578006 | retroCNV | yes |
| XP52GO_1 | chr11 | 39952346 | retroCNV | yes |
| XP52GO_1 | chr12 | 128466525 | retroCNV | yes |
| XP52GO_1 | chr14 | 48626911 | retroCNV | yes |
| XP52GO_1 | chr14 | 66472071 | retroCNV | yes |
| XP52GO_1 | chr14 | 72143500 | retroCNV | yes |
| XP52GO_1 | chr15 | 33662861 | retroCNV | yes |
| XP52GO_1 | chr17 | 10722502 | retroCNV | yes |
| XP52GO_1 | chr18 | 22088110 | retroCNV | yes |
| XP52GO_1 | chr2 | 31985611 | retroCNV | yes |
| XP52GO_1 | chr2 | 128493720 | retroCNV | yes |
| XP52GO_1 | chr2 | 203522644 | retroCNV | yes |
| XP52GO_1 | chr2 | 210392963 | retroCNV | yes |
| XP52GO_1 | chr20 | 51102655 | retroCNV | yes |
| XP52GO_1 | chr3 | 77983665 | retroCNV | yes |
| XP52GO_1 | chr5 | 86540336 | retroCNV | yes |
| XP52GO_1 | chr5 | 158405809 | retroCNV | yes |
| XP52GO_1 | chr6 | 124364609 | retroCNV | yes |
| XP52GO_1 | chr6 | 167871481 | retroCNV | yes |
| XP52GO_1 | chr8 | 5940020 | retroCNV | yes |
| XP52GO_1 | chr8 | 65871921 | retroCNV | yes |
| XP52GO_1 | chr9 | 107202554 | retroCNV | yes |
| XP52GO_2 | chr10 | 28132927 | MEI-Alu | yes |
| XP52GO_2 | chr10 | 46647423 | MEI-Alu | yes |
| XP52GO_2 | chr12 | 111618446 | MEI-Alu | yes |
| XP52GO_2 | chr16 | 24805704 | MEI-Alu | yes |
| XP52GO_2 | chr17 | 75596573 | MEI-Alu | yes |
| XP52GO_2 | chr6 | 10762465 | MEI-Alu | yes |

| | | | | |
|---|---|---|---|---|
| XP52GO_2 | chr8 | 116034794 | MEI-Alu | yes |
| XP52GO_2 | chr2 | 73503424 | MEI-L1 | yes |
| XP52GO_2 | chr3 | 113410353 | MEI-L1 | yes |
| XP52GO_2 | chr17 | 71139551 | retroCNV | yes |
| XP52GO_2 | chr4 | 123674956 | retroCNV | yes |
| XP52GO_2 | chr7 | 131251214 | retroCNV | yes |
| XP85GO_1 | chr11 | 48911557 | MEI-Alu | yes |
| XP85GO_1 | chr12 | 28136067 | MEI-Alu | yes |
| XP85GO_1 | chr12 | 38984453 | MEI-Alu | yes |
| XP85GO_1 | chr13 | 108133285 | MEI-Alu | yes |
| XP85GO_1 | chr13 | 42688592 | MEI-Alu | yes |
| XP85GO_1 | chr13 | 46521218 | MEI-Alu | yes |
| XP85GO_1 | chr13 | 79624957 | MEI-Alu | yes |
| XP85GO_1 | chr16 | 14405443 | MEI-Alu | yes |
| XP85GO_1 | chr16 | 53841069 | MEI-Alu | yes |
| XP85GO_1 | chr17 | 21814103 | MEI-Alu | yes |
| XP85GO_1 | chr17 | 28869902 | MEI-Alu | yes |
| XP85GO_1 | chr4 | 108659090 | MEI-Alu | yes |
| XP85GO_1 | chr4 | 183463282 | MEI-Alu | yes |
| XP85GO_1 | chr4 | 65764464 | MEI-Alu | yes |
| XP85GO_1 | chr7 | 23626066 | MEI-Alu | yes |
| XP85GO_1 | chr9 | 124044401 | MEI-Alu | yes |
| XP85GO_1 | chr15 | 47215131 | MEI-L1 | yes |
| XP85GO_1 | chr18 | 39874252 | MEI-L1 | yes |
| XP85GO_1 | chr2 | 137488062 | MEI-L1 | yes |
| XP85GO_1 | chr2 | 146263358 | MEI-L1 | yes |
| XP85GO_1 | chr3 | 111556218 | MEI-L1 | yes |
| XP85GO_1 | chr3 | 58278429 | MEI-L1 | yes |
| XP85GO_1 | chr9 | 79861141 | MEI-L1 | yes |
| XP85GO_1 | chr2 | 55676601 | MEI-SVA | yes |
| XP85GO_1 | chr10 | 99365030 | retroCNV | yes |
| XP85GO_1 | chr11 | 74699711 | retroCNV | yes |
| XP85GO_1 | chr14 | 30252861 | retroCNV | yes |
| XP85GO_1 | chr15 | 40258743 | retroCNV | yes |
| XP85GO_1 | chr15 | 55565856 | retroCNV | yes |
| XP85GO_1 | chr16 | 86405752 | retroCNV | yes |
| XP85GO_1 | chr19 | 46607282 | retroCNV | yes |
| XP85GO_1 | chr22 | 44887892 | retroCNV | yes |
| XP85GO_1 | chr3 | 117912232 | retroCNV | yes |
| XP85GO_1 | chr3 | 177037638 | retroCNV | yes |
| XP85GO_1 | chr5 | 99281539 | retroCNV | yes |
| XP85GO_1 | chr6 | 150811672 | retroCNV | yes |
| XP85GO_2 | chr1 | 218429247 | MEI-Alu | yes |
| XP85GO_2 | chr1 | 231796363 | MEI-Alu | yes |
| XP85GO_2 | chr11 | 56535887 | MEI-Alu | yes |
| XP85GO_2 | chr12 | 121806053 | MEI-Alu | yes |
| XP85GO_2 | chr14 | 59987437 | MEI-Alu | yes |

| | | | | |
|---|---|---|---|---|
| XP85GO_2 | chr15 | 39399404 | MEI-Alu | yes |
| XP85GO_2 | chr15 | 40954299 | MEI-Alu | yes |
| XP85GO_2 | chr15 | 78513317 | MEI-Alu | yes |
| XP85GO_2 | chr16 | 18948781 | MEI-Alu | yes |
| XP85GO_2 | chr17 | 56870222 | MEI-Alu | yes |
| XP85GO_2 | chr2 | 113348869 | MEI-Alu | yes |
| XP85GO_2 | chr2 | 114633159 | MEI-Alu | yes |
| XP85GO_2 | chr2 | 22717723 | MEI-Alu | yes |
| XP85GO_2 | chr2 | 54889450 | MEI-Alu | yes |
| XP85GO_2 | chr22 | 19373922 | MEI-Alu | yes |
| XP85GO_2 | chr3 | 122726900 | MEI-Alu | yes |
| XP85GO_2 | chr3 | 57421657 | MEI-Alu | yes |
| XP85GO_2 | chr3 | 99007932 | MEI-Alu | yes |
| XP85GO_2 | chr6 | 151076419 | MEI-Alu | yes |
| XP85GO_2 | chr7 | 1092039 | MEI-Alu | yes |
| XP85GO_2 | chr7 | 78107875 | MEI-Alu | yes |
| XP85GO_2 | chr7 | 98568474 | MEI-Alu | yes |
| XP85GO_2 | chr8 | 61915551 | MEI-Alu | yes |
| XP85GO_2 | chr9 | 26476357 | MEI-Alu | yes |
| XP85GO_2 | chr9 | 82290303 | MEI-Alu | yes |
| XP85GO_2 | chr9 | 99385086 | MEI-Alu | yes |
| XP85GO_2 | chr1 | 201221526 | MEI-L1 | yes |
| XP85GO_2 | chr1 | 75833543 | MEI-L1 | yes |
| XP85GO_2 | chr11 | 28991341 | MEI-L1 | yes |
| XP85GO_2 | chr2 | 15026487 | MEI-L1 | yes |
| XP85GO_2 | chr2 | 186987891 | MEI-L1 | yes |
| XP85GO_2 | chr20 | 22117082 | MEI-L1 | yes |
| XP85GO_2 | chr7 | 108188819 | MEI-L1 | yes |
| XP85GO_2 | chr9 | 27241882 | MEI-L1 | yes |
| XP85GO_2 | chr6 | 153108721 | MEI-SVA | yes |
| XP85GO_2 | chr9 | 93075916 | MEI-SVA | yes |
| XP85GO_2 | chr1 | 114738217 | retroCNV | yes |
| XP85GO_2 | chr1 | 161282685 | retroCNV | yes |
| XP85GO_2 | chr11 | 64599164 | retroCNV | yes |
| XP85GO_2 | chr15 | 96751192 | retroCNV | yes |
| XP85GO_2 | chr16 | 72122927 | retroCNV | yes |
| XP85GO_2 | chr17 | 6108894 | retroCNV | yes |
| XP85GO_2 | chr17 | 57424738 | retroCNV | yes |
| XP85GO_2 | chr18 | 59100745 | retroCNV | yes |
| XP85GO_2 | chr19 | 17555979 | retroCNV | yes |
| XP85GO_2 | chr2 | 45823048 | retroCNV | yes |
| XP85GO_2 | chr2 | 54710268 | retroCNV | yes |
| XP85GO_2 | chr2 | 143674834 | retroCNV | yes |
| XP85GO_2 | chr2 | 159189792 | retroCNV | yes |
| XP85GO_2 | chr22 | 26623361 | retroCNV | yes |
| XP85GO_2 | chr3 | 122149653 | retroCNV | yes |
| XP85GO_2 | chr5 | 87822085 | retroCNV | yes |

| | | | | |
|---|---|---|---|---|
| XP85GO_2 | chr5 | 123001489 | retroCNV | yes |
| XP85GO_2 | chr6 | 22208564 | retroCNV | yes |
| XP85GO_2 | chr7 | 54679749 | retroCNV | yes |
| XP85GO_2 | chr7 | 88366469 | retroCNV | yes |
| XP85GO_2 | chr7 | 96949512 | retroCNV | yes |
| XP85GO_2 | chr7 | 151395256 | retroCNV | yes |
| XP85GO_2 | chr8 | 50082791 | retroCNV | yes |
| XP85GO_2 | chr8 | 62314115 | retroCNV | yes |
| XP85GO_2 | chr9 | 89405550 | retroCNV | yes |
| XP85GO_2 | chr9 | 136483902 | retroCNV | yes |
| XP85GO_2 | chrX | 112618715 | retroCNV | yes |

| Supplementary Table 7 | | | | |
|---|---|---|---|---|
| **Sample** | **Chr** | **Position** | **Gene_host** | **Region** |
| XP03GO | chr9 | 123035315 | RABGAP1 | exonic |
| XP06GO_1 | chr9 | 114930289 | TNFSF8 | exonic |
| XP06GO_2 | chr1 | 206485306 | IKBKE | exonic |
| XP06GO_2 | chr2 | 159858461 | LY75-CD302 | exonic |
| XP06GO_2 | chr5 | 140365871 | SLC4A9 | exonic |
| XP06GO_2 | chr7 | 43625598 | STK17A | exonic |
| XP06GO_2 | chr9 | 128635413 | WDR34 | exonic |
| XP06GO_4 | chr16 | 8930336 | USP7 | exonic |
| XP11GO | chr19 | 1581509 | MBD3 | exonic |
| XP11GO | chr8 | 22692988 | EGR3 | exonic |
| XP45GO | chr1 | 116536227 | CD58 | exonic |
| XP45GO | chr3 | 32890868 | TRIM71 | exonic |
| XP45GO | chr8 | 123025031 | DERL1 | exonic |
| XP52GO_1 | chr15 | 33662861 | RYR3 | exonic |
| XP85GO_2 | chr16 | 72122927 | PMFBP1 | exonic |
| XP85GO_2 | chr9 | 89405550 | SEMA4D | exonic |
| XP03GO | chr11 | 73391889 | RELT | intronic |
| XP03GO | chr12 | 5799706 | ANO2 | intronic |
| XP03GO | chr17 | 32157815 | RHOT1 | intronic |
| XP03GO | chr2 | 96259884 | TMEM127 | intronic |
| XP03GO | chr2 | 114469868 | DPP10 | intronic |
| XP03GO | chr20 | 8125823 | PLCB1 | intronic |
| XP03GO | chr3 | 42201917 | TRAK1 | intronic |
| XP03GO | chr3 | 142566603 | ATR | intronic |
| XP03GO | chr4 | 40797843 | NSUN7 | intronic |
| XP03GO | chr4 | 98517880 | TSPAN5 | intronic |
| XP03GO | chr4 | 188107213 | TRIML2 | intronic |
| XP03GO | chr5 | 87281071 | RASA1 | intronic |
| XP03GO | chr5 | 142977287 | ARHGAP26 | intronic |
| XP06GO_1 | chr12 | 62302147 | USP15 | intronic |
| XP06GO_1 | chr17 | 34151550 | ASIC2 | intronic |
| XP06GO_1 | chr3 | 14842879 | FGD5 | intronic |
| XP06GO_1 | chr3 | 180986068 | DNAJC19 | intronic |
| XP06GO_1 | chr3 | 186100783 | ETV5 | intronic |
| XP06GO_1 | chr5 | 11629149 | CTNND2 | intronic |
| XP06GO_1 | chr8 | 47677159 | SPIDR | intronic |
| XP06GO_1 | chr8 | 101739598 | NCALD | intronic |
| XP06GO_1 | chr8 | 133055875 | TG | intronic |
| XP06GO_2 | chr1 | 111959485 | KCND3 | intronic |
| XP06GO_2 | chr12 | 343209 | KDM5A | intronic |
| XP06GO_2 | chr12 | 106594694 | RFX4 | intronic |

| | | | | |
|---|---|---|---|---|
| XP06GO_2 | chr2 | 23972787 | UBXN2A | intronic |
| XP06GO_2 | chr21 | 29297506 | BACH1 | intronic |
| XP06GO_2 | chr3 | 12829439 | CAND2 | intronic |
| XP06GO_2 | chr3 | 48683547 | NCKIPSD | intronic |
| XP06GO_2 | chr4 | 99413345 | ADH7 | intronic |
| XP06GO_2 | chr5 | 39115382 | FYB | intronic |
| XP06GO_2 | chr6 | 6193235 | F13A1 | intronic |
| XP06GO_2 | chr6 | 108589443 | FOXO3 | intronic |
| XP06GO_4 | chr1 | 22729253 | EPHB2 | intronic |
| XP06GO_4 | chr1 | 201303808 | PKP1 | intronic |
| XP06GO_4 | chr2 | 223908560 | WDFY1 | intronic |
| XP11GO | chr1 | 176028861 | RFWD2 | intronic |
| XP11GO | chr16 | 57100779 | CPNE2 | intronic |
| XP11GO | chr16 | 68569361 | ZFP90 | intronic |
| XP11GO | chr17 | 11328943 | SHISA6 | intronic |
| XP11GO | chr17 | 81957100 | NOTUM | intronic |
| XP11GO | chr18 | 7619420 | PTPRM | intronic |
| XP11GO | chr19 | 52308189 | ZNF480 | intronic |
| XP11GO | chr2 | 65092929 | RAB1A | intronic |
| XP11GO | chr22 | 30952155 | MORC2 | intronic |
| XP11GO | chr3 | 13367614 | NUP210 | intronic |
| XP11GO | chr4 | 6545462 | PPP2R2C | intronic |
| XP11GO | chr4 | 31026179 | PCDH7 | intronic |
| XP11GO | chr4 | 44258565 | KCTD8 | intronic |
| XP11GO | chr4 | 48809166 | OCIAD1 | intronic |
| XP11GO | chr4 | 109820286 | GAR1 | intronic |
| XP11GO | chr7 | 75951606 | POR | intronic |
| XP11GO | chr8 | 109478786 | PKHD1L1 | intronic |
| XP45GO | chr1 | 59552137 | FGGY | intronic |
| XP45GO | chr13 | 113727350 | GRK1 | intronic |
| XP45GO | chr2 | 178495137 | PLEKHA3 | intronic |
| XP45GO | chr22 | 32793639 | SYN3 | intronic |
| XP45GO | chr3 | 173636930 | NLGN1 | intronic |
| XP45GO | chr4 | 168482442 | DDX60L | intronic |
| XP45GO | chr4 | 185251360 | SNX25 | intronic |
| XP52GO_1 | chr1 | 175342356 | TNR | intronic |
| XP52GO_1 | chr10 | 45578006 | MARCH8 | intronic |
| XP52GO_1 | chr12 | 128466525 | TMEM132C | intronic |
| XP52GO_1 | chr14 | 72143500 | RGS6 | intronic |
| XP52GO_1 | chr17 | 10722502 | TMEM220 | intronic |
| XP52GO_1 | chr2 | 31985611 | MEMO1 | intronic |
| XP52GO_1 | chr2 | 203522644 | RAPH1 | intronic |
| XP52GO_1 | chr6 | 124364609 | NKAIN2 | intronic |
| XP52GO_1 | chr6 | 167871481 | MLLT4 | intronic |

| | | | | |
|---|---|---|---|---|
| XP52GO_2 | chr7 | 131251214 | MKLN1 | intronic |
| XP85GO_1 | chr10 | 99365030 | CNNM1 | intronic |
| XP85GO_1 | chr11 | 74699711 | CHRDL2 | intronic |
| XP85GO_1 | chr15 | 40258743 | PAK6 | intronic |
| XP85GO_1 | chr15 | 55565856 | PYGO1 | intronic |
| XP85GO_1 | chr19 | 46607282 | CALM3 | intronic |
| XP85GO_1 | chr22 | 44887892 | PHF21B | intronic |
| XP85GO_1 | chr3 | 177037638 | TBL1XR1 | intronic |
| XP85GO_1 | chr6 | 150811672 | PLEKHG1 | intronic |
| XP85GO_2 | chr1 | 114738217 | CSDE1 | intronic |
| XP85GO_2 | chr1 | 161282685 | PCP4L1 | intronic |
| XP85GO_2 | chr11 | 64599164 | SLC22A12 | intronic |
| XP85GO_2 | chr17 | 6108894 | WSCD1 | intronic |
| XP85GO_2 | chr17 | 57424738 | MSI2 | intronic |
| XP85GO_2 | chr19 | 17555979 | COLGALT1 | intronic |
| XP85GO_2 | chr2 | 45823048 | PRKCE | intronic |
| XP85GO_2 | chr2 | 143674834 | ARHGAP15 | intronic |
| XP85GO_2 | chr2 | 159189792 | TANC1 | intronic |
| XP85GO_2 | chr22 | 26623361 | CRYBA4 | intronic |
| XP85GO_2 | chr5 | 123001489 | SNX24 | intronic |
| XP85GO_2 | chr7 | 151395256 | WDR86 | intronic |
| XP85GO_2 | chr8 | 50082791 | SNTG1 | intronic |
| XP85GO_2 | chr8 | 62314115 | NKAIN3 | intronic |

| Supplementary Table 9 | | | | |
|---|---|---|---|---|
| EntrezID | Gene symbol | Mutation count | ChrmLoc start | ChrmLoc end |
| 100131827 | ZNF717 | 150 | -75729960 | -75785549 |
| 996 | CDC27 | 82 | -47117702 | -47189295 |
| 7273 | TTN | 80 | -178525990 | -178807423 |
| 94025 | MUC16 | 41 | -8848843 | -8981342 |
| 3321 | IGSF3 | 35 | -116574407 | -116667733 |
| 5042 | PABPC3 | 34 | 25096135 | 25099254 |
| 154664 | ABCA13 | 26 | 48171457 | 48647497 |
| 259293 | TAS2R30 | 25 | -11133284 | -11134244 |
| 150483 | TEKT4 | 25 | 94871429 | 94876823 |
| 8481 | OFD1 | 25 | 13734712 | 13769353 |
| 54768 | HYDIN | 22 | 0 | -93427 |
| 643677 | CCDC168 | 22 | -102729366 | -102759072 |
| 129446 | XIRP2 | 22 | 166888479 | 167259751 |
| 84059 | ADGRV1 | 22 | 90558796 | 91164437 |
| 387893 | KMT5A | 21 | 123384131 | 123409353 |
| 57730 | ANKRD36B | 19 | -97504797 | -97589965 |
| 1767 | DNAH5 | 19 | -13690328 | -13944688 |
| 1488 | CTBP2 | 18 | -124984316 | -125006018 |
| 54798 | DCHS2 | 17 | -154231741 | -154491799 |
| 7399 | USH2A | 17 | -216173947 | -216423448 |
| 23345 | SYNE1 | 17 | -152121683 | -152636851 |
| 338 | APOB | 16 | -21001428 | -21044073 |
| 401024 | FSIP2 | 16 | 185738803 | 185833290 |
| 100288801 | FRG2C | 15 | 75664190 | 75667217 |
| 4585 | MUC4 | 14 | 16276 | 47963 |
| 114788 | CSMD3 | 14 | -112222929 | -113377152 |
| 93035 | PKHD1L1 | 14 | 109362460 | 109537207 |
| 10106 | CTDSP2 | 14 | -57819926 | -57846729 |
| 158471 | PRUNE2 | 13 | -76611375 | -76906087 |
| 27445 | PCLO | 13 | -82820477 | -83162884 |
| 53353 | LRP1B | 13 | -140231422 | -142131016 |
| 6262 | RYR2 | 13 | 237042183 | 237833988 |
| 1286 | COL4A4 | 13 | -227002715 | -227164205 |
| 25832 | NBPF14 | 12 | -148531384 | -148595717 |
| 1285 | COL4A3 | 12 | 227164623 | 227314792 |
| 144535 | CFAP54 | 12 | 96489576 | 96875555 |
| 79776 | ZFHX4 | 12 | 76681246 | 76867281 |
| 6101 | RP1 | 12 | 54616095 | 54630834 |
| 6708 | SPTA1 | 12 | -158610703 | -158686715 |
| 6263 | RYR3 | 12 | 33310966 | 33866121 |
| 6323 | SCN1A | 11 | -165984640 | -166128020 |

| | | | | |
|---|---|---|---|---|
| 340990 | OTOG | 11 | 17547258 | 17646044 |
| 138474 | TAF1L | 11 | -32629453 | -32635669 |
| 728378 | POTEF | 11 | -130073534 | -130129222 |
| 4588 | MUC6 | 11 | -1012822 | -1036718 |
| 2483 | FRG1 | 11 | 189940871 | 189963191 |
| 80309 | SPHKAP | 11 | -227979953 | -228181645 |
| 221981 | THSD7A | 10 | -11370364 | -11832198 |
| 169044 | COL22A1 | 10 | -138588234 | -138914041 |
| 2678 | GGT1 | 10 | 24583749 | 24629005 |
| 64478 | CSMD1 | 10 | -2935360 | -4994914 |
| 139378 | ADGRG4 | 10 | 136300962 | 136416890 |
| 285175 | UNC80 | 10 | 209771831 | 209999295 |
| 58508 | KMT2C | 10 | -152134924 | -152436003 |
| 286464 | CFAP47 | 10 | 35919733 | 35990150 |
| 8701 | DNAH11 | 10 | 21543038 | 21901839 |
| 8029 | CUBN | 9 | -16823965 | -17129811 |
| 9177 | HTR3B | 9 | 113908840 | 113946561 |
| 55870 | ASH1L | 9 | -155335267 | -155562803 |
| 319089 | TTC6 | 9 | 37772382 | 37842717 |
| 389763 | SPATA31D1 | 9 | 81988771 | 81995253 |
| 256076 | COL6A5 | 9 | 130345515 | 130484846 |
| 388697 | HRNR | 9 | -152212075 | -152224193 |
| 56171 | DNAH7 | 9 | -195737702 | -196068837 |
| 375248 | ANKRD36 | 9 | 97113152 | 97264520 |
| 4036 | LRP2 | 9 | -169127108 | -169362534 |
| 1769 | DNAH8 | 9 | 38715310 | 39030792 |
| 91074 | ANKRD30A | 9 | 37125856 | 37232567 |
| 79026 | AHNAK | 9 | -62515901 | -62546806 |
| 4619 | MYH1 | 8 | -10492309 | -10518542 |
| 317754 | POTED | 8 | 13609776 | 13645823 |
| 1755 | DMBT1 | 8 | 122560753 | 122643736 |
| 56154 | TEX15 | 8 | -30831543 | -30913008 |
| 56776 | FMN2 | 8 | 240091882 | 240475185 |
| 287 | ANK2 | 8 | 112860986 | 113383740 |
| 2260 | FLG | 8 | -38411137 | -38467845 |
| 1281 | COL3A1 | 8 | 188974372 | 189012746 |
| 5649 | RELN | 8 | -103471783 | -103989516 |
| 8913 | CACNA1G | 8 | 50560714 | 50627471 |
| 2977 | GUCY1A2 | 8 | -106674018 | -107018476 |
| 5314 | PKHD1 | 8 | -51720848 | -52087613 |
| 5789 | PTPRD | 8 | -8314245 | -10613002 |
| 26047 | CNTNAP2 | 8 | 2412 | 204591 |
| 140766 | ADAMTS14 | 8 | 70672505 | 70762439 |
| 91156 | IGFN1 | 8 | 201190823 | 201228952 |

| | | | | |
|---|---|---|---|---|
| 79747 | ADGB | 8 | 146598971 | 146815461 |
| 8516 | ITGA8 | 8 | -15513948 | -15720335 |
| 9353 | SLIT2 | 8 | 20251904 | 20620561 |
| 7204 | TRIO | 7 | 14143341 | 14510204 |
| 22915 | MMRN1 | 7 | 89894853 | 89954610 |
| 140803 | TRPM6 | 7 | -74722494 | -74887347 |
| 3791 | KDR | 7 | -55078480 | -55125595 |
| 6299 | SALL1 | 7 | -51135974 | -51150597 |
| 60676 | PAPPA2 | 7 | 176463174 | 176691205 |
| 5334 | PLCL1 | 7 | 197804592 | 198149863 |
| 773 | CACNA1A | 7 | -13206442 | -13506479 |
| 114805 | GALNT13 | 7 | 153871921 | 154454177 |
| 65217 | PCDH15 | 7 | -53821098 | -54801231 |
| 131873 | COL6A6 | 7 | 130560333 | 130678137 |
| 131544 | CRYBG3 | 7 | 97822010 | 97944984 |
| 8972 | MGAM | 7 | 410 | 68627 |
| 6261 | RYR1 | 7 | 38433690 | 38587564 |
| 463 | ZFHX3 | 7 | -72782886 | -73059021 |
| 4052 | LTBP1 | 7 | 33134595 | 33399508 |
| 57705 | WDFY4 | 7 | 48685462 | 48982956 |
| 221806 | VWDE | 7 | -12330884 | -12403941 |
| 10763 | NES | 7 | -156668762 | -156677407 |
| 1129 | CHRM2 | 7 | 136869245 | 137020213 |
| 2153 | F5 | 7 | -169511950 | -169586481 |
| 577 | ADGRB3 | 7 | 68635281 | 69389506 |
| 83872 | HMCN1 | 7 | 185734390 | 186190949 |
| 84239 | ATP13A4 | 7 | -193398966 | -193554895 |
| 6332 | SCN7A | 7 | -166403572 | -166486971 |
| 120114 | FAT3 | 7 | 92224817 | 92896473 |
| 3802 | KIR2DL3 | 7 | 71370 | 85919 |
| 55567 | DNAH3 | 7 | -20933110 | -21159441 |
| 545 | ATR | 7 | -142449234 | -142578733 |
| 26278 | SACS | 7 | -23328830 | -23375508 |
| 53904 | MYO3A | 7 | 25934228 | 26023488 |
| 79799 | UGT2A3 | 7 | -68928463 | -68951804 |
| 2200 | FBN1 | 7 | -48408312 | -48645709 |
| 2913 | GRM3 | 7 | 86643913 | 86864876 |
| 5800 | PTPRO | 7 | 15546610 | 15598331 |
| 168090 | C6orf118 | 6 | -165279663 | -165309605 |
| 284217 | LAMA1 | 6 | -6941741 | -7117797 |
| 79633 | FAT4 | 6 | 125314954 | 125492932 |
| 6098 | ROS1 | 6 | -117287352 | -117425942 |
| 23198 | PSME4 | 6 | -53864068 | -53970993 |
| 344148 | NCKAP5 | 6 | -132671789 | -133568463 |

| | | | | |
|---|---|---|---|---|
| 56131 | PCDHB4 | 6 | 141121817 | 141125621 |
| 8671 | SLC4A4 | 6 | 71339019 | 71572083 |
| 9381 | OTOF | 6 | -26457202 | -26478124 |
| 121256 | TMEM132D | 6 | -129071725 | -129904025 |
| 4626 | MYH8 | 6 | -10390322 | -10421950 |
| 26033 | ATRNL1 | 6 | 115093364 | 115171369 |
| 26166 | RGS22 | 6 | -99960935 | -100105626 |
| 23338 | JADE2 | 6 | 134525669 | 134583227 |
| 8499 | PPFIA2 | 6 | -81257974 | -81598371 |
| 5979 | RET | 6 | 43104731 | 43127614 |
| 55636 | CHD7 | 6 | 60741258 | 60868027 |
| 727897 | MUC5B | 6 | 1223065 | 1262172 |
| 84700 | MYO18B | 6 | 25742187 | 26031045 |
| 23524 | SRRM2 | 6 | 2752637 | 2771412 |
| 1770 | DNAH9 | 6 | 11882901 | 11969748 |
| 1496 | CTNNA2 | 6 | 79523182 | 80648863 |
| 6328 | SCN3A | 6 | -165087525 | -165204050 |
| 155038 | GIMAP8 | 6 | 150450629 | 150479393 |
| 84627 | ZNF469 | 6 | 88382958 | 88440751 |
| 1278 | COL1A2 | 6 | 94394894 | 94431227 |
| 730 | C7 | 6 | 40909496 | 40984643 |
| 57683 | ZDBF2 | 6 | 388647 | 428064 |
| 84181 | CHD6 | 6 | -41402082 | -41618377 |
| 375337 | TOPAZ1 | 6 | 44241885 | 44332098 |
| 83889 | WDR87 | 6 | -37884822 | -37906594 |
| 79987 | SVEP1 | 6 | -110365248 | -110579741 |
| 219578 | ZNF804B | 6 | 88759699 | 89338528 |
| 403273 | OR5H14 | 6 | 98149385 | 98150318 |
| 6326 | SCN2A | 6 | 165294054 | 165392304 |
| 4308 | TRPM1 | 6 | -3286820 | -3447037 |
| 8989 | TRPA1 | 6 | -72021249 | -72075584 |
| 5788 | PTPRC | 6 | 198639039 | 198695171 |
| 2066 | ERBB4 | 6 | -211375716 | -212538802 |
| 113146 | AHNAK2 | 6 | -104937252 | -104970507 |
| 1010 | CDH12 | 6 | -21750865 | -22213800 |
| 146754 | DNAH2 | 6 | 7717743 | 7833711 |
| 51725 | FBXO40 | 6 | 121593378 | 121630294 |
| 8021 | NUP214 | 6 | 131190138 | 131234663 |
| 400986 | ANKRD36C | 6 | -95848931 | -95991824 |
| 85445 | CNTNAP4 | 6 | 76277400 | 76560755 |
| 55187 | VPS13D | 6 | 12230029 | 12512046 |
| 9113 | LATS1 | 6 | -149658152 | -149718101 |
| 5801 | PTPRQ | 6 | -70638079 | -70920738 |
| 2042 | EPHA3 | 6 | 89107620 | 89400345 |

| | | | | |
|---|---|---|---|---|
| 140453 | MUC17 | 6 | 101020080 | 101058859 |
| 286234 | SPATA31E1 | 6 | 87882876 | 87888902 |
| 50939 | IMPG2 | 6 | -101222545 | -101320575 |
| 6335 | SCN9A | 6 | -166195186 | -166375987 |
| 3339 | HSPG2 | 6 | -21822243 | -21937310 |
| 1131 | CHRM3 | 6 | 239386567 | 239915449 |
| 4625 | MYH7 | 6 | -23412739 | -23435660 |
| 729 | C6 | 6 | -41142145 | -41261469 |
| 7099 | TLR4 | 6 | 117704402 | 117724735 |
| 93034 | NT5C1B | 5 | -18562870 | -18589569 |
| 5144 | PDE4D | 5 | -58969040 | -59587625 |
| 5243 | ABCB1 | 5 | -87503016 | -87600884 |
| 231 | AR | 5 | -134442349 | -134459239 |
| 2565 | GABRG1 | 5 | -46035769 | -46124054 |
| 139105 | BEND2 | 5 | -18162930 | -18220886 |
| 120892 | LRRK2 | 5 | 40224996 | 40369284 |
| 1756 | DMD | 5 | -31119225 | -31266954 |
| 11148 | HHLA2 | 5 | 108296489 | 108378284 |
| 144568 | A2ML1 | 5 | 8822620 | 8876787 |
| 84075 | FSCB | 5 | -44504150 | -44507283 |
| 3681 | ITGAD | 5 | 31393334 | 31426505 |
| 386674 | KRTAP10-6 | 5 | -44591267 | -44592505 |
| 65125 | WNK1 | 5 | 752578 | 911452 |
| 440279 | UNC13C | 5 | 53978440 | 54633439 |
| 221061 | FAM171A1 | 5 | -15211644 | -15371289 |
| 1310 | COL19A1 | 5 | 69866555 | 70212467 |
| 1630 | DCC | 5 | 52340196 | 53535898 |
| 56126 | PCDHB10 | 5 | 141192352 | 141195647 |
| 2044 | EPHA5 | 5 | -65319566 | -65670489 |
| 9378 | NRXN1 | 5 | -49918504 | -51032536 |
| 1287 | COL4A5 | 5 | 108439837 | 108697545 |
| 1778 | DYNC1H1 | 5 | 101964572 | 102056443 |
| 114784 | CSMD2 | 5 | -33513997 | -34165230 |
| 64072 | CDH23 | 5 | 71811882 | 71815947 |
| 2335 | FN1 | 5 | -215360864 | -215436068 |
| 10178 | TENM1 | 5 | -124375905 | -124963817 |
| 282763 | OR51B5 | 5 | -5342513 | -5505652 |
| 5803 | PTPRZ1 | 5 | 121873160 | 122062036 |
| 2904 | GRIN2B | 5 | -13537336 | -13981602 |
| 6546 | SLC8A1 | 5 | -40097269 | -40452090 |
| 57493 | HEG1 | 5 | -124965709 | -125055997 |
| 8924 | HERC2 | 5 | 22494850 | 22579725 |
| 1306 | COL15A1 | 5 | 98943906 | 99070787 |
| 2915 | GRM5 | 5 | -88504642 | -89063659 |

| | | | | |
|---|---|---|---|---|
| 1261 | CNGA3 | 5 | 98346455 | 98398601 |
| 10902 | BRD8 | 5 | -138156883 | -138178669 |
| 653489 | RGPD3 | 5 | -106403405 | -106468413 |
| 83417 | FCRL4 | 5 | -157573747 | -157598085 |
| 1539 | CYLC2 | 5 | 102995332 | 103018488 |
| 1293 | COL6A3 | 5 | -237324011 | -237414207 |
| 57578 | UNC79 | 5 | 447743 | 822438 |
| 22839 | DLGAP4 | 5 | 36461467 | 36528633 |
| 26960 | NBEA | 5 | 35476808 | 35672736 |
| 51196 | PLCE1 | 5 | 93993930 | 94332823 |
| 23074 | UHRF1BP1L | 5 | -100069907 | -100142874 |
| 403274 | OR5H15 | 5 | 98168699 | 98169641 |
| 55083 | KIF26B | 5 | 245154984 | 245709432 |
| 84072 | HORMAD1 | 5 | -150698059 | -150720895 |
| 202333 | CMYA5 | 5 | 79689835 | 79800221 |
| 79674 | VEPH1 | 5 | -157259742 | -157499656 |
| 255631 | COL24A1 | 5 | -85729232 | -86156471 |
| 2157 | F8 | 5 | -154835792 | -155022723 |
| 81792 | ADAMTS12 | 5 | -33751195 | -33891990 |
| 83893 | SPATA16 | 5 | -172889356 | -173141235 |
| 8470 | SORBS2 | 5 | -185585443 | -185775854 |
| 147658 | ZNF534 | 5 | 52429147 | 52452309 |
| 23352 | UBR4 | 5 | -19074510 | -19210266 |
| 9919 | SEC16A | 5 | -136440104 | -136483040 |
| 131149 | OTOL1 | 5 | 161496807 | 161503942 |
| 92949 | ADAMTSL1 | 5 | 18474152 | 18684954 |
| 26053 | AUTS2 | 5 | 69598474 | 70793506 |
| 1826 | DSCAM | 5 | -40010998 | -40847158 |
| 221037 | JMJD1C | 5 | -63167220 | -63269223 |
| 2195 | FAT1 | 5 | -186587793 | -186723856 |
| 344905 | ATP13A5 | 5 | -193274788 | -193378753 |
| 256714 | MAP7D2 | 5 | -20006712 | -20116907 |
| 4928 | NUP98 | 5 | -3675018 | -3797554 |
| 114548 | NLRP3 | 5 | 247416172 | 247448817 |
| 346007 | EYS | 5 | -65334912 | -65579804 |
| 126859 | AXDND1 | 5 | 179366043 | 179554735 |
| 6476 | SI | 5 | -164978897 | -165078496 |
| 2318 | FLNC | 5 | 128830405 | 128859272 |
| 121364 | OR10A7 | 5 | 55221024 | 55221975 |
| 80731 | THSD7B | 5 | 136765544 | 137677718 |
| 84433 | CARD11 | 5 | -2906141 | -3043867 |
| 2903 | GRIN2A | 5 | -9753407 | -10182908 |
| 11141 | IL1RAPL1 | 5 | 28587445 | 29956713 |
| 56164 | STK31 | 5 | 23710218 | 23832511 |

| | | | | |
|---|---|---|---|---|
| 10071 | MUC12 | 5 | 100969622 | 101018949 |
| 728747 | ANKRD20A4 | 5 | NA | NA |
| 4018 | LPA | 5 | -160531481 | -160664275 |
| 22998 | LIMCH1 | 5 | 41360806 | 41700044 |
| 8925 | HERC1 | 5 | -63608617 | -63833948 |
| 3075 | CFH | 5 | 196652042 | 196747504 |
| 1302 | COL11A2 | 5 | -4357097 | -4386870 |
| 6547 | SLC8A3 | 5 | -70044214 | -70188951 |
| 1612 | DAPK1 | 5 | 87498534 | 87708634 |
| 9024 | BRSK2 | 5 | 1389933 | 1462689 |
| 1280 | COL2A1 | 5 | -47972966 | -48004476 |
| 56112 | PCDHGA3 | 5 | 141343828 | 141512973 |
| 477 | ATP1A2 | 5 | 160115758 | 160143584 |
| 5651 | TMPRSS15 | 5 | -18269115 | -18403785 |
| 27145 | FILIP1 | 5 | -75456389 | -75493800 |
| 2243 | FGA | 5 | -154583125 | -154590742 |
| 23230 | VPS13A | 5 | 77177444 | 77385190 |
| 114795 | TMEM132B | 5 | 125326615 | 125662377 |
| 1657 | DMXL1 | 5 | 119071054 | 119249129 |
| 65220 | NADK | 5 | -1751231 | -1758642 |
| 25834 | MGAT4C | 5 | -85973293 | -86256391 |
| 686 | BTD | 5 | 15601744 | 15653714 |
| 23165 | NUP205 | 5 | 135557916 | 135648751 |
| 1008 | CDH10 | 5 | -24487099 | -24644978 |
| 2070 | EYA4 | 5 | 133241356 | 133532120 |
| 55911 | APOBR | 5 | 28494642 | 28498964 |
| 8085 | KMT2D | 5 | -49018977 | -49060794 |
| 10847 | SRCAP | 5 | 30699170 | 30741409 |
| 57519 | STARD9 | 5 | 42575605 | 42720998 |
| 23499 | MACF1 | 5 | 39084166 | 39487138 |
| 491 | ATP2B2 | 5 | -10324022 | -10505586 |
| 64116 | SLC39A8 | 5 | -102261663 | -102324312 |
| 4647 | MYO7A | 5 | 77128245 | 77215241 |
| 4978 | OPCML | 5 | -132414664 | -132943637 |
| 57569 | ARHGAP20 | 5 | -110577042 | -110713189 |
| 81442 | OR6N2 | 5 | -158776681 | -158777635 |
| 29951 | PDZRN4 | 5 | 41188319 | 41574745 |
| 340527 | NHSL2 | 5 | 71911087 | 72143574 |
| 9734 | HDAC9 | 5 | 18086824 | 18668843 |
| 7776 | ZNF236 | 5 | 76823759 | 76972899 |

| Supplementary Table 10 | | | | |
|---|---|---|---|---|
| EntrezID | Gene symbol | Mutation count | ChrmLoc start | ChrmLoc end |
| 94025 | MUC16 | 41 | -8848843 | -8981342 |
| 114788 | CSMD3 | 14 | -112222929 | -113377152 |
| 4585 | MUC4 | 14 | 16276 | 47963 |
| 53353 | LRP1B | 13 | -140231422 | -142131016 |
| 58508 | KMT2C | 10 | -152134924 | -152436003 |
| 26047 | CNTNAP2 | 8 | 2412 | 204591 |
| 5789 | PTPRD | 8 | -8314245 | -10613002 |
| 1281 | COL3A1 | 8 | 188974372 | 189012746 |
| 2913 | GRM3 | 7 | 86643913 | 86864876 |
| 3791 | KDR | 7 | -55078480 | -55125595 |
| 120114 | FAT3 | 7 | 92224817 | 92896473 |
| 463 | ZFHX3 | 7 | -72782886 | -73059021 |
| 545 | ATR | 7 | -142449234 | -142578733 |
| 2066 | ERBB4 | 6 | -211375716 | -212538802 |
| 8021 | NUP214 | 6 | 131190138 | 131234663 |
| 5979 | RET | 6 | 43104731 | 43127614 |
| 6098 | ROS1 | 6 | -117287352 | -117425942 |
| 2042 | EPHA3 | 6 | 89107620 | 89400345 |
| 79633 | FAT4 | 6 | 125314954 | 125492932 |
| 9113 | LATS1 | 6 | -149658152 | -149718101 |
| 5788 | PTPRC | 6 | 198639039 | 198695171 |
| 1496 | CTNNA2 | 6 | 79523182 | 80648863 |
| 83417 | FCRL4 | 5 | -157573747 | -157598085 |
| 2903 | GRIN2A | 5 | -9753407 | -10182908 |
| 1630 | DCC | 5 | 52340196 | 53535898 |
| 1280 | COL2A1 | 5 | -47972966 | -48004476 |
| 8085 | KMT2D | 5 | -49018977 | -49060794 |
| 231 | AR | 5 | -134442349 | -134459239 |
| 653489 | RGPD3 | 5 | -106403405 | -106468413 |
| 2195 | FAT1 | 5 | -186587793 | -186723856 |
| 26960 | NBEA | 5 | 35476808 | 35672736 |
| 84433 | CARD11 | 5 | -2906141 | -3043867 |
| 4928 | NUP98 | 5 | -3675018 | -3797554 |
| 1008 | CDH10 | 5 | -24487099 | -24644978 |
| 2033 | EP300 | 4 | 41092591 | 41180077 |
| 80312 | TET1 | 4 | 68560336 | 68694485 |
| 7015 | TERT | 4 | -1253147 | -1295068 |
| 5783 | PTPN13 | 4 | 86594314 | 86815159 |
| 4214 | MAP3K1 | 4 | 56815548 | 56896151 |
| 23365 | ARHGEF12 | 4 | 120385326 | 120489937 |
| 6491 | STIL | 4 | -47250138 | -47314147 |

| | | | | |
|---|---|---|---|---|
| 57448 | BIRC6 | 4 | 32357027 | 32618898 |
| 23261 | CAMTA1 | 4 | 6785323 | 6888201 |
| 4853 | NOTCH2 | 4 | -119935112 | -120069703 |
| 2045 | EPHA7 | 4 | -93409653 | -93419559 |
| 675 | BRCA2 | 4 | 32315507 | 32400268 |
| 57670 | KIAA1549 | 4 | -138831380 | -138981389 |
| 283149 | BCL9L | 3 | -118896135 | -118910542 |
| 23476 | BRD4 | 3 | -15247035 | -15280451 |
| 10320 | IKZF1 | 3 | 50304715 | 50405100 |
| 80243 | PREX2 | 3 | 67952045 | 68106009 |
| 9611 | NCOR1 | 3 | -16091569 | -16194639 |
| 6495 | SIX1 | 3 | -60643432 | -60649477 |
| 492 | ATP2B3 | 3 | 153536121 | 153582929 |
| 6424 | SFRP4 | 3 | -37905932 | -37916817 |
| 2132 | EXT2 | 3 | 44095677 | 44251961 |
| 3702 | ITK | 3 | 157180839 | 157255185 |
| 168975 | CNBD1 | 3 | 86866414 | 87382859 |
| 23067 | SETD1B | 3 | 121804008 | 121832656 |
| 7409 | VAV1 | 3 | 6772707 | 6857361 |
| 472 | ATM | 3 | 108223066 | 108369102 |
| 6756 | SSX1 | 3 | 48255391 | 48267444 |
| 4297 | KMT2A | 3 | 118436491 | 118526832 |
| 2177 | FANCD2 | 3 | 10026436 | 10099343 |
| 25925 | ZNF521 | 3 | -25061923 | -25352166 |
| 4627 | MYH9 | 3 | -36281279 | -36387967 |
| 5787 | PTPRB | 3 | -70515869 | -70637429 |
| 5077 | PAX3 | 3 | -222293634 | -222298998 |
| 2065 | ERBB3 | 3 | 56080107 | 56085617 |
| 1788 | DNMT3A | 3 | -25281451 | -25341925 |
| 57674 | RNF213 | 3 | 80260851 | 80321462 |
| 2316 | FLNA | 3 | -154348531 | -154374638 |
| 1015 | CDH17 | 3 | -94127161 | -94217278 |
| 2322 | FLT3 | 3 | -28003273 | -28100576 |
| 10142 | AKAP9 | 3 | 91940861 | 92110673 |
| 8241 | RBM10 | 3 | 47145220 | 47186813 |
| 63976 | PRDM16 | 3 | 3069202 | 3438621 |
| 867 | CBL | 3 | 119206338 | 119308149 |
| 3718 | JAK3 | 3 | -17824783 | -17847982 |
| 9582 | APOBEC3B | 2 | 38982346 | 38992779 |
| 9203 | ZMYM3 | 2 | -71249204 | -71254649 |
| 6760 | SS18 | 2 | -26016252 | -26090647 |
| 6446 | SGK1 | 2 | -134169255 | -134174866 |
| 5604 | MAP2K1 | 2 | 66386911 | 66491544 |
| 668 | FOXL2 | 2 | -138944223 | -138947137 |

| | | | | |
|---|---|---|---|---|
| 9612 | NCOR2 | 2 | -124324414 | -124567612 |
| 2778 | GNAS | 2 | 58839747 | 58911192 |
| 5324 | PLAG1 | 2 | -56160908 | -56211273 |
| 2099 | ESR1 | 2 | 151805704 | 152103273 |
| 23405 | DICER1 | 2 | -95086227 | -95158010 |
| 55728 | N4BP2 | 2 | 40056849 | 40158252 |
| 604 | BCL6 | 2 | -187721380 | -187736501 |
| 8805 | TRIM24 | 2 | 138460258 | 138589996 |
| 26040 | SETBP1 | 2 | 44680072 | 44877419 |
| 29102 | DROSHA | 2 | -31400494 | -31532093 |
| 54738 | FEV | 2 | -218981086 | -218985184 |
| 6801 | STRN | 2 | -36837697 | -36966536 |
| 8289 | ARID1A | 2 | 26696014 | 26782104 |
| 23085 | ERC1 | 2 | 991222 | 1495931 |
| 5395 | PMS2 | 2 | -5970925 | -6009049 |
| 5537 | PPP6C | 2 | -125146572 | -125189939 |
| 4791 | NFKB2 | 2 | 102394109 | 102402529 |
| 238 | ALK | 2 | -29192773 | -29223900 |
| 2719 | GPC3 | 2 | -133535745 | -133985594 |
| 3815 | KIT | 2 | 54657956 | 54740715 |
| 5292 | PIM1 | 2 | 37170151 | 37175428 |
| 1277 | COL1A1 | 2 | -50184100 | -50201631 |
| 580 | BARD1 | 2 | -214725645 | -214809683 |
| 4773 | NFATC2 | 2 | -51386962 | -51542719 |
| 29974 | A1CF | 2 | -50799408 | -50885675 |
| 1501 | CTNND2 | 2 | -10971840 | -11588917 |
| 6608 | SMO | 2 | 129188632 | 129213544 |
| 54880 | BCOR | 2 | -40051252 | -40177277 |
| 64324 | NSD1 | 2 | 177134228 | 177300213 |
| 6926 | TBX3 | 2 | -114670254 | -114684175 |
| 5426 | POLE | 2 | -132623761 | -132687342 |
| 8929 | PHOX2B | 2 | -41744081 | -41748725 |
| 2122 | MECOM | 2 | -169083506 | -169663712 |
| 3899 | AFF3 | 2 | -99545418 | -100105583 |
| 8295 | TRRAP | 2 | 98878489 | 99013243 |
| 6249 | CLIP1 | 2 | -122271430 | -122422669 |
| 1436 | CSF1R | 2 | -150053290 | -150113372 |
| 6000 | RGS7 | 2 | -240775513 | -241357230 |
| 4152 | PCM1 | 2 | -50266881 | -50281774 |
| 27436 | EML4 | 2 | 42169352 | 42332548 |
| 3371 | TNC | 2 | -115019575 | -115118157 |
| 2324 | FLT4 | 2 | -180601505 | -180649600 |
| 9098 | USP6 | 2 | 5128808 | 5174991 |
| 2064 | ERBB2 | 2 | 39700063 | 39728658 |

| | | | | |
|---|---|---|---|---|
| 4644 | MYO5A | 2 | -52307283 | -52528880 |
| 65268 | WNK2 | 2 | 93184929 | 93320572 |
| 5087 | PBX1 | 2 | 164559359 | 164851823 |
| 4436 | MSH2 | 2 | 47403066 | 47483228 |
| 6935 | ZEB1 | 2 | 31319215 | 31529804 |
| 324 | APC | 2 | 112737884 | 112846239 |
| 8313 | AXIN2 | 2 | -65528564 | -65561648 |
| 862 | RUNX1T1 | 2 | -91954971 | -92095230 |
| 51059 | FAM135B | 2 | -138130022 | -138497730 |
| 3195 | TLX1 | 2 | 101131299 | 101137789 |
| 345930 | ECT2L | 2 | 138796110 | 138904070 |
| 5187 | PER1 | 2 | -8140471 | -8152404 |
| 57105 | CYSLTR2 | 2 | 48653928 | 48711225 |
| 92017 | SNX29 | 2 | 11976733 | 12574287 |
| 286 | ANK1 | 2 | -41653224 | -41896741 |
| 1387 | CREBBP | 2 | -3725054 | -3880120 |
| 6938 | TCF12 | 2 | 57219429 | 57289853 |
| 23013 | SPEN | 2 | 15847706 | 15940456 |
| 3575 | IL7R | 2 | 35856890 | 35879603 |
| 346389 | MACC1 | 2 | -20134655 | -20217384 |
| 776 | CACNA1D | 2 | 53494610 | 53813730 |
| 29072 | SETD2 | 2 | -47016435 | -47164113 |
| 7486 | WRN | 2 | 31033809 | 31176137 |
| 476 | ATP1A1 | 1 | 116383369 | 116404776 |
| 84101 | USP44 | 1 | -95516559 | -95548802 |
| 4914 | NTRK1 | 1 | 156860878 | 156881850 |
| 4595 | MUTYH | 1 | -45329241 | -45340440 |
| 7175 | TPR | 1 | -186311653 | -186375253 |
| 140885 | SIRPA | 1 | 1895406 | 1940592 |
| 9967 | THRAP3 | 1 | 36224442 | 36305356 |
| 10274 | STAG1 | 1 | -136336235 | -136752378 |
| 9321 | TRIP11 | 1 | -91965990 | -92040059 |
| 1964 | EIF1AX | 1 | -20124524 | -20141838 |
| 85414 | SLC45A3 | 1 | -205657850 | -205680509 |
| 4233 | MET | 1 | 116672195 | 116769910 |
| 10721 | POLQ | 1 | -121431430 | -121545988 |
| 7048 | TGFBR2 | 1 | 30606600 | 30694141 |
| 51684 | SUFU | 1 | 102503971 | 102633535 |
| 5900 | RALGDS | 1 | -133097721 | -133129401 |
| 64979 | BRIP1 | 1 | -1798384 | -1799837 |
| 51592 | TRIM33 | 1 | -114392792 | -114511203 |
| 1108 | CHD4 | 1 | -6570081 | -6607379 |
| 595 | CCND1 | 1 | 69641155 | 69654474 |
| 7994 | KAT6A | 1 | -41942048 | -42051987 |

| | | | | |
|---|---|---|---|---|
| 114825 | PWWP2A | 1 | -160075884 | -160119450 |
| 4629 | MYH11 | 1 | -1361145 | -1515023 |
| 5450 | POU2AF1 | 1 | -111352255 | -111379275 |
| 2115 | ETV1 | 1 | -13891230 | -13991425 |
| 1616 | DAXX | 1 | -33318557 | -33323016 |
| 3091 | HIF1A | 1 | 61695512 | 61748258 |
| 10019 | SH2B3 | 1 | 111434861 | 111451623 |
| 2078 | ERG | 1 | -38380027 | -38498504 |
| 7253 | TSHR | 1 | 80955620 | 81108940 |
| 3105 | HLA-A | 1 | 1144512 | 1289979 |
| 51176 | LEF1 | 1 | -108047544 | -108168956 |
| 3845 | KRAS | 1 | -25205245 | -25250929 |
| 607 | BCL9 | 1 | 147541500 | 147626214 |
| 6389 | SDHA | 1 | 218319 | 257082 |
| 5796 | PTPRK | 1 | -313982 | -869933 |
| 9639 | ARHGEF10 | 1 | 1823925 | 1958641 |
| 641 | BLM | 1 | 90717345 | 90816166 |
| 3092 | HIP1 | 1 | -75533297 | -75738941 |
| 5156 | PDGFRA | 1 | 54229292 | 54298245 |
| 3662 | IRF4 | 1 | 391751 | 411443 |
| 11177 | BAZ1A | 1 | -34752730 | -34875360 |
| 5884 | RAD17 | 1 | 69371034 | 69414801 |
| 7799 | PRDM2 | 1 | 13749414 | 13788081 |
| 8496 | PPFIBP1 | 1 | 27524205 | 27695564 |
| 9968 | MED12 | 1 | 71118595 | 71142449 |
| 2735 | GLI1 | 1 | 57460150 | 57472264 |
| 4913 | NTHL1 | 1 | -2039819 | -2047834 |
| 405 | ARNT | 1 | -150809713 | -150876599 |
| 8028 | MLLT10 | 1 | 21534171 | 21557185 |
| 653 | BMP5 | 1 | -55753652 | -55875590 |
| 6777 | STAT5B | 1 | -42199176 | -42276391 |
| 7248 | TSC1 | 1 | -132891348 | -132944616 |
| 3725 | JUN | 1 | -58780790 | -58784047 |
| 26524 | LATS2 | 1 | -20973035 | -21061586 |
| 2264 | FGFR4 | 1 | 177086914 | 177098144 |
| 7531 | YWHAE | 1 | -1344274 | -1400222 |
| 27 | ABL2 | 1 | -179099330 | -179229677 |
| 613 | BCR | 1 | 23180508 | 23318037 |
| 999 | CDH1 | 1 | 68737291 | 68835536 |
| 1656 | DDX6 | 1 | -118747763 | -118791164 |
| 2475 | MTOR | 1 | -11106534 | -11262551 |
| 4851 | NOTCH1 | 1 | -136494432 | -136546048 |
| 6917 | TCEA1 | 1 | -53966555 | -54022448 |
| 8648 | NCOA1 | 1 | 24584476 | 24770701 |

| | | | | |
|---|---|---|---|---|
| 3977 | LIFR | 1 | -38474667 | -38595405 |
| 2006 | ELN | 1 | 74028172 | 74069906 |
| 5093 | PCBP1 | 1 | 70087476 | 70089202 |
| 23092 | ARHGAP26 | 1 | 142770376 | 143229011 |
| 2308 | FOXO1 | 1 | -40555666 | -40666641 |
| 27125 | AFF4 | 1 | -132875395 | -132963634 |
| 90827 | ZNF479 | 1 | -57119618 | -57139864 |
| 64919 | BCL11B | 1 | -99169287 | -99272197 |
| 3084 | NRG1 | 1 | 32548310 | 32728618 |
| 1009 | CDH11 | 1 | -64943752 | -65122063 |
| 7128 | TNFAIP3 | 1 | 137867258 | 137883312 |
| 2309 | FOXO3 | 1 | 108559824 | 108684774 |
| 6605 | SMARCE1 | 1 | -40624961 | -40647818 |
| 23157 | SEPT6 | 1 | -119615724 | -119693168 |
| 9869 | SETDB1 | 1 | 150926362 | 150945320 |
| 4088 | SMAD3 | 1 | 67166154 | 67195195 |
| 7403 | KDM6A | 1 | 44873174 | 45112612 |
| 10735 | STAG2 | 1 | 123961705 | 124102655 |
| 330 | BIRC3 | 1 | 102317483 | 102339403 |
| 1655 | DDX5 | 1 | -64498253 | -64506866 |
| 6927 | HNF1A | 1 | 120978542 | 121002511 |
| 2262 | GPC5 | 1 | 91398620 | 92867234 |
| 1345 | COX6C | 1 | -99877864 | -99893707 |
| 57492 | ARID1B | 1 | 157036426 | 157210779 |
| 1956 | EGFR | 1 | 55019020 | 55208080 |
| 1500 | CTNND1 | 1 | 57761801 | 57819540 |
| 25766 | PRPF40B | 1 | 49623560 | 49644665 |
| 3227 | HOXC11 | 1 | 53973125 | 53977643 |
| 2000 | ELF4 | 1 | -130063957 | -130110497 |
| 6092 | ROBO2 | 1 | 77098011 | 77649963 |
| 2199 | FBLN2 | 1 | 13568739 | 13638402 |
| 5925 | RB1 | 1 | 48303750 | 48481890 |
| 8218 | CLTCL1 | 1 | -19179473 | -19291719 |
| 4610 | MYCL | 1 | -39899611 | -39901917 |
| 57167 | SALL4 | 1 | -51782330 | -51802521 |
| 5727 | PTCH1 | 1 | -95442979 | -95509266 |
| 3209 | HOXA13 | 1 | -27194370 | -27200091 |
| 8243 | SMC1A | 1 | -53374148 | -53422728 |
| 9715 | FAM131B | 1 | -143353399 | -143362713 |
| 4089 | SMAD4 | 1 | 51030212 | 51085041 |
| 1662 | DDX10 | 1 | 108665068 | 108940926 |
| 6421 | SFPQ | 1 | -35182935 | -35193145 |
| 442444 | FAM47C | 1 | 37008365 | 37011664 |
| 5605 | MAP2K2 | 1 | -4090321 | -4124122 |

| | | | | |
|---|---|---|---|---|
| 1639 | DCTN1 | 1 | -74361154 | -74374667 |
| 3572 | IL6ST | 1 | -55935096 | -55994963 |
| 23305 | ACSL6 | 1 | -131949973 | -132011817 |
| 340578 | DCAF12L2 | 1 | -126163498 | -126166289 |
| 2623 | GATA1 | 1 | 48786589 | 48794310 |
| 10342 | TFG | 1 | 100709289 | 100748967 |
| 4609 | MYC | 1 | 127735433 | 127742951 |
| 7454 | WAS | 1 | 48683798 | 48691426 |
| 54790 | TET2 | 1 | 105146875 | 105279803 |
| 51755 | CDK12 | 1 | 39461485 | 39534565 |
| 27044 | SND1 | 1 | 127652193 | 128092593 |
| 5894 | RAF1 | 1 | -12583600 | -12664117 |
| 3762 | KCNJ5 | 1 | 128891355 | 128921163 |
| 2625 | GATA3 | 1 | 8054687 | 8075198 |
| 4261 | CIITA | 1 | 10877201 | 10936393 |
| 8493 | PPM1D | 1 | 60600192 | 60666279 |
| 399 | RHOH | 1 | 40196906 | 40244764 |
| 54894 | RNF43 | 1 | -58353675 | -58417534 |
| 6657 | SOX2 | 1 | 181711924 | 181714435 |
| 2175 | FANCA | 1 | -89737548 | -89816647 |
| 11328 | FKBP9 | 1 | 32957403 | 33006931 |
| 7157 | TP53 | 1 | -7668401 | -7687550 |
| 4330 | MN1 | 1 | -27748276 | -27801756 |
| 55422 | ZNF331 | 1 | 53521013 | 53580269 |
| 11200 | CHEK2 | 1 | -28687742 | -28741834 |
| 5903 | RANBP2 | 1 | 108719481 | 108785809 |
| 27086 | FOXP1 | 1 | -70954709 | -71064924 |
| 3936 | LCP1 | 1 | -46125922 | -46182177 |

| Supplementary Table 22 | | |
|---|---|---|
| **Plasma membrane** | | |
| **ID** | **Gene symbol** | **Gene name** |
| 9177 | HTR3B | 5-hydroxytryptamine receptor 3B |
| 92949 | ADAMTSL1 | ADAMTS like 1 |
| 113146 | AHNAK2 | AHNAK nucleoprotein 2 |
| 79026 | AHNAK | AHNAK nucleoprotein |
| 154664 | ABCA13 | ATP binding cassette subfamily A member 13 |
| 5243 | ABCB1 | ATP binding cassette subfamily B member 1 |
| 84239 | ATP13A4 | ATPase 13A4 |
| 344905 | ATP13A5 | ATPase 13A5 |
| 477 | ATP1A2 | ATPase Na+/K+ transporting subunit alpha 2 |
| 491 | ATP2B2 | ATPase plasma membrane Ca2+ transporting 2 |
| 114784 | CSMD2 | CUB and Sushi multiple domains 2 |
| 114788 | CSMD3 | CUB and Sushi multiple domains 3 |
| 1630 | DCC | DCC netrin 1 receptor |
| 22839 | DLGAP4 | DLG associated protein 4 |
| 1826 | DSCAM | DS cell adhesion molecule |
| 2042 | EPHA3 | EPH receptor A3 |
| 2044 | EPHA5 | EPH receptor A5 |
| 2195 | FAT1 | FAT atypical cadherin 1 |
| 120114 | FAT3 | FAT atypical cadherin 3 |
| 79633 | FAT4 | FAT atypical cadherin 4 |
| 8924 | HERC2 | HECT and RLD domain containing E3 ubiquitin protein ligase 2 |
| 53353 | LRP1B | LDL receptor related protein 1B |
| 4036 | LRP2 | LDL receptor related protein 2 |
| 317754 | POTED | POTE ankyrin domain family member D |
| 6098 | ROS1 | ROS proto-oncogene 1, receptor tyrosine kinase |
| 577 | ADGRB3 | adhesion G protein-coupled receptor B3 |
| 84059 | ADGRV1 | adhesion G protein-coupled receptor V1 |
| 287 | ANK2 | ankyrin 2 |
| 55911 | APOBR | apolipoprotein B receptor |
| 338 | APOB | apolipoprotein B |
| 1008 | CDH10 | cadherin 10 |
| 1010 | CDH12 | cadherin 12 |
| 64072 | CDH23 | cadherin related 23 |
| 773 | CACNA1A | calcium voltage-gated channel subunit alpha1 A |
| 8913 | CACNA1G | calcium voltage-gated channel subunit alpha1 G |
| 202333 | CMYA5 | cardiomyopathy associated 5 |
| 84433 | CARD11 | caspase recruitment domain family member 11 |
| 1129 | CHRM2 | cholinergic receptor muscarinic 2 |
| 1131 | CHRM3 | cholinergic receptor muscarinic 3 |
| 2153 | F5 | coagulation factor V |
| 2157 | F8 | coagulation factor VIII |
| 8029 | CUBN | cubilin |
| 1261 | CNGA3 | cyclic nucleotide gated channel subunit alpha 3 |
| 54798 | DCHS2 | dachsous cadherin-related 2 |
| 1612 | DAPK1 | death associated protein kinase 1 |
| 1756 | DMD | dystrophin |
| 2066 | ERBB4 | erb-b2 receptor tyrosine kinase 4 |
| 221061 | FAM171A1 | family with sequence similarity 171 member A1 |
| 2243 | FGA | fibrinogen alpha chain |

| | | |
|---|---|---|
| 2260 | FGFR1 | fibroblast growth factor receptor 1 |
| 2335 | FN1 | fibronectin 1 |
| 27145 | FILIP1 | filamin A interacting protein 1 |
| 2318 | FLNC | filamin C |
| 56776 | FMN2 | formin 2 |
| 2565 | GABRG1 | gamma-aminobutyric acid type A receptor subunit gamma1 |
| 2678 | GGT1 | gamma-glutamyltransferase 1 |
| 2903 | GRIN2A | glutamate ionotropic receptor NMDA type subunit 2A |
| 2904 | GRIN2B | glutamate ionotropic receptor NMDA type subunit 2B |
| 2913 | GRM3 | glutamate metabotropic receptor 3 |
| 2915 | GRM5 | glutamate metabotropic receptor 5 |
| 3339 | HSPG2 | heparan sulfate proteoglycan 2 |
| 8516 | ITGA8 | integrin subunit alpha 8 |
| 3681 | ITGAD | integrin subunit alpha D |
| 11141 | IL1RAPL1 | interleukin 1 receptor accessory protein like 1 |
| 3802 | KIR2DL1 | killer cell immunoglobulin like receptor, two Ig domains and long cytoplasmic tail 1 |
| 3791 | KDR | kinase insert domain receptor |
| 120892 | LRRK2 | leucine rich repeat kinase 2 |
| 8972 | MGAM | maltase-glucoamylase |
| 23499 | MACF1 | microtubule actin crosslinking factor 1 |
| 10071 | MUC12 | mucin 12, cell surface associated |
| 94025 | MUC16 | mucin 16, cell surface associated |
| 140453 | MUC17 | mucin 17, cell surface associated |
| 4585 | MUC4 | mucin 4, cell surface associated |
| 727897 | MUC5B | mucin 5B, oligomeric mucus/gel-forming |
| 4588 | MUC6 | mucin 6, oligomeric mucus/gel-forming |
| 9378 | NRXN1 | neurexin 1 |
| 26960 | NBEA | neurobeachin |
| 121364 | OR10A7 | olfactory receptor family 10 subfamily A member 7 |
| 403273 | OR5H14 | olfactory receptor family 5 subfamily H member 14 |
| 403274 | OR5H15 | olfactory receptor family 5 subfamily H member 15 |
| 282763 | OR51B5 | olfactory receptor family 51 subfamily B member 5 |
| 81442 | OR6N2 | olfactory receptor family 6 subfamily N member 2 |
| 4978 | OPCML | opioid binding protein/cell adhesion molecule like |
| 5144 | PDE4D | phosphodiesterase 4D |
| 51196 | PLCE1 | phospholipase C epsilon 1 |
| 5334 | inactive | phospholipase C like 1 |
| 5788 | PTPRC | protein tyrosine phosphatase receptor type C |
| 5789 | PTPRD | protein tyrosine phosphatase receptor type D |
| 5800 | PTPRO | protein tyrosine phosphatase receptor type O |
| 5801 | PTPRR | protein tyrosine phosphatase receptor type R |
| 5803 | PTPRZ1 | protein tyrosine phosphatase receptor type Z1 |
| 56112 | PCDHGA3 | protocadherin gamma subfamily A, 3 |
| 65217 | PCDH15 | protocadherin related 15 |
| 5649 | RELN | reelin |
| 5979 | RET | ret proto-oncogene |
| 6261 | RYR1 | ryanodine receptor 1 |
| 6262 | RYR2 | ryanodine receptor 2 |
| 6263 | RYR3 | ryanodine receptor 3 |
| 6323 | SCN1A | sodium voltage-gated channel alpha subunit 1 |
| 6326 | SCN2A | sodium voltage-gated channel alpha subunit 2 |
| 6328 | SCN3A | sodium voltage-gated channel alpha subunit 3 |
| 6335 | SCN9A | sodium voltage-gated channel alpha subunit 9 |
| 64116 | SLC39A8 | solute carrier family 39 member 8 |

| | | |
|---|---|---|
| 8671 | SLC4A4 | solute carrier family 4 member 4 |
| 6546 | SLC8A1 | solute carrier family 8 member A1 |
| 6547 | SLC8A3 | solute carrier family 8 member A3 |
| 8470 | SORBS2 | sorbin and SH3 domain containing 2 |
| 6708 | SPTA1 | spectrin alpha, erythrocytic 1 |
| 6476 | SI | sucrase-isomaltase |
| 259293 | TAS2R30 | taste 2 receptor member 30 |
| 10178 | TENM1 | teneurin transmembrane protein 1 |
| 221981 | THSD7A | thrombospondin type 1 domain containing 7A |
| 80731 | THSD7B | thrombospondin type 1 domain containing 7B |
| 7273 | TTN | titin |
| 7099 | TLR4 | toll like receptor 4 |
| 8989 | TRPA1 | transient receptor potential cation channel subfamily A member 1 |
| 4308 | TRPM1 | transient receptor potential cation channel subfamily M member 1 |
| 140803 | TRPM6 | transient receptor potential cation channel subfamily M member 6 |
| 23352 | UBR4 | ubiquitin protein ligase E3 component n-recognin 4 |
| 440279 | UNC13C | unc-13 homolog C |
| 57578 | UNC79 | unc-79 homolog, NALCN channel complex subunit |
| 285175 | UNC80 | unc-80 homolog, NALCN channel complex subunit |
| 79674 | VEPH1 | ventricular zone expressed PH domain containing 1 |
